# A Randomized Trial of Otilimab in Severe COVID-19 Pneumonia (OSCAR)

**DOI:** 10.1101/2021.04.14.21255475

**Authors:** Jatin Patel, Albertus Beishuizen, Xavier Bocca Ruiz, Hatem Boughanmi, Anthony Cahn, Gerard J Criner, Katherine Davy, Javier de-Miguel-Díez, Sofia Fernandes, Bruno François, Anubha Gupta, Kate Hanrott, Timothy Hatlen, Dave Inman, John D Isaacs, Emily Jarvis, Natalia Kostina, Jean-Claude Lacherade, Pedro Martinez-Ayala, Charlene McEvoy, Rosana Muñoz-Bermúdez, Jessica Neisen, Gaëtan Plantefeve, Lorrie Schifano, Lee Schwab, Zainab Shahid, Michinori Shirano, Julia E. Smith, Eduardo Sprinz, Charlotte Summers, Nicolas Terzi, Mark A Tidswell, Russell Williamson, Duncan Wyncoll, Mark Layton

## Abstract

**BACKGROUND:** Increasing age is a risk factor for COVID-19 severity and mortality; emerging science implicates GM-CSF and dysregulated myeloid cell responses in the pathophysiology of severe COVID-19.

**METHODS:** We conducted a large, global, double-blind, randomized, placebo-controlled study evaluating a single 90 mg infusion of otilimab (human anti-GM-CSF monoclonal) plus standard of care in adults hospitalized with severe COVID-19 respiratory failure and systemic inflammation, stratified by age and clinical status. Primary outcome was the proportion of patients alive and free of respiratory failure at Day 28; secondary endpoints included all-cause mortality at Day 60.

**RESULTS:** Overall, 806 patients were randomized (1:1); 71% of patients receiving otilimab were alive and free of respiratory failure at Day 28 versus 67% receiving placebo, although this did not reach statistical significance (model-adjusted difference 5.3% [95% CI −0.8, 11.4]; p=0.09). However, there was a benefit in the pre-defined ≥70-year age group (model-adjusted difference 19.1% [95% CI 5.2, 33.1]; nominal p=0.009); these patients also had a reduction of 14.4% (95% CI 0.9, 27.9%; nominal p=0.04) in model-adjusted all-cause mortality at Day 60. Safety findings were comparable between otilimab and placebo, and consistent with severe COVID-19.

**CONCLUSIONS:** Although not statistically significant in the overall population, otilimab demonstrated a substantial benefit in patients aged ≥70, possibly reflecting a population that could benefit from therapeutic blocking of GM-CSF in severe COVID-19 where myeloid cell dysregulation is predominant. These findings are being confirmed in a further cohort of patients aged ≥70 in Part 2 of this study. (ClinicalTrials.gov number: NCT04376684).

## Introduction

Reports of severe pneumonia associated with a novel 2019 coronavirus (SARS-CoV-2) began to emerge from Wuhan, China in January 2020. Cases often required intensive care and were associated with significant mortality, with some hospitalized patients having elevated inflammatory cytokines, including granulocyte-monocyte colony-stimulating factor (GM-CSF) and interleukin 6 (IL-6), compared with healthy controls.^1,2^ The severity and spectrum of the disease (COVID-19) resulting from this infection and associated immunopathology has been the subject of intense clinical research spanning trials of a wide range of potential therapeutic agents.^3-11^ Severe disease is characterized by systemic inflammation, dysregulated myeloid cell responses, and respiratory and/or cardiovascular failure.^12-16^ Older age and chronic health conditions, such as obesity, diabetes and cardiovascular disease, are risk factors for severe disease.^17,18^ In particular, increasing age is strongly associated with worsened disease severity, intensive care unit (ICU) admission, and increased mortality.^19-21^

GM-CSF has been implicated as a key cytokine in driving and perpetuating the hyperinflammation observed in severe COVID-19.^22-25^ Otilimab is a well-tolerated and effective, high-affinity, anti–GM-CSF monoclonal antibody that has been shown to reduce inflammatory disease activity in rheumatoid arthritis (RA).^26^ We designed a double-blind, placebo-controlled, randomized clinical trial (OSCAR [Otilimab in Severe COVID-19 Related Disease]; ClinicalTrials.gov identifier NCT04376684) to investigate whether otilimab was able to improve clinical outcomes in patients hospitalized with respiratory failure and systemic inflammation secondary to severe COVID-19. During the study design, we aimed to address the multiple challenges of conducting a high-quality clinical trial within the context of a pandemic. We were conscious of the large number of uncontrolled, cohort, open-label, and small therapeutic trials that had been initiated, and the frequent changes in new treatments introduced into clinical practice, often with minimal data to support their emergency use. We designed our protocol to minimize the burden on researchers where possible, to reflect ethnic diversity, and adapted it as necessary to evolving standards of care.

## Methods

### Study design and patient selection

Enrollment into Part 1 of this double-blind, placebo-controlled study began on 28 May 2020 and ended on 15 November 2020, with the last patient completing Day 60 on 13 January 2021. Patients were recruited from 108 study sites in 17 countries: Argentina (6), Belgium (2), Brazil (4), Canada (4), Chile (1), France (12), India (10), Japan (8), Mexico (4), the Netherlands (5), Peru (3), Poland (5), Russian Federation (8), South Africa (5), Spain (7), the United Kingdom (4), and the United States (20) (appendix 1). The study enrolled hospitalized adult patients (≥18 to ≤79 years of age), with confirmed SARS-CoV-2 pneumonia, new onset hypoxemia requiring any of high-flow oxygen (≥15 L/min), non-invasive ventilation or mechanical ventilation (MV; duration ≤48 hours prior to dose), and increased biological markers of systemic inflammation (C-reactive protein or serum ferritin above local upper limit of normal). Key exclusion criteria were: death considered likely within 48 hours, multiple organ failure according to the investigator’s opinion or a Sequential Organ Failure Assessment (SOFA) score >10 if in the ICU, or use of extracorporeal membrane oxygenation, hemofiltration/dialysis, high-dose (>0.15 μg/kg/min) noradrenaline (or equivalent) or >1 vasopressor. Eligible patients were planned to be in clinical status Category 5 or 6 according to the GSK-modified WHO ordinal scale^27^ where: 1 = not hospitalized, no limitation of activity; 2 = not hospitalized, limitation of activity; 3 = hospitalized, no oxygen therapy; 4 = hospitalized, low-flow oxygen by mask or nasal prongs; 5 = hospitalized, high-flow oxygen (≥15 L/min), continuous positive airway pressure, bilevel positive airway pressure non-invasive ventilation; 6 = hospitalized, intubation and MV; 7 = hospitalized, MV plus additional organ support; 8 = death. Additional details are provided in the protocol.

As a precaution, a safety cohort was planned, whereby initial dosing for the first four Category 5 patients was staggered and safety data were independently reviewed prior to enrollment of an initial safety cohort of 16 additional Category 5 patients, before the study progressed to the main cohort of patients.

### Study treatments

Patients were centrally randomized 1:1 by interactive response technology in a blinded manner to either a single one-hour intravenous (IV) infusion of otilimab 90 mg (aiming to achieve serum otilimab concentrations through to Day 7 similar to steady-state trough in patients with RA) or placebo (saline). In the safety cohort a randomization block size of two was used for the first and second sentinel pairs, and four for the remaining 16 patients. In the main cohort, patients were stratified by clinical status (Category 5 or 6) and age group (<60, 60 to 69, and ≥70 years) with a block size of four.

An unblinded pharmacist dispensed the study intervention, ensuring no differences in labelling or time taken to dispense between the two interventions. Investigators, who enrolled the patients, and the patients remained blinded to assigned study intervention throughout the study. In addition, all patients received standard care according to institutional protocols. Medications not permitted prior to enrollment or during the study included monoclonal antibodies (*e*.*g*. tocilizumab, sarilumab), immunosuppressants, and chronic oral corticosteroid use >10 mg/day prednisone equivalent for a non-COVID-19–related condition. Additional or prior medications for COVID-19–related disease were permitted if part of local institutional policies and not part of a clinical trial.

### Assessments and outcome measures

Patients were assessed daily until discharge from the investigational site or Day 28, whichever was sooner, in addition to follow up at Days 42 and 60. Clinical status was assessed according to the GSK-modified WHO ordinal scale as described above. In addition to standard care, data were collected on ventilation status and the clinical features of COVID-19, with blood samples collected for pharmacokinetic and pharmacodynamic biomarker analysis.

The primary endpoint was the proportion of patients alive and free of respiratory failure (clinical status Categories 1, 2, 3, or 4) at Day 28. Secondary efficacy outcomes were: all-cause mortality at Day 60 (all-cause mortality at Day 28 was defined post hoc); time to all-cause mortality up to Day 60; proportion of patients alive and free of respiratory failure at Days 7, 14, 42, and 60; time to recovery from respiratory failure up to Day 28; proportion of patients alive and independent of supplementary oxygen (clinical status Categories 1, 2, or 3) at Days 7, 14, 28, 42, and 60; time to last dependence on supplementary oxygen up to Day 28; admission to ICU up to Day 28; time to final ICU discharge up to Day 28; and, revised prior to unblinding from “time to final hospital discharge up to Day 28”, to time to first discharge from investigational site up to Day 60, and time to first discharge to a non-hospital residence up to Day 60. Exploratory endpoints are listed in the protocol. Occurrence of adverse events (AEs) and serious adverse events (SAEs) was reported up to Day 60 and recorded according to the system organ class and preferred terms in the Medical Dictionary for Regulatory Activities (MedDRA), v23.1.

### Statistical analysis

The study used a group sequential design, using a Lan-DeMets alpha-spending function to control the type I error with four interim analyses planned for futility using Pocock analogue rules, and two interim analyses planned for efficacy using the O’Brien-Fleming analogue rules.^28^ The final interim analysis was not performed as the study completed recruitment prior to the data-cut being reached. Full details of the design parameters and decision criteria are included in the statistical analysis plan.

A sample size of 800 patients provided approximately 90% power to detect a difference of 12% in the proportion of patients alive and free of respiratory failure at a one-sided 2.5% significance level and an assumed placebo response rate of 45%.

The primary endpoint was analyzed using logistic regression adjusting for treatment, age group and clinical status (Category 5 and Category 6) at baseline. Missing data in the overall primary analysis were imputed using multiple imputation, assuming data are missing at random, and adjusting for analysis covariates. The primary endpoint was also analyzed by pre-defined subgroups of clinical status (Categories 5 and 6) and age group (<60, 60 to 69, and ≥70 years), as detailed in the statistical analysis plan.

The primary population for analysis included all patients who were randomized and received study drug (modified intent-to-treat [mITT]). Results are presented with two-sided p-values. Additional details are provided in the statistical analysis plan.

### Study oversight

The study was conducted in accordance with the Declaration of Helsinki, Council for International Organizations of Medical Sciences International Ethical Guidelines, International Conference on Harmonization, Good Clinical Practice, and applicable country-specific regulatory requirements. The protocol was approved by relevant institutional review boards (IRB/IEC). Before patient enrollment, informed consent was obtained written/orally from the patient or written/orally from the patient’s legally authorized representative. An Independent Data Monitoring Committee (IDMC) monitored in-stream unblinded safety and efficacy data throughout the study and at pre-defined interim analyses.

## Results

### Patients

A total of 851 patients were screened, 806 were randomized, and 793 included in the mITT population (395 in the otilimab group and 398 in the placebo group) (Figure 1). Within the pre-defined stratification groups, 630 (78%) were in clinical status Category 5 and 176 (22%) in Category 6; with 363 (45%), 262 (33%), and 181 (22%) in the <60, 60 to 69, and ≥70-year age groups, respectively. The safety population included 397 treated with otilimab and 396 in the placebo group, because two patients randomized to placebo were incorrectly dosed with otilimab. By Day 60, 379/403 (94%) patients in the otilimab group and 388/403 (96%) on placebo either completed the study or had died. None of the patients discontinued participation because of safety reasons.

**Figure 1.**
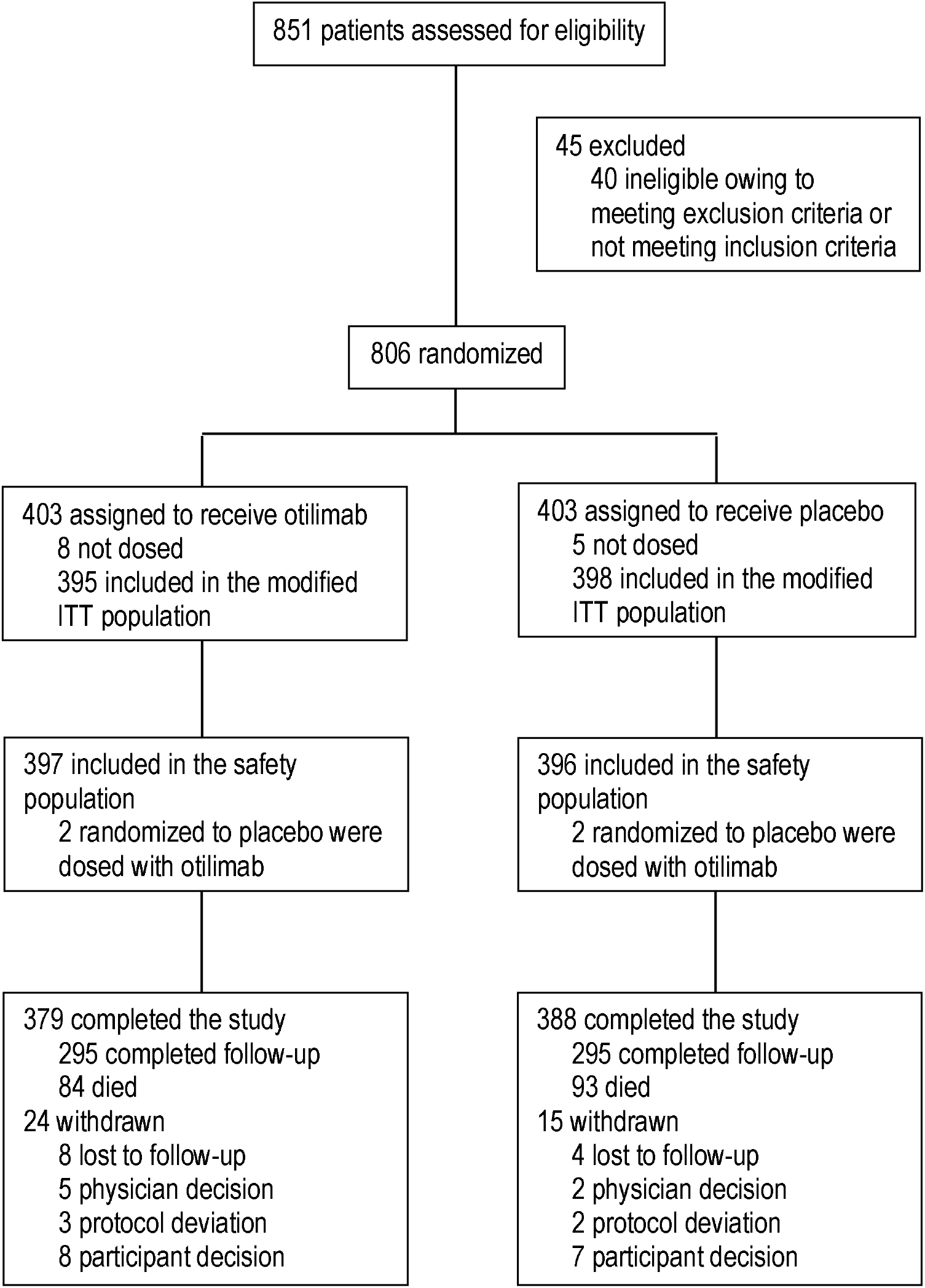
CONSORT diagram.

Baseline demographic and disease characteristics were generally well balanced between the two study groups and were reflective of severe COVID-19 with elevated levels of markers of systemic inflammation and GM-CSF (Table 1) compared with healthy controls (data not shown). The mean (± standard deviation) age was 59.8 (±11.7) years in the otilimab group and 59.4 (±11.9) years in the placebo group. Most patients were pre-treated with COVID-19 medications; specifically, 83% received corticosteroids (including dexamethasone), 34% received remdesivir, and 6% received convalescent plasma prior to randomization. Two patients (<1%) in the otilimab group and 7 (2%) in the placebo group received tocilizumab post-study treatment, and none received sarilumab.

**Table 1.**
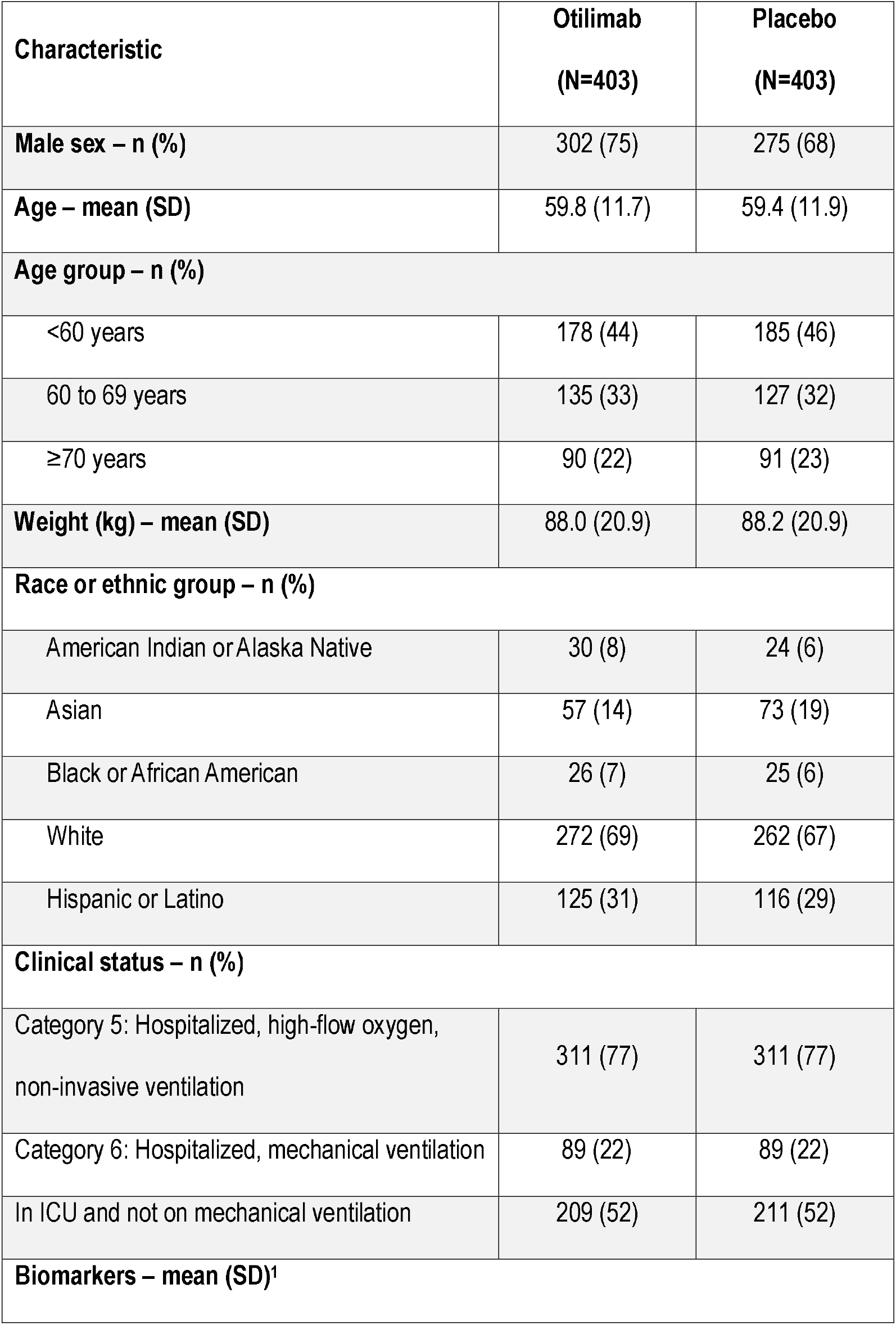
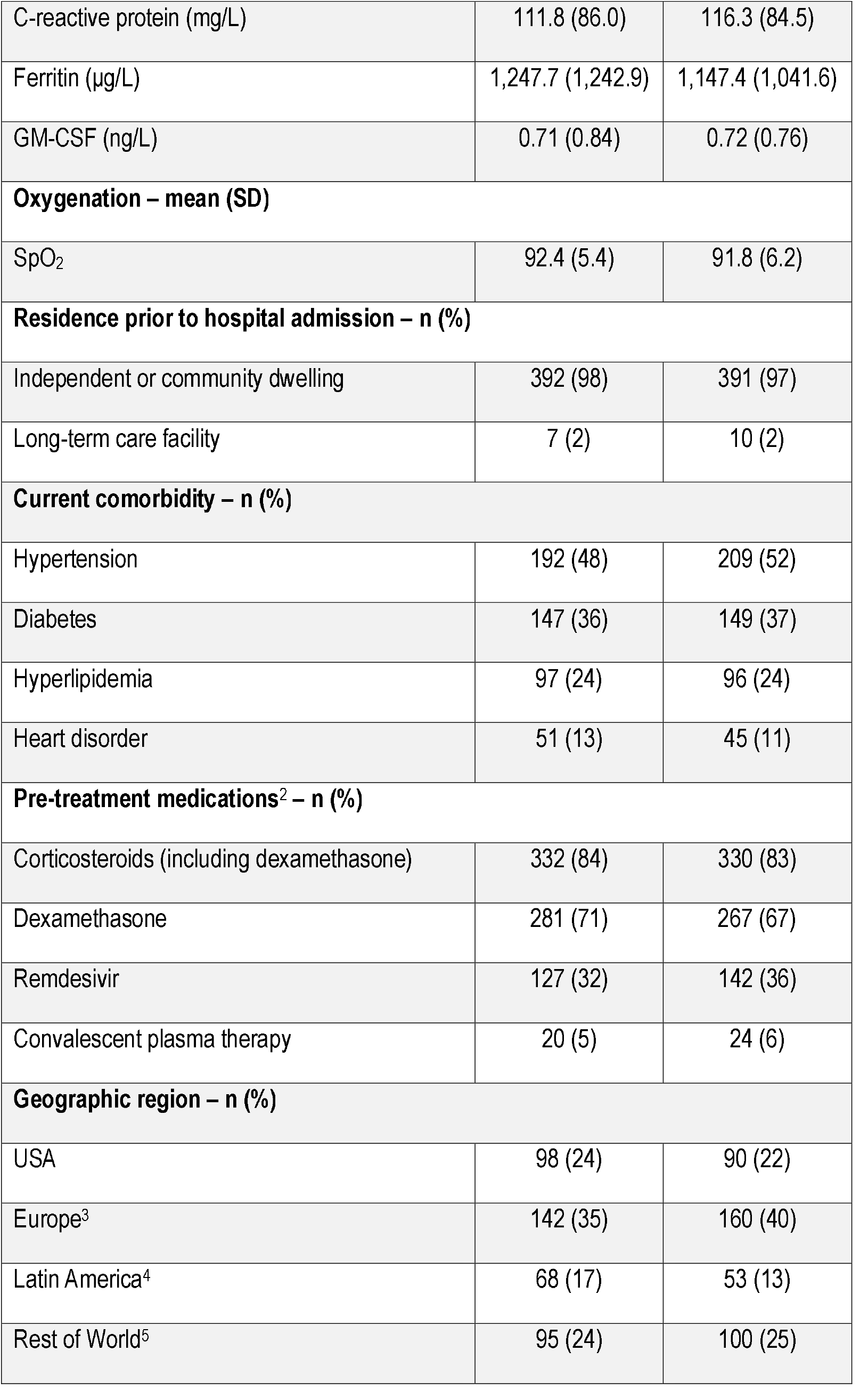
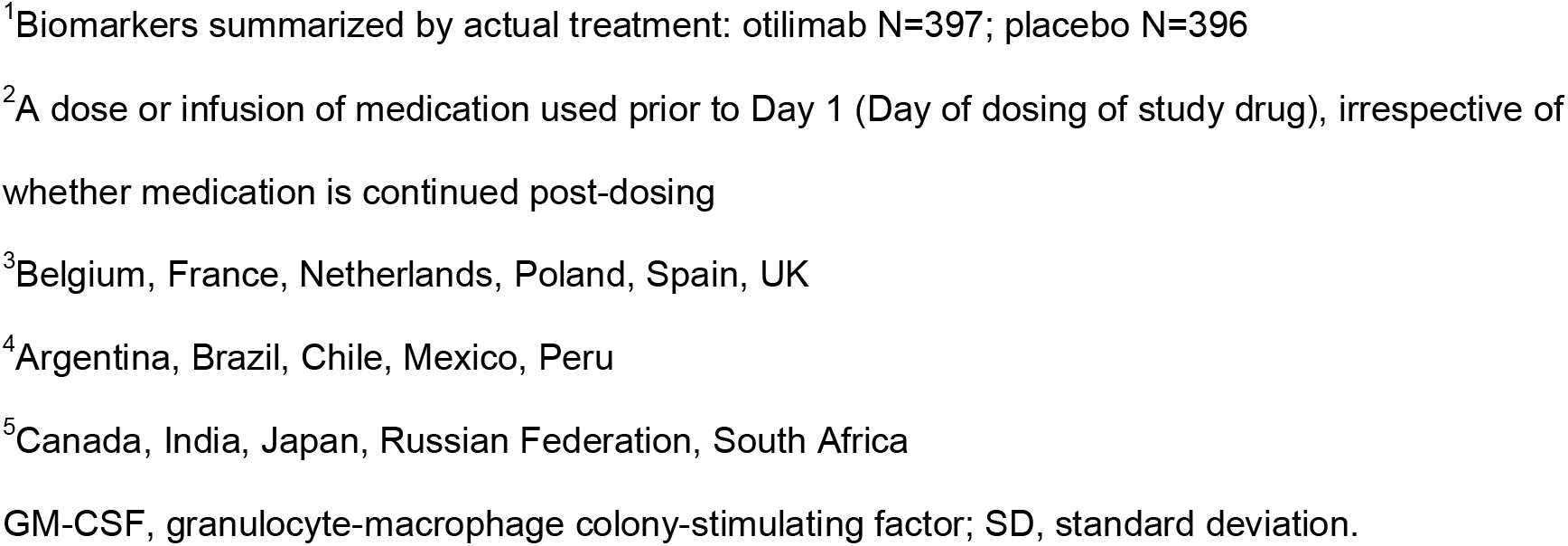
Demographic and clinical characteristics of patients at baseline.

| **Characteristic** | **Otilimab**<br>**(N=403)** | **Placebo**<br>**(N=403)** |
| --- | --- | --- |
| **Male sex – n (%)** | 302 (75) | 275 (68) |
| **Age – mean (SD)** | 59.8 (11.7) | 59.4 (11.9) |
| **Age group – n (%)** |  |  |
| <60 years | 178 (44) | 185 (46) |
| 60 to 69 years | 135 (33) | 127 (32) |
| ≥70 years | 90 (22) | 91 (23) |
| **Weight (kg) – mean (SD)** | 88.0 (20.9) | 88.2 (20.9) |
| **Race or ethnic group – n (%)** |  |  |
| American Indian or Alaska Native | 30 (8) | 24 (6) |
| Asian | 57 (14) | 73 (19) |
| Black or African American | 26 (7) | 25 (6) |
| White | 272 (69) | 262 (67) |
| Hispanic or Latino | 125 (31) | 116 (29) |
| **Clinical status – n (%)** |  |  |
| Category 5: Hospitalized, high-flow oxygen, non-invasive ventilation | 311 (77) | 311 (77) |
| Category 6: Hospitalized, mechanical ventilation | 89 (22) | 89 (22) |
| In ICU and not on mechanical ventilation | 209 (52) | 211 (52) |
| **Biomarkers – mean (SD)<sup>1</sup>** |  |  |
| C-reactive protein (mg/L) | 111.8 (86.0) | 116.3 (84.5) |
| Ferritin (µg/L) | 1,247.7 (1,242.9) | 1,147.4 (1,041.6) |
| GM-CSF (ng/L) | 0.71 (0.84) | 0.72 (0.76) |
| **Oxygenation – mean (SD)** |  |  |
| SpO<sub>2</sub> | 92.4 (5.4) | 91.8 (6.2) |
| **Residence prior to hospital admission – n (%)** |  |  |
| Independent or community dwelling | 392 (98) | 391 (97) |
| Long-term care facility | 7 (2) | 10 (2) |
| **Current comorbidity – n (%)** |  |  |
| Hypertension | 192 (48) | 209 (52) |
| Diabetes | 147 (36) | 149 (37) |
| Hyperlipidemia | 97 (24) | 96 (24) |
| Heart disorder | 51 (13) | 45 (11) |
| **Pre-treatment medications<sup>2</sup> – n (%)** |  |  |
| Corticosteroids (including dexamethasone) | 332 (84) | 330 (83) |
| Dexamethasone | 281 (71) | 267 (67) |
| Remdesivir | 127 (32) | 142 (36) |
| Convalescent plasma therapy | 20 (5) | 24 (6) |
| **Geographic region – n (%)** |  |  |
| USA | 98 (24) | 90 (22) |
| Europe<sup>3</sup> | 142 (35) | 160 (40) |
| Latin America<sup>4</sup> | 68 (17) | 53 (13) |
| Rest of World<sup>5</sup> | 95 (24) | 100 (25) |

<sup>1</sup>Biomarkers summarized by actual treatment: otilimab N=397; placebo N=396
<sup>2</sup>A dose or infusion of medication used prior to Day 1 (Day of dosing of study drug), irrespective of whether medication is continued post-dosing
<sup>3</sup>Belgium, France, Netherlands, Poland, Spain, UK
<sup>4</sup>Argentina, Brazil, Chile, Mexico, Peru
<sup>5</sup>Canada, India, Japan, Russian Federation, South Africa
GM-CSF, granulocyte-macrophage colony-stimulating factor; SD, standard deviation.

### Primary outcome

Although 71% of patients in the otilimab group were alive and free of respiratory failure at Day 28 compared with 67% on placebo (Figure 2), the model-adjusted difference of 5.3% did not reach statistical significance (95% CI −0.8, 11.4; p=0.09). Similar model-adjusted treatment differences of 5.9% (95% CI −0.8, 12.7) and 4.6% (95% CI −9.6, 18.8) were observed for patients in Categories 5 and 6 (Figure 2). However, although the pre-defined subgroup analysis by age revealed little benefit in patients aged <60 or 60 to 69 years, there was a nominally significant model-adjusted difference of 19.1% (95% CI 5.2, 33.1; p=0.009) in patients ≥70 years (Figure 2).

**Figure 2.**
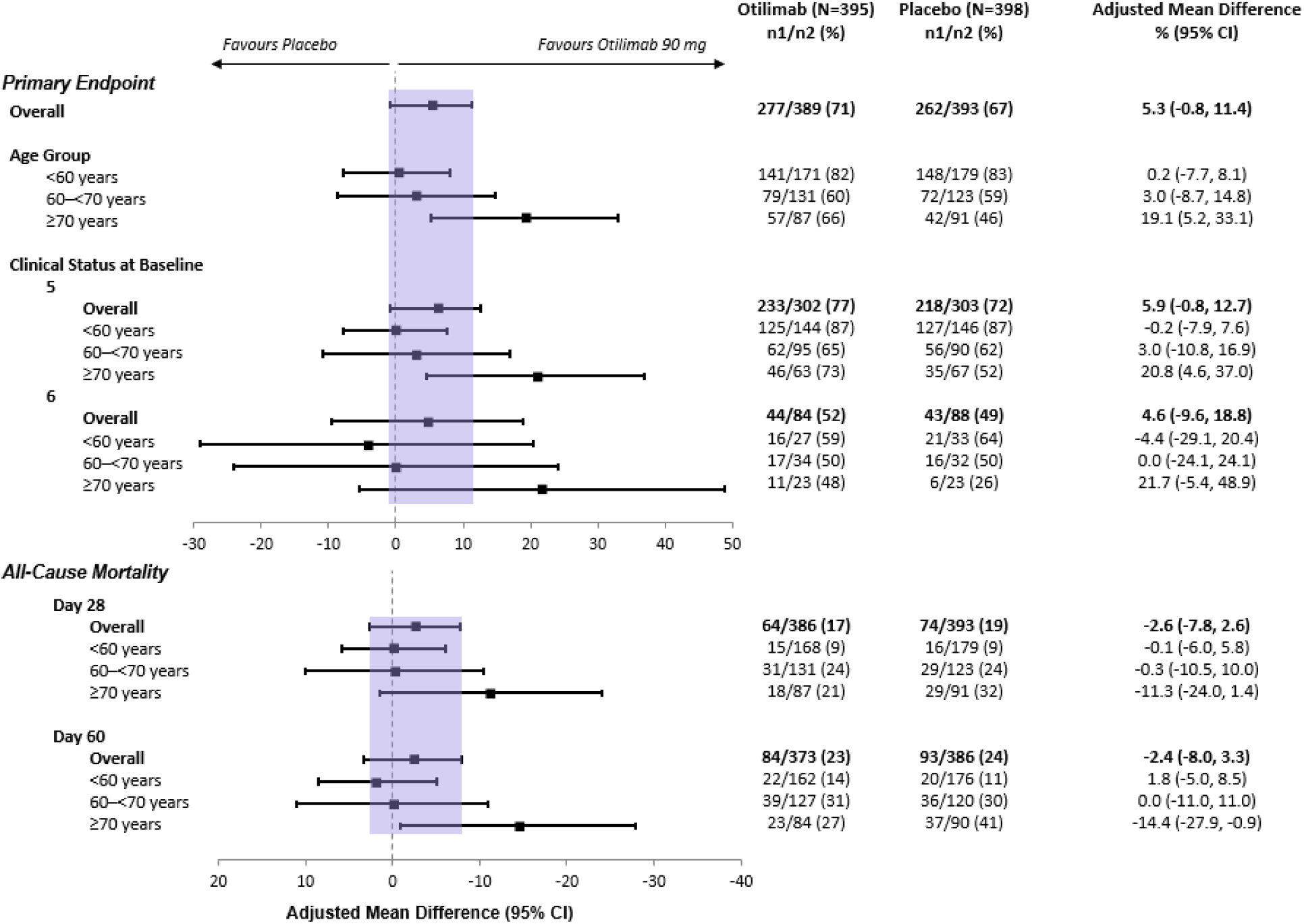
Proportion of patients alive and free of respiratory failure at Day 28 (primary endpoint), and all-cause mortality at Day 28 and Day 60. CI, confidence interval; n1, number of patients with the event; n2, number of patients with non-missing data at timepoint

Although *post-hoc* analyses suggested a greater benefit with otilimab in females and patients with comorbidities (particularly hypertension), there was little impact of geographic region or pre-treatment (and mostly continued) use of dexamethasone (Figure S1A; appendix 2). However, the response in patients aged ≥70 years was consistent regardless of disease severity (Figure 2), dexamethasone use, or comorbidities (Figure S1B; appendix 2).

### All-cause mortality

In the mITT population, all-cause mortality at Day 60 was 23% in the otilimab group compared with 24% on placebo (model-adjusted difference −2.4% [95% CI −8.0, 3.3]; p=0.41) (Figure 2). However, in the ≥70-year age group there was lower mortality at Day 60 with otilimab (27%) versus placebo (41%) (model-adjusted difference −14.4% [95% CI −27.9, −0.9]; nominal p=0.04). Additionally, there was a trend of reduced mortality at Day 28 with otilimab in a *post-hoc* analysis showing a model-adjusted difference of −11.3% (95% CI −24.0, 1.4; p=0.09) in this subgroup (Figure 2).

### Additional *post-hoc* secondary and exploratory efficacy endpoints

All other secondary and exploratory efficacy endpoints were analyzed for the mITT population and also *post hoc* by age groups. In the ≥70-year age group, there was a greater proportion of responders (Figures 3A-L) with otilimab versus placebo for all secondary endpoints. Treatment effects were generally apparent 7–10 days after dosing (Figures 3A-L). For the exploratory endpoint of change from baseline in fraction of inspired oxygen (FiO_2_) a greater reduction was observed in patients on otilimab compared to those on placebo in both the mITT population and ≥70-year group (Figures 3M and N).

**Figure 3.**
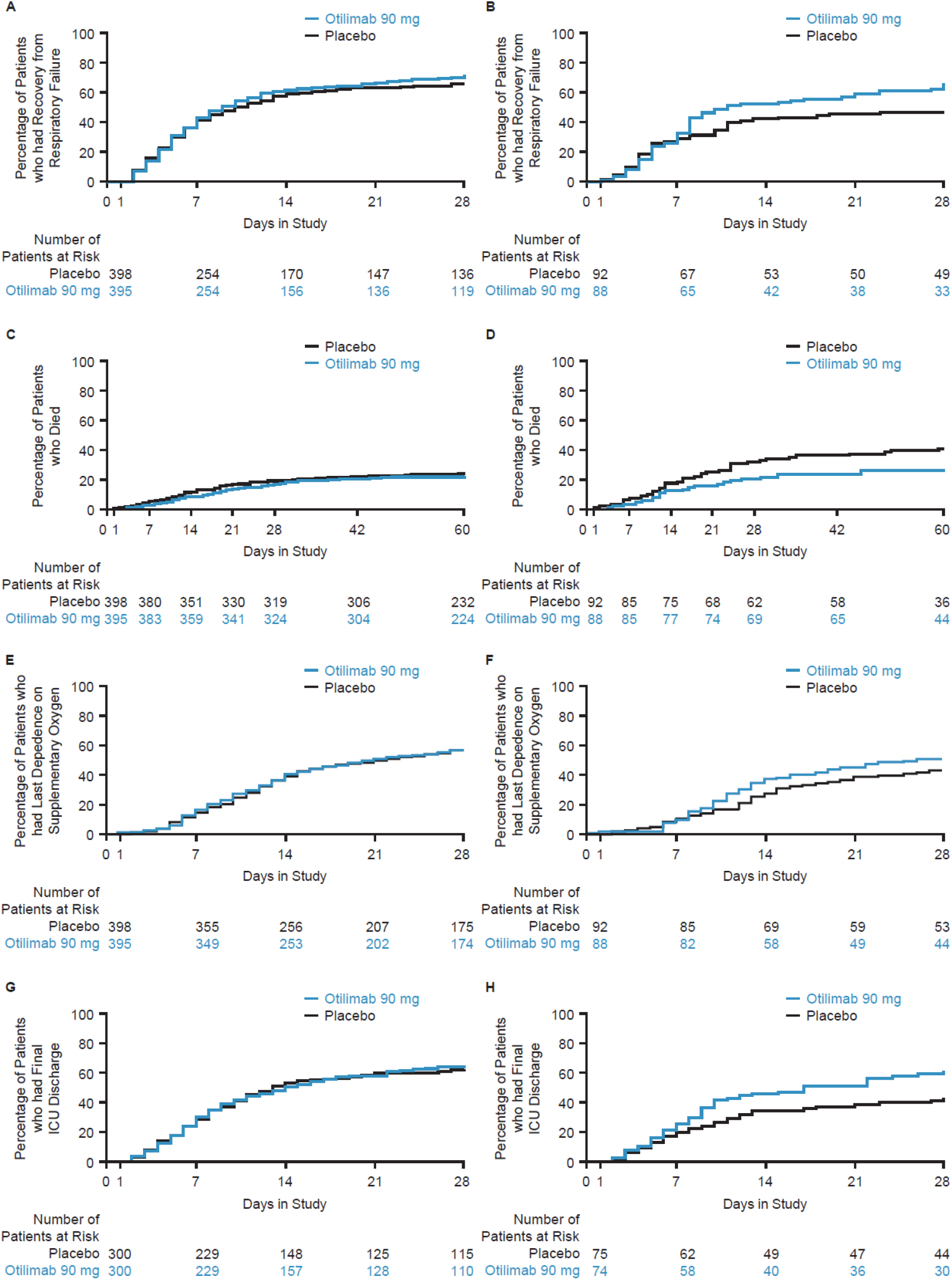
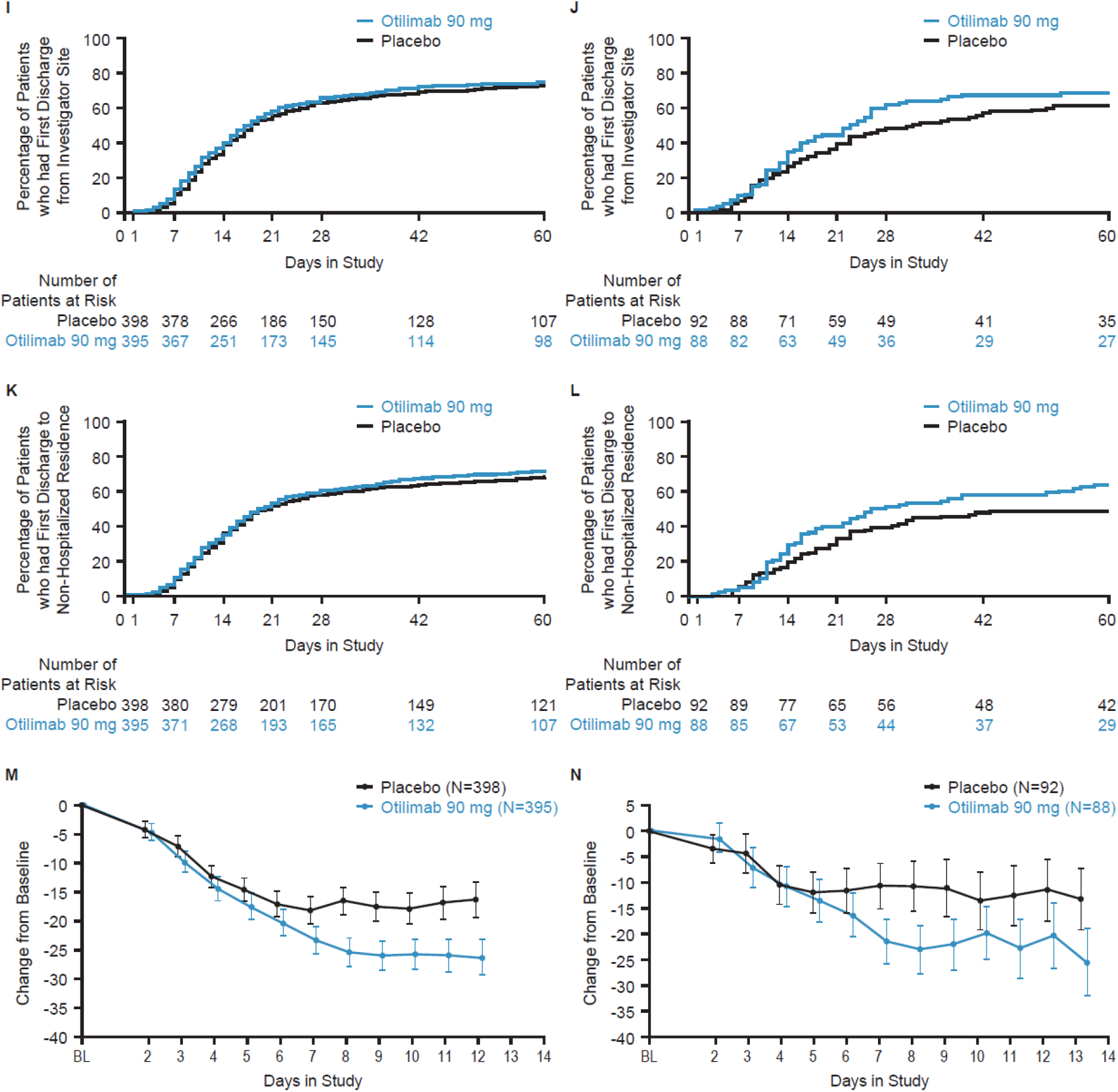
Key secondary endpoints and exploratory endpoint (time to event analyses) Secondary endpoints: Kaplan-Meier time to recovery from respiratory failure up to Day 28 in the mITT population (A) and ≥70 year group (B); Kaplan-Meier time to all-cause mortality up to Day 60 in the mITT population (C) and ≥70 year group (D); Kaplan-Meier time to last dependence on supplementary oxygen up to Day 28 in the mITT population (E) and ≥70 year group (F); Kaplan-Meier time to final ICU discharge up to Day 28 in the mITT ICU population (G) and ≥70 year group (H); Kaplan-Meier time to First Discharge from Investigator Site up to Day 60 in the mITT population (I) and ≥70 year group (J); Kaplan-Meier time to final hospital discharge (to Non-Hospitalized Residence) up to Day 60 in the mITT population (K) and ≥70 year group (L); exploratory endpoint: Mean Change from Baseline (95% CI) in Concentration of Inspired Oxygen (FiO_2_) Trimmed^1^ Sample in the mITT population (M) and ≥70 year group (N) ^1^Analyzed using the trimmed means approach^37^ where the proportion of data to be trimmed was determined by the amount of missing data due to intercurrent events. Results presented until >50% of population experience the intercurrent event.

### Pharmacokinetics

A single dose of otilimab resulted in mean C_max_ of 20.2 μg/mL after dosing on Day 1 and 1.9 μg/mL on Day 7, with coefficient of variation of 43% and 69% respectively; similar levels were achieved in the ≥70 age group (Supplementary Figure S2). In the otilimab arm, GM-CSF levels at Day 2, proximal to C_max_, were reduced by 95% to a mean of 0.037 ng/L with 255/381 (67%) samples falling below the assay lower limit of quantification (0.036 ng/L); levels in the placebo arm remained unchanged (data not shown).

### Safety

Overall, safety findings, including the scope of AEs and SAEs, were reflective of the severe COVID-19 population. Of note, an overall pattern of increasing frequency of AEs and SAEs was observed with advancing age irrespective of treatment assignment. The most common AEs and SAEs in the safety population and each age subgroup (defined post hoc) are listed in Table 2. No safety signals related to treatment with otilimab were identified.

**Table 2.**
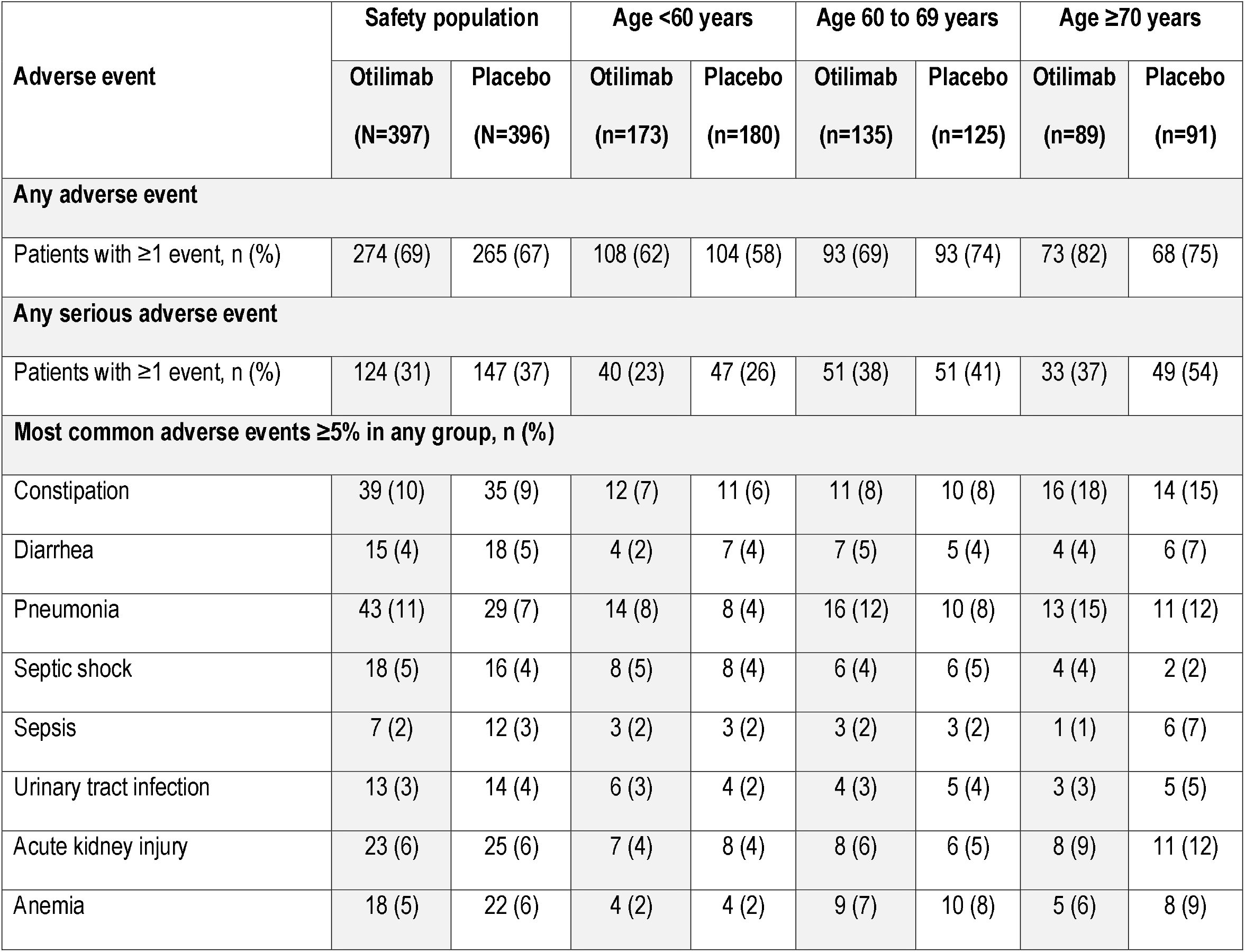
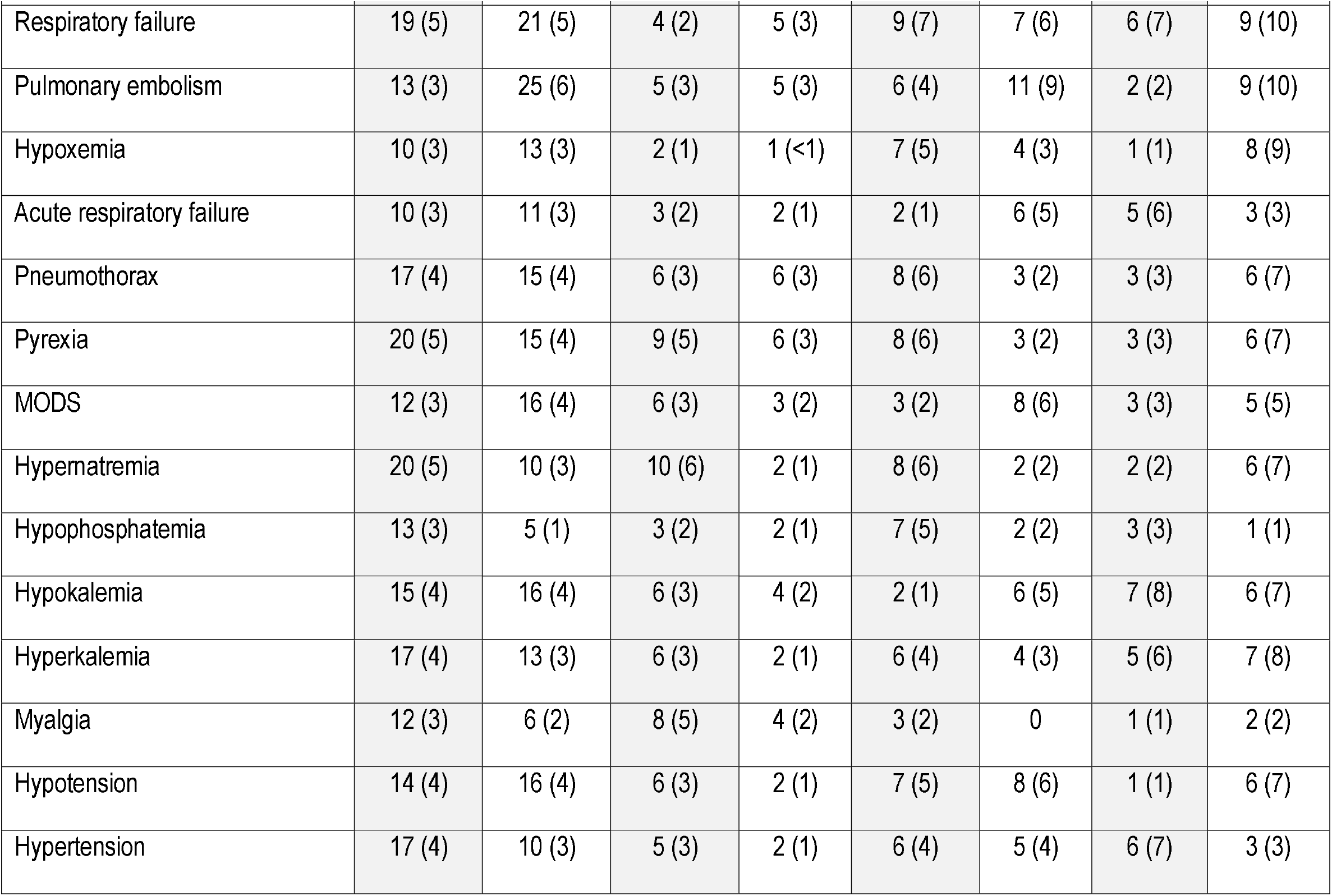
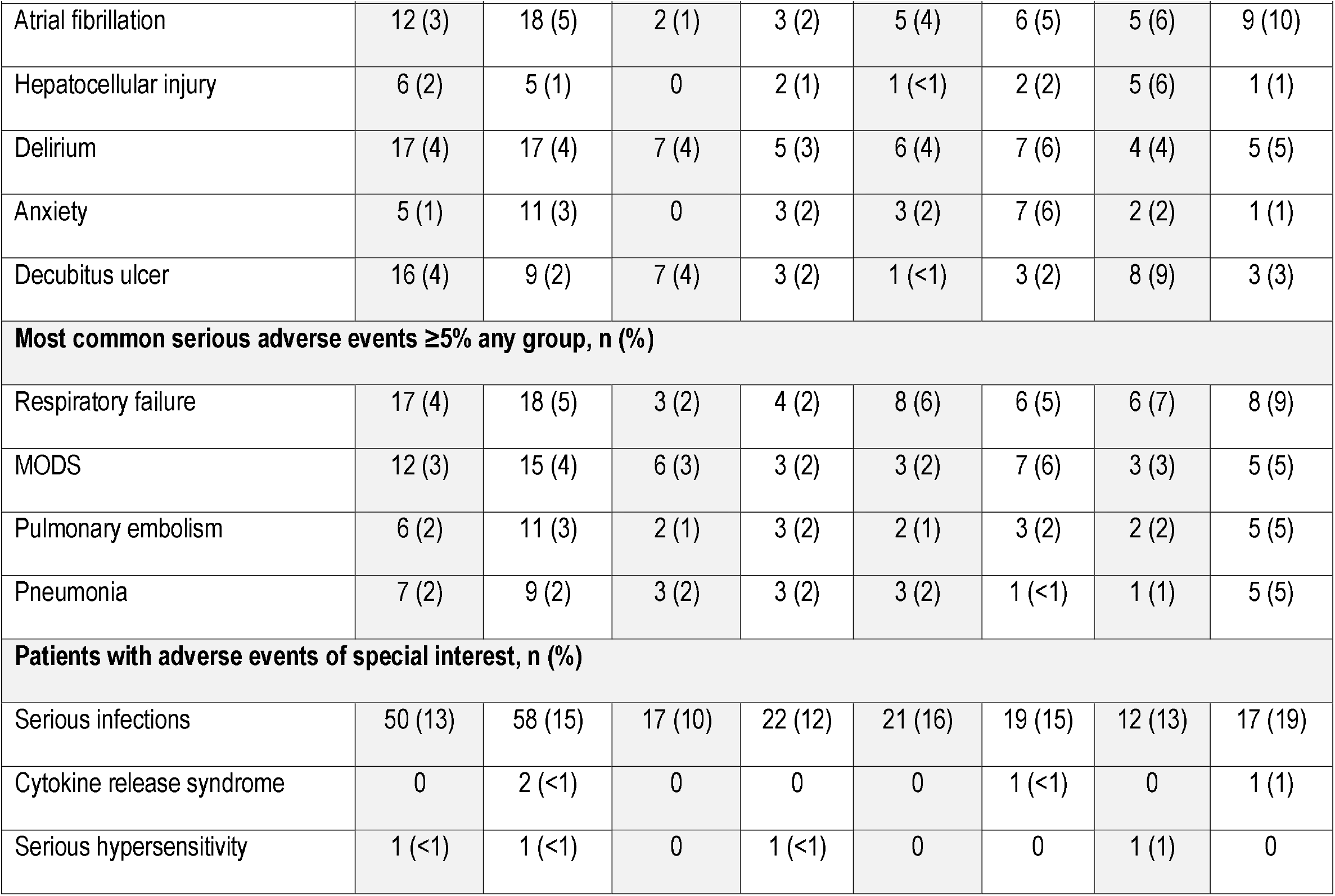
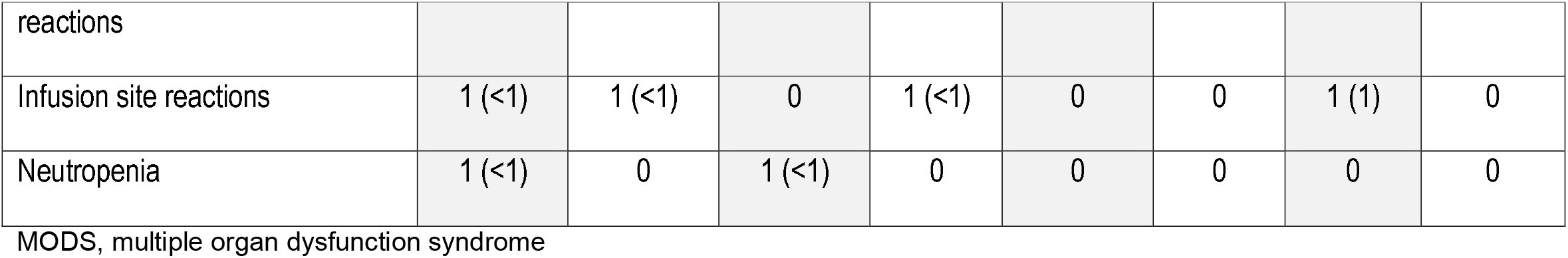
Adverse Events reported by preferred term occurring in ≥5% of patients (safety population and by age group)

In patients ≥70 years who received otilimab, there were reduced frequencies of investigator-reported hypoxemia and pulmonary embolism compared with placebo. This observation may be linked to the greater recovery from respiratory failure with otilimab seen in this ≥70-year subgroup of patients compared with the total safety population.

## Discussion

This double-blind, placebo-controlled, randomized clinical trial in hospitalized adults with severe COVID-19 pneumonia (clinical status Categories 5 and 6) showed that otilimab was associated with a model-adjusted increase of 5.3% in the proportion of patients alive and free of respiratory failure (clinical status Categories 1, 2, 3, or 4) at Day 28. Although this result did not reach statistical significance in the overall population, in the pre-defined subgroup of patients aged ≥70 years, treatment with otilimab resulted in significantly more patients alive and free of respiratory failure at Day 28 than those on placebo (by a model-adjusted difference of 19.1%), and a corresponding decrease in all-cause mortality at Day 60. The safety findings in all age groups were as expected for a population with severe COVID-19 pneumonia, with the most common SAE being respiratory failure. Fewer SAEs were observed in the subgroup of patients ≥70 years treated with otilimab versus placebo, notably including pulmonary embolism and hypoxemia.

The biological plausability of the observed clinical benefit in patients ≥70 years is supported by the observation that older patients may be predisposed to inappropriate, myeloid cell-driven hyperinflammation as a consequence of normal aging of the immune system (immunosenescence and inflammaging).^29,30^ The marked systemic inflammation associated with severe COVID-19 may, at least in part, be driven by dysregulated myeloid cells,^31-36^ which may be enhanced by the early upregulation of GM-CSF in patients >70 years.^24^ Therefore older patients, who are least likely to recover with standard of care alone, may preferentially benefit from targeting the dysregulated myeloid cell responses observed in severe COVID-19 by inhibiting GM-CSF compared with younger patients.

The treatment benefits oberved in the ≥70 year group were apparent across the majority of the secondary endpoints assessed over time, and were generally maintained to Day 60. The observed trends in improved oxygenation response to otilimab may be reflective of an early mechanistic response.

Assumptions made about placebo group responses during the design of the study in March 2020 proved to significantly underestimate the observed response (67% *cf*. 45%). This difference likely reflects the substantial improvements in the standard of care of COVID-19 during our study timeframe, including ventilation practices and corticosteroid use, amongst many other changes to patient management and healthcare resourcing.

At the time of writing, severe COVID-19 is still prevalent in older adults (with additional waves in many countries), and the high mortality in this age group supports the urgent need for additional treatment options for these most at-risk patients. We have therefore expedited enrollment of an additional cohort of patients ≥70 years with severe COVID-19 to urgently confirm these findings in an extension of the OSCAR study.

## Supporting information

CONSORT checklist

Reporting and analysis plan

Protocol

## Data Availability

GSK makes available anonymized individual participant data and associated documents from interventional clinical studies, which evaluate medicines upon approval of proposals submitted to www.clinicalstudydatarequest.com. To access original data for studies that have been re-analyzed, other types of GSK sponsored research, for study documents without patient-level data and for clinical studies not listed, please submit an enquiry via the website.

## Acknowledgments

The authors thank the patients and their families who participated in this clinical study, as well as the doctors, nurses, pharmacists, and other allied health professionals at all the sites.

Medical writing support, in the form of editorial checks, figure development and assistance with submission, was provided by Clare Slater, PhD CMPP, and Leigh O’Connor-Jones, PhD, of Fishawack Indicia Ltd. UK, part of Fishawack Health (funded by GSK).

## Supplementary material

Investigator and site list (appendix 1), figures S1 and S2 (appendix 2), protocol, statistical analysis plan

## References

1. Huang C, Wang Y, Li X, et al. Clinical features of patients infected with 2019 novel coronavirus in Wuhan, China. Lancet 2020;395:497–506.

2. Zhou Y, Fu B, Zheng X, et al. Pathogenic T cells and inflammatory monocytes incite inflammatory storm in severe COVID-19 patients. Nat Sci Rev 2020:nwaa041.

3. Beigel JH, Tomashek KM, Dodd LE, et al. Remdesivir for the Treatment of Covid-19 - Final Report. N Engl J Med 2020;383:1813–26.

4. Cao B, Wang Y, Wen D, et al. A Trial of Lopinavir-Ritonavir in Adults Hospitalized with Severe Covid-19. N Engl J Med 2020;382:1787–99.

5. De Luca G, Cavalli G, Campochiaro C, et al. GM-CSF blockade with mavrilimumab in severe COVID-19 pneumonia and systemic hyperinflammation: a single-centre, prospective cohort study. Lancet Rheumatol 2020;2:e465–e73.

6. Horby P, Mafham M, Linsell L, et al. Effect of Hydroxychloroquine in Hospitalized Patients with Covid-19. N Engl J Med 2020;383:2030–40.

7. Horby P, Lim WS, Emberson JR, et al. Dexamethasone in Hospitalized Patients with Covid-19. N Engl J Med 2021;384:693–704.

8. Horby PW, Pessoa-Amorim G, Peto L, et al. Tocilizumab in patients admitted to hospital with COVID-19 (RECOVERY): preliminary results of a randomised, controlled, open-label, platform trial. medRxiv Posted online February 11, 2021:DOI: 10.1101/2021.02.11.21249258.

9. Kalil AC, Patterson TF, Mehta AK, et al. Baricitinib plus Remdesivir for Hospitalized Adults with Covid-19. N Engl J Med 2021;384:795–807.

10. Rosas IO, Brau N, Waters M, et al. Tocilizumab in Hospitalized Patients with Severe Covid-19 Pneumonia. N Engl J Med 2021:DOI: 10.1056/NEJMoa2028700.

11. Temesgen Z, Assi M, Shweta FNU, et al. GM-CSF Neutralization With Lenzilumab in Severe COVID-19 Pneumonia: A Case-Cohort Study. Mayo Clin Proc 2020;95:2382–94.

12. Carsana L, Sonzogni A, Nasr A, et al. Pulmonary post-mortem findings in a series of COVID-19 cases from northern Italy: a two-centre descriptive study. Lancet Infect Dis 2020;20:1135–40.

13. Nienhold R, Ciani Y, Koelzer VH, et al. Two distinct immunopathological profiles in autopsy lungs of COVID-19. Nat Commun 2020;11:5086.

14. Chen T, Wu D, Chen H, et al. Clinical characteristics of 113 deceased patients with coronavirus disease 2019: retrospective study. BMJ 2020;368:m1091.

15. Schurink B, Roos E, Radonic T, et al. Viral presence and immunopathology in patients with lethal COVID-19: a prospective autopsy cohort study. Lancet Microbe 2020;1:e290–e9.

16. Wang S, Fu L, Huang K, Han J, Zhang R, Fu Z. Neutrophil-to-lymphocyte ratio on admission is an independent risk factor for the severity and mortality in patients with coronavirus disease 2019. The Journal of infection 2021;82:e16–e8.

17. Grasselli G, Zangrillo A, Zanella A, et al. Baseline Characteristics and Outcomes of 1591 Patients Infected With SARS-CoV-2 Admitted to ICUs of the Lombardy Region, Italy. JAMA 2020;323:1574–81.

18. Mani VR, Kalabin A, Valdivieso SC, Murray-Ramcharan M, Donaldson B. New York Inner City Hospital COVID-19 Experience and Current Data: Retrospective Analysis at the Epicenter of the American Coronavirus Outbreak. J Med Internet Res 2020;22:e20548.

19. ICNARC report on COVID-19 in critical care. ICNARC, 2021. (Accessed 19 March, 2021, at https://www.icnarc.org/DataServices/Attachments/Download/8b3a0f95-de88-eb11-912e-00505601089b.)

20. Imam Z, Odish F, Gill I, et al. Older age and comorbidity are independent mortality predictors in a large cohort of 1305 COVID-19 patients in Michigan, United States. J Intern Med 2020;288:469–76.

21. Older Adults: At greater risk of requiring hospitalization or dying if diagnosed with COVID-19. 2021. (Accessed April 8, 2021, at https://www.cdc.gov/coronavirus/2019-ncov/need-extra-precautions/older-adults.html#:~:text=CDC&20recommends&20that&20adults&2065,supply&20of&20COVID&2D19&20vaccines.)

22. Lang FM, Lee KMC, Teijaro JR, Becher B, Hamilton JA. GM-CSF-based treatments in COVID-19: reconciling opposing therapeutic approaches. Nat Rev Immunol 2020;20:507–14.

23. Mehta P, Porter JC, Manson JJ, et al. Therapeutic blockade of granulocyte macrophage colony-stimulating factor in COVID-19-associated hyperinflammation: challenges and opportunities. Lancet Respir Med 2020;8:822–30.

24. Thwaites RS, Sanchez Sevilla Uruchurtu A, Siggins MK, et al. Inflammatory profiles across the spectrum of disease reveal a distinct role for GM-CSF in severe COVID-19. Sci Immunol 2021;6:eabg9873.

25. Bonaventura A, Vecchié A, Wang TS, et al. Targeting GM-CSF in COVID-19 Pneumonia: Rationale and Strategies. Frontiers in Immunology 2020;11.

26. Buckley CD, Simón-Campos JA, Zhdan V, et al. Efficacy, patient-reported outcomes, and safety of the anti-granulocyte macrophage colony-stimulating factor antibody otilimab (GSK3196165) in patients with rheumatoid arthritis: a randomised, phase 2b, dose-ranging study. The Lancet Rheumatology 2020;2:e677–e88.

27. R&D Blueprint novel Coronavirus COVID-19 Therapeutic Trial Synopsis. 2020. (Accessed March 22, 2021, at https://www.who.int/blueprint/priority-diseases/key-action/COVID-19_Treatment_Trial_Design_Master_Protocol_synopsis_Final_18022020.pdf.)

28. Lan KKG, DeMets DL. Discrete Sequential Boundaries for Clinical Trials. Biometrika 1983;70:659–63.

29. Hazeldine J, Lord JM. Immunesenescence: A Predisposing Risk Factor for the Development of COVID-19? Front Immunol 2020;11:573662.

30. Pietrobon AJ, Teixeira FME, Sato MN. Immunosenescence and Inflammaging: Risk Factors of Severe COVID-19 in Older People. Front Immunol 2020;11:579220.

31. Schulte-Schrepping J, Reusch N, Paclik D, et al. Severe COVID-19 Is Marked by a Dysregulated Myeloid Cell Compartment. Cell 2020;182:1419-40.e23.

32. Szabo PA, Dogra P, Gray JI, et al. Longitudinal profiling of respiratory and systemic immune responses reveals myeloid cell-driven lung inflammation in severe COVID-19. Immunity 2021:DOI: 10.1016/j.immuni.2021.03.005.

33. Merad M, Martin JC. Pathological inflammation in patients with COVID-19: a key role for monocytes and macrophages. Nat Rev Immunol 2020;20:355–62.

34. Dorward DA, Russell CD, Um IH, et al. Tissue-Specific Immunopathology in Fatal COVID-19. Am J Respir Crit Care Med 2021;203:192–201.

35. Giamarellos-Bourboulis EJ, Netea MG, Rovina N, et al. Complex Immune Dysregulation in COVID-19 Patients with Severe Respiratory Failure. Cell host & microbe 2020;27:992-1000.e3.

36. Mann ER, Menon M, Knight SB, et al. Longitudinal immune profiling reveals key myeloid signatures associated with COVID-19. Science Immunology 2020;5:eabd6197.

37. Permutt T, Li F. Trimmed means for symptom trials with dropouts. Pharm Stat 2017;16:20–8.

