## Supplement for "A Randomized Trial of Otilimab in Severe COVID-19 Pneumonia (OSCAR)"

**Supplementary Appendix**

This appendix has been provided by the authors to give readers additional information about their work.

Appendix 1

#### List of investigators and hospitals

Argentina

Sanatorio de La Trinidad Mitre, Buenos Aires, Argentina. Javier Altclas, M.D., Mariana Patricia Berte, Adriana Victoria Diaz Balocchi, Maria Verónica Latini, Daniela Malano Barletta, Claudia Cristina Salgueira, M.D., Analia Rosana Santa Maria.

Sanatorio Mayo Privado Sanatoria Allende S.A, Cordoba, Argentina. German Ambasch, M.D., Jorge Luis Tambini Diaz.

Clinica Montegrande-Clinica Privada Monte Grande, Buenos Aires, Argentina. Pedro Xavier Bocca Ruiz, M.D., Xavier Antonio Bocca Pereira, Sebastian Caravaggio, Victor Hugo Pecci, Feliciano Petro Moreno.

Clinca Privada Colombo, Cordoba, Argentina. Hugo Raul Colombo, M.D., Ph.D., Marianela Colombo.

Clínica Privada Independencia de Munro, Buenos Aires, Argentina. Pablo Alexis Christian Doreski, M.D., M.B.A., Jorge Oscar Fandi, Celia Sara Giler, Fernando Ricardo Racca Velasquez, Damian Gonzalo Rutolo, Oscar Alberto Salva.

Sanatorio Allende S.A., Córdoba, Argentina. Fernando Oscar Riera, M.D., Aldana Mano,
Gabriela Virginia Peukert, Carlos Federico Romero.

Belgium

CHU Dinant Godinne, Cliniques Universitaires Catholic University of Louvain Namur, Yvoir, Belgium. Nathalie Ausselet, M.D., Pierre Bulpa, M.D., Benedicte M.J.B. Delaere, M.D., Alain M. Dive, M.D., Patrick Evrard, M.D., Geoffrey Horlait, M.D., Isabelle Michaux, M.D., Pascal Reper, M.D.

UZ Brussel, Brussels, Belgium. Elisabeth De Waele, M.D., Ph.D., Marc W. Diltoer, M.D., Ph.D., Joop Jonckheer, M.D., Michael Mekeirele, M.D., Duc Nam Nguyen, M.D., Ph.D., Matthias Raes, M.D., Simon de Ridder, M.D., Domien Vanhonacker, M.D.

Brazil

Hospital das Clinicas da Faculdade de Medicina da Universidade de São Paulo, São Paulo, Brazil. Esper Georges Kallas, M.D., PhD., Angela Carvalho Freitas, Gabriel Fialkovitz da Costa Leite, Lucas Chaves Netto.

Universidade Federal de Minas Gerais, Minas Gerais, Brazil. Jorge Andrade Pinto, M.D., Thiago Bragança Lana Silveira Ataide, Renan Detoffol Bragança, Helena Duani, Flavia Gomes Faleiro Ferreira.

Hospital Alemão Oswaldo Cruz, Hospital Sirio Libanes Instituto de Ciências, São Paulo, Brazil. Victor Augusto Hamamoto Sato, M.D., Daniela Ghidetti Mangas Catarino, Jose Victor Gomes Costa, Erico Souza De Oliveira, M.D., Sara Mohrbacher, M.D., Leonardo Victor Barbosa Pereira, M.D., Ph.D.

Hospital de Clínicas de Porto Alegre, Porto Alegre, Brazil. Eduardo Sprinz, M.D., Ph.D., M.Sc., Gustavo Leal Agune, Beatriz Arns, Ana Elize Barin, Murillo Machado Cipolat, Julio Cezar Gonçalves Cordeiro dos Santos, Leonardo Martins Pires, Guilherme Geraldo Lovato Sorio.

Canada

Hôpital du Sacré Cœur de Montréal, Montréal, Québec, Canada. Martin Albert, M.D., Francis Bernard, Yiorgos Alexandros Cavayas, M.D., Karim Serri, David Williamson.

UBC James Hogg Research Centre Saint Paul's Hospital, Vancouver, British Columbia, Canada. John Boyd, M.D., Adam Peets, Demetrios Sirounis.

CISSS de la Montérégie-Centre/Hôpital Charles Le Moyne, Taschereau, Québec, Canada. Germain Poirier, M.D., C.M., F.R.C.P.C., Antoine Delage, Louise Passerini.

Unite de recherche Clinique du CISSS des Laurentides, Saint-Jerome, Québec, Canada. Sebastien Poulin, M.D., Louay Mardini, M.D., Yves Pesant, M.D.

Chile

Clinica Las Condes, Las Condes, Santiago, Chile. Tomas Regueira Heskia, M.D., Ph.D., Andrés Alberto Ferre Contreras, M.D.

France

HCL – Centre Hospitalier Lyon Sud, Pierre-Bénite, France. Bernard Allaouchiche, M.D., Ph.D., Marie Darien, M.D., Donatien De Seissan De Marignan, M.D., Marion Didier, M.D., Emilie Joffredo, M.D., Maxime Lecocq, M.D., Alain Lepape, M.D., Mélanie Levrard, M.D., Florent Wallet, M.D.

CHU d'Angers, Angers, France. Pierre Asfar, M.D., Ph.D., François Beloncle, M.D., Julien Demiselle, M.D., Hélène Julien, M.D., Achille Kouatchet, M.D., Satar Mortaza, M.D., Pierre-Yves Olivier, M.D., Marc Pierrot, M.D., Vincent Souday, M.D.

CH Valenciennes - Hôpital Jean Bernard, Valenciennes, France. Hatem Boughanmi, M.D., Fabien Lambiotte, M.D., Piehr Saint Leger, M.D.

GH Pitié-Salpêtrière, Paris, France. Jean-Michel Constantin, M.D., Ph.D., Mona Assefi, M.D., Marine Le Corre, M.D., Bao-Long Nguyen, M.D., Cyril Quemeneur, M.D., Guillaume Savary, M.D., Agathe Selves, M.D.

Centre Hospitalier Universitaire de Limoges, Limoges, France. Bruno François, M.D., Thomas Daix, M.D., Arnaud Desachy, M.D., Bruno Evrard, M.D., Anne-Laure Fedou, M.D., Guillaume Gilbert, M.D., Marine Goudelin, M.D., Amandine Sanson, M.D., Julien Vaidie, M.D., Philippe Vignon, M.D., Ph.D.

CHD Vendée – Site De La Roche-sur-Yon, La Roche-Sur-Yon, France. Jean-Claude Lacherade, M.D., Marie-Ange Azais, M.D., Konstantinos Bachoumas, M.D., Rémi Bernardon, M.D., Gauthier Blonz, M.D., Gwenhaël Colin, M.D., Matthieu Henry-Lagarrigue, M.D., Christine Lebert, M.D., Laurent Martin-Lefevre, M.D., Caroline Pouplet, M.D., Isabelle Vinatier, M.D., Aihem Yehia, M.D.

CHU Amiens-Picardie - Site Sud, Amiens, France. Julien Maizel, M.D., Ph.D., Abdallah Al Salameh, M.D., Claire Andrejak, M.D., Damien Basille, M.D., Clément Brault, M.D., Mélanie Drucbert, M.D., Mathieu Guilbart, M.D., Jean-Philippe Lanoix, M.D., Michel Slama, M.D., Sandrine Soriot-Thomas, M.D., Yoann Zerbib, M.D.

Hôpitaux Universitaires de Strasbourg - Nouvel Hôpital Civil, Strasbourg, France. Ferhat Meziani, M.D., Ph.D., Jessy Cattelan, M.D., Raphaël Clere-Jehl, M.D., Julie Helms, M.D., Christine Kummerlen, M.D., Hamid Merdji, M.D., Alexandra Monnier, M.D., Hassene Rahmani, M.D., Antoine Studer, M.D.

Groupe Hospitalier Sud Ile de France, Melun, France. Mehran Monchi, M.D., Esther Mbakulu Muanda, M.D., Franck Pourcine, M.D.

Centre Hospitalier Victor Dupouy, Argenteuil, France. Gaëtan Plantefeve, M.D., Damien Contou, M.D., Megan Fraisse, M.D., Elsa Logre, M.D.

Hôpitaux Universitaires de Strasbourg - Hôpital de Hautepierre, Service de Médecine Intensive et Réanimation, Strasbourg, France. Francis Schneider, M.D., Ph.D., Mathieu Baldacini, M.D., Thien Nga Chamaraux, M.D., Pierre Diemunsch, M.D., Sophie Diemunsch, M.D., Jean-Etienne Herbrecht, M.D., Pierre-Olivier Ludes, M.D., Guillaume Morel, M.D., Eric Noll, M.D., Julien Pottecher, M.D.

CHU Grenoble Alpes – Site Nord La Tronche - Hôpital Michallon, La Tronche, France. Nicolas Terzi, M.D., Anaïs Dartevel, M.D., Louis-Marie Galerneau, M.D., Côme Gerard, M.D., Guillaume Rigault, M.D., Carole Schwebel, M.D., Florian Sigaud, M.D.

India

Max Smart super Speciality Hospital (A Unit of Gujarmal Modi Hospital and Research center for Medical Science), New Delhi, India. Ritesh Aggarwal, M.B.B.S., D.N.B., Arun Dewan.

Medica Superspecialty Hospital, Kolkata, India. Tanmay Banerjee, M.D., M.B.B.S., Anirban Bose, M.D., Amitabha Saha, M.D.

Government Medical College, Aurangabad, India. Meenakshi Bhattacharya, M.D., Aditya Gudhate, Avinash Humbe.

Saint Theresas Hospital, Hyderabad, India. Nagaraju Boyilla, M.D., M.B.B.S., Akula Venkateshwar Rao.

Government Medical College and Hospital Nagpur, Nagpur, India. Dipti Chand, M.D., M.B.B.S., Nikhil Agarwal, Chandrashekhar Atkar, Sagar Khandare.

Peerless Hospitex Hospital and research center limited, Kolkata, India. Rimita Dey, M.D., M.B.B.S., Subhrojyoti Bhowmik, M.D.

Kasturba Hospital for Infectious Disease, Mumbai, India. Ajay Jhaveri, M.B.B.S., D.N.B., Chandrakant Pandurang Pawar, M.D.

Noble Hospital Private Limited, Pune, India. Reema Kashiva, M.D., M.B.B.S., Prashant Raghunath Potdar.

MGM Medical College and Hospital, Aurangabad, India. Umar Quadri Syed, M.D., M.B.B.S., Rohit Jacob, Nilofer Bano Patel.

Ruby Hall Clinic, Cancer Centre, Pune, India. Kapil Zirpe, M.D., Abhijit Deshmukh.

Japan

Shonan Fujisawa Tokushukai Hospital, Kanagawa, Japan. Makoto Hibino, M.D., Shigeto Horiuchi, Tetsuri Kondo, Shunichi Tobe.

National Center for Global Health and Medicine, Tokyo, Japan. Shinyu Izumi, Ph.D., M.D., Kazuo Hakkaku, Masao Hashimoto, Masayuki Hojo, Motoyasu Iikura, Takashi Katsuno, Kazuki Kawajiri, Yusaku Kusaba, Ayako Mikami, Momoko Morishita, Chie Morita, Go Naka, Susumu Saito, Keita Sakamoto, Yuriko Sugiura, Manabu Suzuki, Jin Takasaki, Hiroshi Takumida, Yoshie Tsujimoto, Akinari Tsukada, Hiromu Watanabe, Yoh Yamaguchi.

Japanese Red Cross Medical Center, Tokyo, Japan. Takehiro Izumo, Ph.D., M.D., Nobuyasu Awano, Kazushi Fujimoto, Munehiro Hayashi, Minoru Inomata, Yuji Kondo, Naoyuki Kuse, Yuta Moroe, Shun Muramatsu, Yutaka Muto, Akira Nomi, Shogo Sagisaka, Kohei Takada, Mari Tone, Tomoyuki Yamashita.

Yokohama City Minato Red Cross Hospital, Kanagawa, Japan. Isao Nagata, M.D., Michiko Fujisawa, Eisaku Nashiki, Kei Sugiki, Taketo Suzuki, Tetsuhiro Takei, Hiroyuki Yamada, Naoki Yonezawa.

Tokyo Medical and Dental University, Tokyo, Japan. Yasuhiro Otomo, M.D., Akira Endo, Takahiro Mitsumura, Yasunari Miyazaki, Kanae Ochiai, Wataru Takayama.

Tokyo Shinjuku Medical Center, Tokyo, Japan. Hidefumi Shimizu, M.D., Yoshimasa Horie, Hiroshi Kojima, Akira Mizoo.

Osaka City General Hospital, Osaka, Japan. Michinori Shirano, Ph.D., M.D., Tomohiro Asaoka, Tetsushi Goto, Ko Iida, Keiji Konishi, Hidenori Nakagawa.

Saitama Medical University Hospital, Saitama, Japan. Norihito Tarumoto, M.D., Kazuo Imai, Noriomi Ishibashi, Shigefumi Maesaki, Jun Sakai.

Mexico

Fundacion Santos y de la Garza Evia I.B.P. Hospital San José TecSalud, Monterrey, Mexico. José-Fernando Castilleja-Leal, M.D., Daniel Davila-Gonzalez, Alberto Garcia-Vega, Arturo Adrian Martinez-Ibarra, Juan Francisco Moreno-Hoyos Abril.

Hospital Civil Fray Antonio Alcalde, Guadalajara, Mexico. Pedro Martinez-Ayala, M.D., Sara-Alejandra Aguirre-Diaz.

Instituto Nacional de Enfermedades Respiratorias “Ismael Cosío Villegas”, Mexico City, Mexico. Amy-Bethel Peralta-Prado, M.D., Victor Hugo Ahumada-Topete, Maria Isabel Leon-Rodriguez.

Instituto Nacional de Ciencias Medicas y Nutricion Salvador Zubiran, Mexico City, Mexico. Juan Gerardo Sierra-Madero, M.D., Claudia Paola Alarcon-Murra, Jose Guillermo Dominguez-Cherit, Karen-Aranza Marañon-Solorio, Bernardo Alfonso Martínez-Guerra, Carlos Torruco-Sotelo.

Netherlands

Medisch Spectrum Twente, Enschede, Netherlands. Albertus (Bert) Beishuizen, M.D., Ph.D., Alexander D. Cornet, M.D., Ph.D., Bob Oude Velthuis, M.D., Jan W. Vermeijden, M.D., Ph.D.

Canisius Wilhelmina Ziekenhuis, Nijmegen, Netherlands. Oscar Hoiting, M.D., Marco A.A. Peters, M.D., Els Rengers, M.D.

Jeroen Bosch Ziekenhuis, Den Bosch, Netherlands. Frans W. (Wim) Rozendaal, M.D., Cornelis P.C. de. Jager, M.D., Ph.D., Miriam A.M. Moviat, M.D., Ph.D., Anne J. Paling, M.D., Florens N. Polderman, M.D., G.A.M. (Astrid) Salet, M.D., Koen S. Simons, M.D., Ph.D.

Amphia Ziekenhuis, Breda, Netherlands. Simone van der Sar, M.D., Remco S. Djamin, M.D., Ph.D., Klaas M. (Merijn) Kant, M.D., Kornelis H. van der Leest, M.D., Ph.D., Sander Talman, M.D.

Ikazia Ziekenhuis, Rotterdam, Netherlands. Fenna J. Schoonderbeek, M.D., Ph.D., Maaike Muller-Ekas, M.D., Ralph V. Pruijsten, M.D., Anna F.C. Schut-Houtgraaf, M.D., Ph.D., Susanne Stads, M.D., Walter van den Tempel, M.D., Henriette F.E.M. Willems, M.D.

Peru

Clínica Internacional - Seda Lima, Lima, Peru. José Luis Cabrera Rivero, Alfredo Gilberto Guerreros Benavides, Eneyda Giuvanela Llerena Zegarra, Lorena Mata Juarez, Teresa Jhovina Perez Rodriguez, Karla Ysabel Sánchez Vallejos, M.D., Hernando Torres Zevallos, M.D.

Hospital Nacional Alberto Sabogal Sologuren, Lima, Peru. Luis Enrique Hercilla Vásquez, M.D., Erika Cecilia Agurto Lescano, Ysabel Marlene Chavez Santillan, Ronald Nilton Guzman Ramos, Freddy Roberto Marchand Gago, Fernando Franz Namuche Ojeda, D'yanira Felieva Rojas Montaño, Miguel Angel Tapia Paredes, Nora Isabel Villoslada Contreras.

Hospital Nacional Edgardo Rebagliati Martins, Lima, Peru. Fernando Cruz Mendo Urbina, M.D., Oscar Melitón Reyna Vargas, Jose Carlos Ruelas Figueroa.

Poland

Wojewodzki Szpital Obserwacyjno-Zakazny, Bydgoszcz, Poland. Dorota Anna Dybowska, M.D., Ph.D., Dorota Kozielewicz, Przemyslaw Slomkowski.

Centralny Szpital Kliniczny MSWiA w Warszawie, Warszawa, Poland. Andrzej M. Fal, M.D., Ph.D., Kamil Adamczyk, Malgorzata Dorobek, Maciej Jankowski, Anna Jasinska, Antoni Okninski, Iwona Pikto-Pietkiewicz, Katarzyna Przybylowska, M.D., Monika Maria Swiderska, Ewa Szczesniak, Rafal Wojtowicz, Justyna Zielinska-Turek.

Wielospecjalistyczny Szpital Miejski im. J. Strusia, City Hospital of Poznan, Poznan, Poland. Blazej Rozplochowski, M.D., Ph.D., Teresa Ganowicz-Kaatz, Jakub Kopaczynski, Sylwia Michalik, Mozer- Iwona Lisewska, Maja Szczerbakowska.

Szpital Uniwersytecki w Krakowie, Oddzial Kliniczny Chorob Zakaznych, Krakow, Poland. Wojciech Serednicki, M.D., Ph.D., Tomasz Gazda, Teresa Kowal, Anna Kwinta, Rafal Swistek, Wojciech Szpunar.

Wojewodzki Szpital Specjalistyczny im. J. Gromkowskiego, Oddzial Chorob Zakaznych ul, Wroclaw, Poland. Krzysztof Simon, M.D., Ph.D., Justyna Janocha, Anna Szymanek-Pasternak, Aleksander Zinczuk.

Russian Federation

Clinic of Bashkir State Medical University, Ufa, Russian Federation. Bulat Bakirov, M.D., Ph.D., Ruslan Maier.

State Budget Healthcare Institution City Clinical Hospital #15 n.a. O.M.Filatov, Moscow, Russian Federation. Ivan G. Gordeev, M.D., Ph.D., Vitaliy Firstov, Vartan Grigoryan, Ilia Kokorin, Ksenia Komissarova, Anna Kozlova, Nina Lapochkina, M.D., Sevinch Mamedguseyinova, M.D., Ksenia Polubatonova, Natalia Alexandrovna Suvorova, M.D.

Budgetary Healthcare Institution of the Voronezh region ‘Voronezh Regional Clinical Hospital #1’, Voronezh, Russian Federation. Natalia E. Kostina, M.D., Ph.D., Svetlana Chuprina, Olga M. Korolkova, M.D., Ph.D., Andrey Vorobyev, M.D.

BUZ of Omsk Region “Regional Clinical Hospital”, Omsk, Russian Federation. Tatiana Kropotina, M.D., Julia Arbuzova, M.D., Elena Elokhina, M.D., Ekaterina Shelyagina, M.D., Boris Statsenko, M.D.

Nizhniy Novgorod City Clinical Hospital # 10, Nizhniy Novgorod, Russian Federation. Ekaterina V. Makarova, M.D., Ph.D., Ludmila Alexandrovna Lomakina, M.D., Natalya Lyubavina, Vladimir Vakhlamov.

Saint Petersburg State Healthcare Institution City Mariinsky Hospital, Saint Petersburg, Russian Federation. Aleksey G. Mamonov, M.D., Vladimir Okhritckii.

Perm Regional Clinical Hospital #1, Perm, Russian Federation. Yuliya Trefilova, M.D., Ph.D., Galina Bykova, Natalia Grigoriadi, Natalia Komarovskaya, Vera Vustina, Tatiana Yakovleva, Yulia Zhelnina.

City Clinical Hospital #14, Ekaterinburg, Russian Federation. Elena Mikhailovna Vishneva, M.D., Ph.D., Ekaterina A. Egorova, M.D., Anna Pavlovna Isakova.

South Africa

University of Stellenbosch, Cape Town, South Africa. Coenraad F.N. Koegelenberg, Ph.D., M.B.Ch.B., M.Med., Brian William Allwood, M.B.Ch.B., Ph.D., Usha Lalla, M.B.Ch.B.

Durban International Clinical research site, Enhancing Care Foundation, Durban, South Africa. Rosie Mngqibisa, M.B.Ch.B., Umesh Gangharam Lalloo, M.B.Ch.B., Sajeeda Mawlana, M.B.Ch.B., Sundrapragasen Pillay, M.B.Ch.B.

Panorama Research, Cape Town, South Africa. Johan Theron, M.B.Ch.B., Daphne Charlotte Richter, M.B.Ch.B., Morne Johan Vorster, M.B.Ch.B.

Clinical Projects Research SA pty Ltd, Worcester, South Africa. Louis J. van Zyl, M.B.Ch.B., M.Med., Marina Naude, M.B.Ch.B., Christo van Dyk, M.B.Ch.B., M.Med., Francois H. van Zyl, M.B.Ch.B.

Life The Glynnwood Hospital, Eastrand Physicians, Benoni, South Africa. Agatha C. Wilhase, M.B.Ch.B., M.C.F.P., M.Fam.Med., Reece Boosi, M.B.Ch.B., Nabeela Kajee, M.B.Ch.B., M.Med., Jayseelan Naidu, M.B.Ch.B., M.Med., Kuven Naidu, M.B.Ch.B., Bilaal Wadee, M.B.Ch.B.

Spain

Hospital Universitario La Princesa, Madrid, Spain. Tamara Alonso Pérez, M.D., María de Churruca Arróspide.

Hospital Universitario Gregorio Marañón, Madrid, Spain. Javier de Miguel Díez, M.D., Ph.D., Walther Iván Girón Matute, Zichen Ji, Luis Puente Maestu, José Rafael Terán Tinedo.

Hospital del Mar, Barcelona, Spain. Rosana Muñoz Bermúdez, M.D., Judith Marín Corral, Francisco José Parrilla Gómez.

Fundación Jiménez Díaz, Madrid, Spain. Germán Peces-Barba Romero, M.D., Ph.D., Itziar Fernández Ormaechea, Carolina María Gotera Rivera, Pablo López Yeste, Marcel José Rodríguez Guzmán, Arnoldo de Jesús Santos Oviedo.

Hospital de San Pedro, Logroño, Spain. Carlos Ruíz Martínez, M.D., Ph.D., Jorge Alba Fernández, Adolfo Calvo Martínez, Dolores del Puerto García, José Andrés Molina Espejo, Alejandra Roncero Lázaro, Esther Saiz Rodrigo, Javier Ugedo Urruela.

Hospital Universitario de Bellvitge, Barcelona, Spain. Salud Santos Pérez, M.D., Ph.D., Ester Cuevas Sales, Mercè Gasa Galmes, Yolanda Ruiz Albert, Guillermo Rafael Suárez Cuartín, Pere Trias Sabrià.

Hospital Universitario Infanta Sofia, Madrid, Spain. Llanos Soler Rangel, M.D., Ph.D., Miguel Ángel González Gallego, María Teresa Ramírez Prieto, Pilar Ruiz Seco, Inés María Suárez García, Miguel Ángel Vázquez Ronda.

United Kingdom

Royal Victoria Infirmary, Newcastle Upon Tyne, United Kingdom. John Isaacs, Ph.D., F.R.C.P., B.Sc.(Hon.), M.B.B.S., Andrew Barr, Ian Clement, Iain McCullagh, David Ashley Price, Amada Sanchez Gonzalez.

Wythenshawe Hospital, Manchester, United Kingdom. Andrew David Martin, M.B.Ch.B., Peter David Gregory Alexander, Andrew Mark Bentley, Tim Felton, Tolga Turgut.

Manchester University NHS Foundation Trust, Manchester Royal Infirmary, Wythenshawe Hospital, Manchester, United Kingdom. Andrew Martin, M.B.Ch.B., M.R.C.P., F.R.C.A., F.F.I.C.M., C.C.T., Jonathan Bannard-Smith, Steve Benington, Patrick Hamilton, James Hanison, Nina Rendell, Anthony Wilson.

Royal Liverpool University Hospital, Liverpool, United Kingdom. Ingeborg Dorothea Maria Welters, M.D., Ph.D., Oliver Hamilton, Brian Johnston, Suleman Mulla, Alicia Waite, Richard Wenstone.

United States

University of California, Davis Medical Center, Sacramento, California, United States. Timothy Albertson, M.D., Stuart H. Cohen, M.D., Brian M. Morrissey, M.D., Christian Sandrock, M.D.

Baltimore VA Medical Center, Baltimore, Maryland, United States. Jacqueline Bork, M.D., Jennifer Y. So, M.D., Rohit Talwani, M.D.

G.V.(Sonny) Montgomery Veterans Administration Medical Center, Jackson, Mississippi, United States. Mary Jane Burton, M.D., Gwendolyn Adams-McAlpine, N.P., Christian D. Weaver, M.D.

Temple University Hospital, Philadelphia, Pennsylvania, United States. Gerard J. Criner, M.D., James C. Brown, M.D., Junad Mir Chowdhury, M.D., Stephen E. Codella, M.D., Parag B. Desai, M.D., Gustavo Adolfo Fernandez Romero, M.D., Rohit Gupta, M.D., Fredric Jaffe, D.O., Navjot Ariyana Kaur, M.D., Samuel L. Krachman, D.O., Albert James Mamary, M.D., Nathaniel Marchetti, D.O., Janpreet Mokha, M.D., Oisin O'Corragain, M.D., Maulin Patel, M.D., Parth M. Rali, M.D., Ekamjeet Singh Randhawa, M.D., Daniel Alejandro Salerno, M.D., M.S., Kartik V. Shenoy, M.D., Omar Sheriff, M.D., Jeffrey I. Stewart, M.D., Maria Elena Vega-Sanchez, M.D., Steven R. Verga, M.D., Ibraheem Fares Mohammad Yousef, M.D., Matthew Zheng, M.D.

Carilion Roanoke Memorial Hospital Carilion Clinic, Roanoke, Virginia, United States. Dorothy C. Garner, M.D., Ekta N. Bansal, M.D., Mariana Gomez De La Espriella, M.D., Ruth Ndolo, R.N., Fazili Tasaduq, M.D., F.A.C.P., Jim Wong, M.D.

The Lundquist Institute for Biomedical Innovation, Harbor UCLA Medical Center, Torrance, California, United States. Timothy J. Hatlen, M.D., Eric S. Daar, M.D.

Mercy Health- Saint Vincent Medical Center, Toledo, Ohio, United States. Luis E. Jauregui-Peredo, M.D., Aouad, Tanos Arlette, M.D., Salil Avasthi, M.D., Tanyanyiwa W. Chinyadza, M.D., Srinivas Katragadda, M.D., James A. Tita, D.O.

Renown Regional Medical Centre, Reno, Nevada, United States. Farah K. Madhani-Lovely, M.D., Daniel Antwi-Amoabeng, M.D., Rudy Tedja, D.O.

Clement J Zablocki Veterans Administration Medical Center, Milwaukee, Wisconsin, United States. Sheran Mahatme, M.D., Nathan D. Gundacker, M.D.

Northwestern Medicine Central DuPage Hospital, Winfield, Illinois, United States. Luis A. Manrique, M.D., Daniel Boyle, M.D.

Regions Hospital, Saint Paul, Minnesota, United States. Charlene E. McEvoy, M.D. M.P.H, Omobosola O. Akinsete, M.D., Charles A. Bruen, M.D., Kealy Rae Ham, M.D., Sarah Rebecca Peglow, M.D.

Methodist Hospital, St. Louis Park, Minnesota, United States. Charlene E. McEvoy, M.D. M.P.H, Amanda Calvin, M.D., Elizabeth Miller, M.D.

Multicare Institute for Research & Innovation, Tacoma, Washington, United States. Patrick Scott Meehan, Jr., M.D., Vinay Malhotra, M.D.

University of Arkansas for Medical Sciences, Little Rock, Arkansas, United States. Nikhil Kumar Meena, M.D., Jose Diego Caceres, M.D., Harmeen Goraya, M.D., Larry Galdoc Johnson, M.D.

Holy Cross Hospital, Silver Spring, Maryland, United States. Lee Edward Schwab, M.D., Bernice Wiredu Aidoo, M.D., Miriam Louise Cameron, M.D., Ramani Reddy, M.D.

Holy Cross Germantown Hospital, Germantown, Maryland, United States. Lee Edward Schwab, M.D., Miriam Louise Cameron, M.D., Farah Munnir Cheema, M.D., Sameer B. Ismailjee, M.D.

Pulmonary Associates of Mobile, Mobile, Alabama, United States. Allan F. Seibert, IV, M.D., Michele L. Hemphill, R.N., Daniel Joseph Pollman, M.D.

Atrium Health, Charlotte, North Carolina, United States. Zainab Shahid, M.D., Michael Sean Boger, M.D., Yvonne Lynnette Carter, M.D., Tedra Claytor, M.D., Lisa E. Davidson, M.D., Azeem Elahi, M.D., Joseph P. Lang, M.D., Michael K. Leonard, M.D., Lewis Hall McCurdy, M.D., Leigh Ann Medaris, M.D., Tue H. Ngo, M.D., Fred C. Papali, M.D., C.M., Christopher Polk, M.D., Nestor Manuel Quezada, M.D., Mindy M. Sampson, M.D., D.O., Jaspal Singh, M.D., Kranthi K. Sitammagari, M.D., Stephanie L. Strollo, M.D., Brice Thomas Taylor, M.D., Shelley S. Towery, R.N., David Allan Weinrib, M.D., Michael Zgoda, M.D., Sara Zulfigar, M.D.

North Florida South Georgia Veterans Health System, Gainesville, Florida, United States. Peruvemba S. Sriram, M.D.

Doylestown Hospital, Doylestown, Pennsylvania, United States. Les A. Szekely, M.D., Pinak Sumant Acharya, M.D., Manuel Jose Jimenez-Serrano, M.D., Kathy Tran-Gast, D.O.

### Appendix 2

#### Figure S1A. Alive and free of respiratory failure (baseline characteristics, medical history, pre-treatment medications) – mITT population

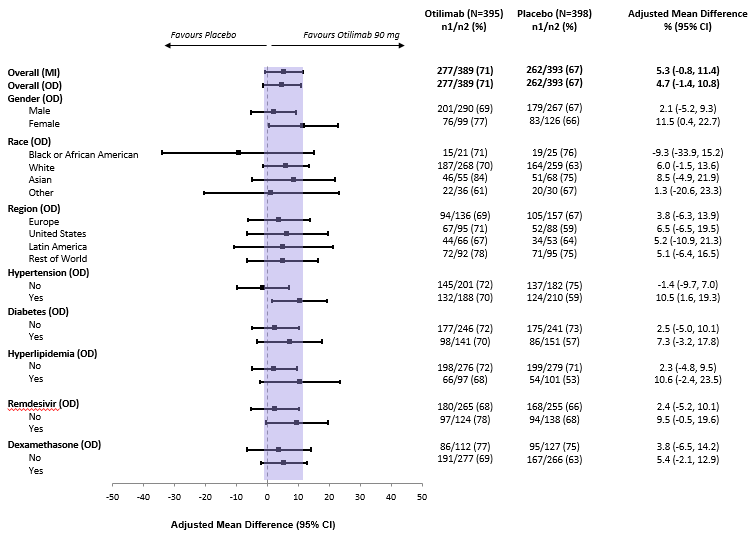

CI, confidence interval; MI, multiple imputation; n1, number of patients with the event; n2, number of patients with non-missing data at timepoint; OD, observed data.

#### Figure S1B. Alive and free of respiratory failure (baseline characteristics, medical history, pre-treatment medications) – mITT ≥70-year group

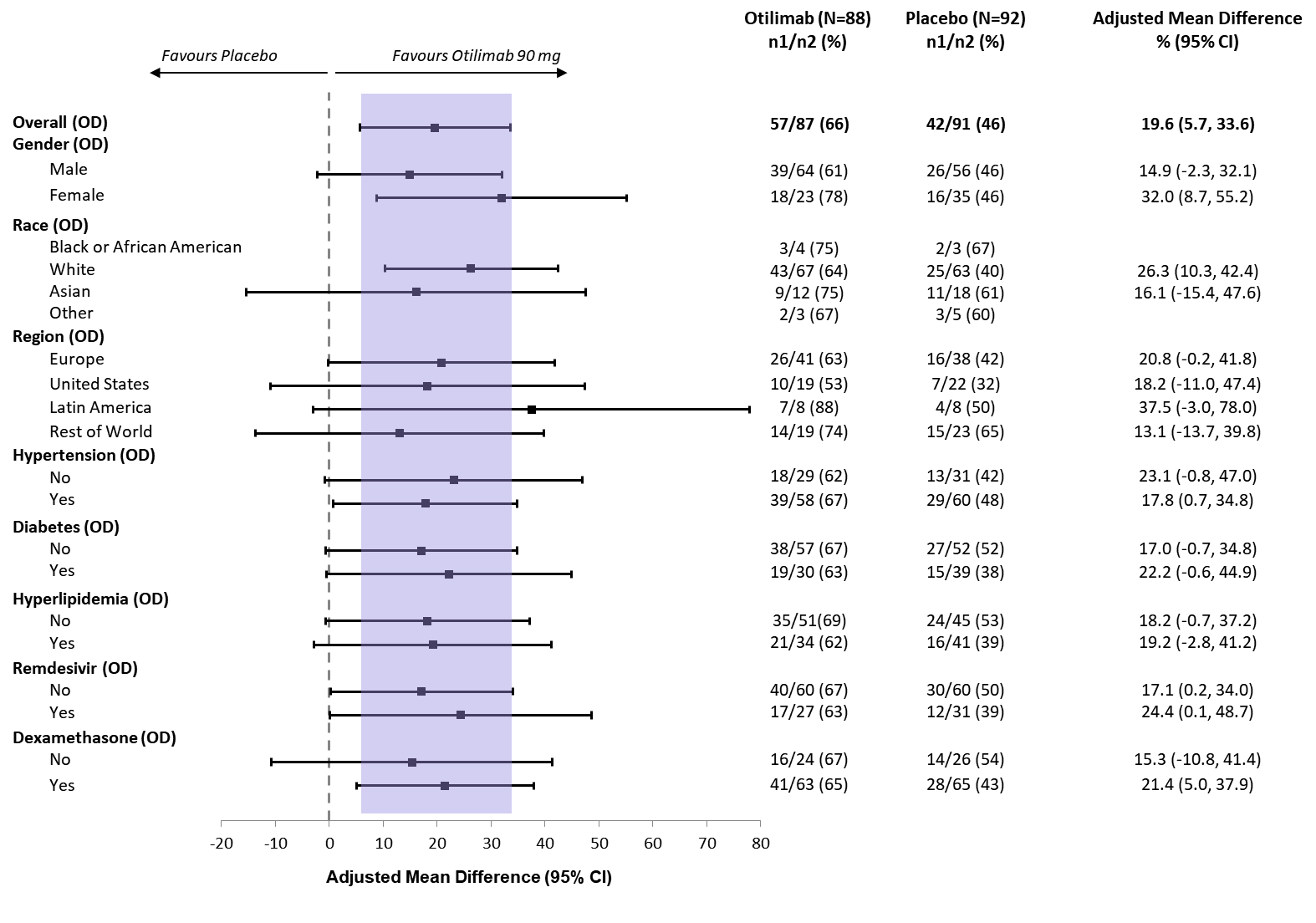

CI, confidence interval; MI, multiple imputation; n1, number of patients with the event; n2, number of patients with non-missing data at timepoint; OD, observed data.

#### Figure S2. Individual pharmacokinetic profiles of otilimab 90 mg in the PK population (A) and ≥70 year group (B)

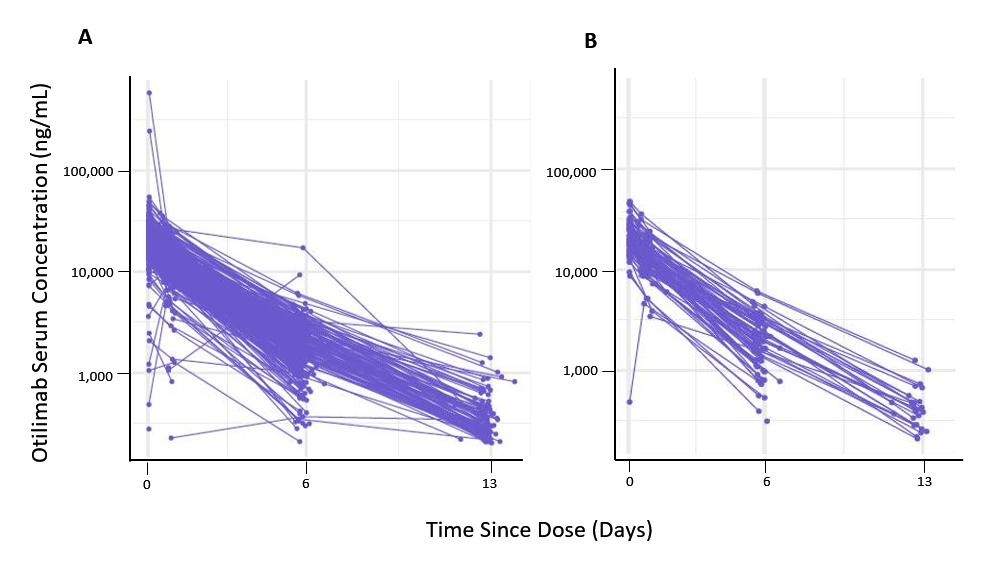
