## Supplementary material for "A Randomized Trial of Otilimab in Severe COVID-19 Pneumonia (OSCAR)": Reporting and analysis plan

| | | |
|---|---|---|
| Division | : | Worldwide Development |
| Information Type | : | Statistical Analysis Plan (SAP) |

### TITLE PAGE

**Protocol Title:** A randomized, double-blind, placebo-controlled, study evaluating the efficacy and safety of otilimab IV in patients with severe pulmonary COVID-19 related disease.

**Protocol Number**: 214094

**Compound Number:** GSK3196165 (otilimab)

**Short Title:** Investigating otilimab in patients with severe pulmonary COVID-19 related disease.

**Sponsor Name**:

GlaxoSmithKline Research & Development Limited
980 Great West Road
Brentford
Middlesex, TW8 9GS
UK

**Regulatory Agency Identifier Number(s)**

| Registry | ID |
|---|---|
| IND | 149754 |
| EudraCT | 2020-001759-42 |

Copyright 2020 the GlaxoSmithKline group of companies. All rights reserved. Unauthorised copying or use of this information is prohibited.

**SAP Author(s):**

| Author |
|---|
| PPD<br>Principal Statistician, Biostatistics |

**SAP Team Review Confirmations (E-mail Confirmation):**

| Approver | Date of E-mail Confirmation |
|---|---|
| PPD<br>Development Scientist | 07-Dec-2020 |
| PPD<br>Physician, Discovery Medicines | 08-Dec-2020 |
| PPD<br>Director, Pharma Safety | 07-Dec-2020 |
| PPD<br>Principal Programmer | 04-Dec-2020 |

**Clinical Statistics Line Approval (eSignature):**

| Approver |
|---|
| PPD<br>Statistics Director, Biostatistics |

5.5. Safety Analyses ... 37
5.5.1. Extent of Exposure ... 37
5.5.2. Adverse Events ... 37
5.5.2.1. Adverse Events of Special Interest ... 39
5.5.3. Additional Safety Assessments ... 39
5.5.3.1. Laboratory Data ... 39
5.5.3.2. Vital Signs ... 43
5.5.3.3. ECG ... 44
5.6. Other Analyses ... 44
5.6.1. Pharmacokinetics (PK) ... 44
5.6.2. Population Pharmacokinetics ... 44
5.6.3. Pharmacodynamics ... 45
5.6.4. Pharmacokinetics/Pharmacodynamics ... 45
5.7. Interim Analyses ... 45

6. SUPPORTING DOCUMENTATION ... 46
6.1. Appendix 1 Abbreviations and Trademarks ... 46
6.1.1. List of Abbreviations ... 46
6.1.2. Trademarks ... 47
6.2. Appendix 2: Changes to Protocol-Planned Analyses ... 48
6.3. Appendix 3: Statistical Modelling ... 49
6.3.1. Multiple Imputation ... 49
6.3.1.1. Adaptive Rounding ... 51
6.3.2. Proportional Odds Regression Model Checking ... 51
6.3.3. Time to Event Model Checking ... 51
6.3.4. Count Data Model Checking ... 51

7. REFERENCES ... 52

### VERSION HISTORY

This Statistical Analysis Plan (SAP) for study 214094 is based on the protocol amendment 2 (GSK Document Number 2020N436091_02) dated 02-JUL-2020.

**Table 1 SAP Version History Summary**

| SAP Version | Approval Date | Change | Rationale |
|---|---|---|---|
| Amendment 3 | Current Version | The key changes include:<br>• Removal of the ~600 participant interim analysis<br>• Inclusion of interim primary endpoint analysis of all participants<br>• Clarification on exploratory endpoints<br>• Other minor, grammatical and typographical corrections to improve readability | Update to the SAP in response to regulatory feedback, removal of the ~600 participant interim analysis, Inclusion of interim primary endpoint analysis of all participants and clarifications. |
| Amendment 2 | 30-Sep-2020 | The key changes include:<br>• Inclusion of additional endpoints related to discharge from investigational site and discharge to non-hospitalized residence<br>• Inclusion of the heart rate potential clinical importance (PCI) ranges<br>• Other minor, grammatical and typographical corrections to improve readability | Update to the SAP in response to regulatory feedback, and clarifications. |
| Amendment 1 | 05-Jun-2020 | The key changes include:<br>• revision of the primary endpoint (where definition of “free of respiratory failure” is participants in categories 1-4 on GSK Ordinal Scale) in line with the protocol amendment<br>• Inclusion of a hierarchy strategy for secondary endpoints<br>• Other minor, grammatical | Update to the SAP following protocol amendment in response to regulatory feedback, and clarifications based on investigator feedback. |

| SAP Version | Approval Date | Change | Rationale |
|---|---|---|---|
| | | and typographical corrections to improve readability | |
| Original SAP | 13-May-2020 | Not Applicable | Original version |

### 1. INTRODUCTION

The purpose of this SAP is to describe the planned analyses to be included in the Clinical Study Report for Study 214094. Details of the planned interim analyses, in addition to the final analyses, are provided.

Additional detail with regards to data handling conventions and the specification of data displays will be provided in the Output and Programming Specification (OPS) document.

#### 1.1. Changes to the Protocol Defined Statistical Analysis Plan

Changes from the originally planned statistical analysis specified in the protocol (GSK Document Number 2020N436091_02) [(Dated: 02- JUL-2020)] are outlined in Table 2.

**Table 2 Changes to Protocol Defined Analysis Plan**

| Protocol | Reporting & Analysis Plan | |
|---|---|---|
| **Statistical Analysis Plan** | **Statistical Analysis Plan** | **Rationale for Changes** |
| The study will utilize a group sequential design, using a Lan-DeMets alpha-spending function to control the type I error with four interim analyses for futility, two early in the study where futility alone will be assessed, using Pocock analogue rules; and two for efficacy, at later times in the study and at the same time as futility, using the O'Brien-Fleming analogue rules [Lan, 1983]. | The study will utilize a group sequential design, using a Lan-DeMets alpha-spending function to control the type I error with three interim analyses for futility, two early in the study where futility alone will be assessed, using Pocock analogue rules; and one for efficacy, at a later time in the study and at the same time as futility, using the O'Brien-Fleming analogue rules [Lan, 1983]. | As recruitment has completed prior to formal interim analyses number 4 (on approximately 600 participants), it was decided to remove the formal analyses on approximately 600 participants and continue to the final analyses only.<br>Note: The final analyses boundary has been adjusted to carry over the alpha that would have been spent on the 600-participant analysis. Hierarchy has been updated accordingly but previous boundaries were not amended. |
| N/A | For the primary endpoint, clarification has been added to detail that the final analyses will be conducted once all participants have been randomised and completed to Day 28 or withdrawn. | Given the ongoing pandemic situation and the urgent unmet need for COVID-19 treatments, to ensure this is met as soon as possible final analyses on the primary endpoint (assessed at Day 28) will be conducted once all participants have been randomised and completed to Day 28 or withdrawn. |

| Protocol | Reporting & Analysis Plan | |
|---|---|---|
| **Statistical Analysis Plan** | **Statistical Analysis Plan** | **Rationale for Changes** |
| N/A | Included text on the proposed hierarchy for testing secondary endpoints, refer to Section 2 for full details. | To account for multiplicity a hierarchy has been proposed for testing secondary endpoints. |
| The secondary endpoint: "Time to final hospital discharge up to Day 28" | Updated secondary endpoints to include:<br>• Time to first discharge from investigational site up to Day 60<br>• Time to first discharge to non-hospitalised residence up to Day 60 | Final hospital discharge is not being collected. Participants may be discharged to an intermediator hospital residence hence we cannot assume discharge from the investigator site is equivalent to discharge from hospital.<br>A post go live change was created to capture discharge to a non-hospitalised residence.<br>eCRF guidelines were updated to clarify re-hospitalisation refers to the investigator site. |
| For exploratory endpoints on improvement in SOFA, Blood Oxygen Saturation ($SpO_2$), Concentration of Inspired Oxygen ($FiO_2$) and $SpO_2/FiO_2$ ratio | Clarified exploratory endpoints to include recovery as part of composite strategy, i.e.:<br>• Recovery or Improvement of at Least 2 Points Relative to Baseline of Sequential Organ Failure Assessment (Sofa) Score up to Day 28<br>• Recovery or Improvement Relative to Baseline in Blood Oxygen Saturation (SpO2),<br>• Recovery or Improvement Relative to Baseline in Concentration of Inspired Oxygen (FiO2)<br>• Recovery or Improvement Relative to Baseline in SpO2/FiO2 ratio | Clarified exploratory endpoints to include recovery as part of composite strategy. |

| Protocol | Reporting & Analysis Plan | |
|---|---|---|
| **Statistical Analysis Plan** | **Statistical Analysis Plan** | **Rationale for Changes** |
| For exploratory endpoints on Time to Improvement of at Least 2 Categories Relative to Baseline on an Ordinal Scale up to Day 60 | For exploratory endpoints on Time to Improvement of at Least 2 Categories Relative to Baseline in Clinical Status up to Day 60 | Clarified exploratory endpoints to use clinical status rather than ordinal scale for consistency |

#### 1.2. Objectives, Endpoints and Estimands

##### 1.2.1. Objectives and Endpoints

| Objectives | Endpoints |
|---|---|
| **Primary** | |
| • To compare the efficacy of otilimab 90mg IV versus placebo. | • Participants alive and free of respiratory failure at Day 28. |
| **Secondary** | |
| • To compare the efficacy of otilimab 90mg IV versus placebo. | • All-cause mortality at Day 60<br>• Time to all-cause mortality at Day 60<br>• Participants alive and free of respiratory failure at Day 7, 14, 42, and 60<br>• Time to recovery from respiratory failure up to Day 28<br>• Participants alive and independent of supplementary oxygen at Day 7, 14, 28, 42, and 60<br>• Time to last dependence on supplementary oxygen up to Day 28<br>• Admission to ICU up to Day 28<br>• Time to final ICU discharge up to Day 28<br>• Time to first discharge from investigational site up to Day 60<br>• Time to first discharge to non-hospitalised residence up to Day 60 |
| • To compare the safety and tolerability of otilimab 90mg IV versus placebo. | • Occurrence of adverse events (AEs) [up to Day 60]<br>• Occurrence of serious adverse events (SAEs) [up to Day 60] |

| Objectives | Endpoints |
|---|---|
| **Exploratory** | |
| • To compare the efficacy of otilimab 90mg IV versus placebo. | **Other endpoints up to Day 28**<br>• Invasive mechanical ventilation (if not previously initiated)<br>• Time to invasive mechanical ventilation (if not previously initiated)<br>• Alive and not invasively mechanically-ventilated<br>• Time to definitive extubation<br>• Improvement of at least 2 points relative to baseline of Sequential Organ Failure Assessment (SOFA) score<br>• Improvement relative to baseline in $SpO_2$, $FiO_2$, and $SpO_2/FiO_2$ ratio<br>• Oxygen-free days<br>• Ventilator-free days<br>• Time to resolution of pyrexia (for at least 48h)<br><br>**Other endpoints up to Day 60**<br>• Clinical status assessed using an ordinal scale assessed at<br>• Days 4, 7, 14, 28, 42, and 60<br>• Time to improvement of at least 2 categories relative to baseline on an ordinal scale<br>• Change in COVID-19 signs and symptoms |
| • To determine the pharmacokinetics (PK) profile of otilimab. | **PK Endpoints up to Day 14**<br>Otilimab apparent clearance (CL/F) and other PK parameters as appropriate using sparse PK sampling |
| • To determine:<br>○ Exposure-response.<br>○ Pharmacodynamic (PD) biomarkers.<br>○ Changes in key markers of inflammation | **PD Endpoints up to Day 28**<br>• Exposure-response relationship for key efficacy, safety and PD endpoints.<br>• Key markers of inflammation including, but not limited to CRP, serum ferritin and inflammatory cytokines as appropriate. |

##### 1.2.2. Estimands

Each study objective is presented below with additional information, including prespecified estimands with related attributes.

**Table 3 Estimands**

| Estimand Category | Estimand | | | |
|---|---|---|---|---|
| | **Variable/Endpoint** | **Population** | **Intercurrent Event Strategy** | **Treatment Comparison** |
| **Primary Objective***:* To compare the efficacy of otilimab 90 mg IV versus placebo | | | | |
| Primary | Participants alive and free of respiratory failure at Day 28 | Entire trial population randomised and receiving treatment (MITT) | Data analysed as collected (treatment policy strategy) | Odds ratio and difference in proportions |
| **Secondary Objective:** To compare the efficacy of otilimab 90 mg IV versus placebo. | | | | |
| Secondary 1 | All-cause mortality at Day 60 | Entire trial population randomised and receiving treatment (MITT) | Data analysed as collected (treatment policy strategy) | Odds ratio and difference in proportions |
| Secondary 2 | Time to all-cause mortality up to Day 60 | Entire trial population randomised and receiving treatment (MITT) | Data analysed as collected (treatment policy strategy) | Hazard ratio |
| Secondary 3 | Participants alive and free of respiratory failure at Day 7, 14, 42, and 60 | Entire trial population randomised and receiving treatment (MITT) | Data analysed as collected (treatment policy strategy) | Odds ratio and difference in proportions |
| Secondary 4 | Time to recovery from respiratory failure up to Day 28 | Entire trial population randomised and receiving treatment (MITT) | Participants who die will be censored at end of follow-up (composite strategy)<br><br>For all other intercurrent events, the data will be analysed as collected (treatment policy strategy) | Hazard ratio |

| Estimand Category | Estimand | | | |
|---|---|---|---|---|
| | **Variable/Endpoint** | **Population** | **Intercurrent Event Strategy** | **Treatment Comparison** |
| Secondary 5 | Participants alive and independent of supplementary oxygen at Day 7, 14, 28, 42, and 60 | Entire trial population randomised and receiving treatment (MITT) | Data analysed as collected (treatment policy strategy) | Odds ratio and difference in proportions |
| Secondary 6 | Time to last dependence on supplementary oxygen up to Day 28 | Entire trial population randomised and receiving treatment (MITT) | Participants who die will be censored at end of follow-up (composite strategy)<br><br>For all other intercurrent events, the data will be analysed as collected (treatment policy strategy) | Hazard ratio |
| Secondary 7 | Admission to ICU up to Day 28 | Entire trial population randomised and receiving treatment (MITT) who are not in ICU at time of dosing | Participants who die will be considered to have been admitted to ICU (negative outcome) (composite strategy)<br><br>For all other intercurrent events the data will be analysed as collected (treatment policy strategy) | Odds ratio and difference in proportions |
| Secondary 8 | Time to final ICU discharge up to Day 28 | Entire trial population randomised and receiving treatment (MITT) who are in ICU at time of dosing | Participants who die will be censored at end of follow-up (composite strategy)<br><br>All other intercurrent events the data will be analysed as collected (treatment policy strategy) | Hazard ratio |

| **Estimand Category** | **Estimand** | | | |
|---|---|---|---|---|
| | **Variable/Endpoint** | **Population** | **Intercurrent Event Strategy** | **Treatment Comparison** |
| Secondary 9 | Time to first discharge from investigational site up to Day 60 | Entire trial population randomised and receiving treatment (MITT) | Participants who die will be censored at end of follow-up (composite strategy)<br><br>All other intercurrent events the data will be analysed as collected (treatment policy strategy) | Hazard ratio |
| Secondary 10 | Time to first discharge to non-hospitalised residence up to Day 60 | Entire trial population randomised and receiving treatment (MITT) | Participants who die will be censored at end of follow-up (composite strategy)<br><br>All other intercurrent events the data will be analysed as collected (treatment policy strategy) | Hazard ratio |
| **Secondary Objective:** To compare the safety and tolerability of otilimab 90 mg IV versus placebo | | | | |
| Secondary 11 | Occurrence of adverse events (AEs) [up to Day 60] | Entire trial population receiving treatment (Safety) | Data analysed as collected (treatment policy strategy) | Frequency & Percentage of Participants |
| Secondary 12 | Occurrence of serious adverse events (SAEs) [up to Day 60] | Entire trial population receiving treatment (Safety) | Data analysed as collected (treatment policy strategy) | Frequency & Percentage of Participants |

#### 1.3. Study Design

Overview of Study Design and Key Features

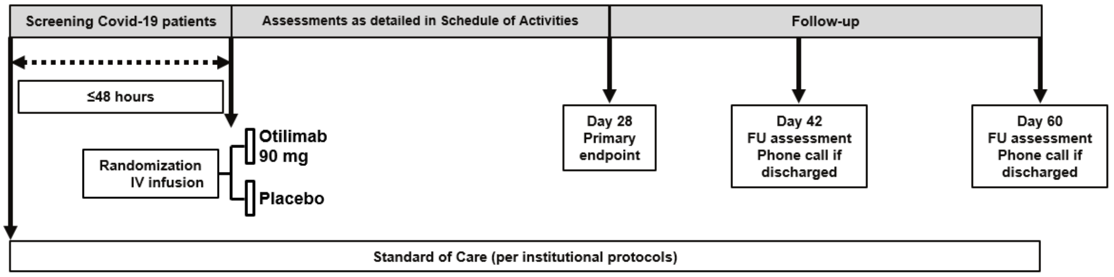

| | |
|---|---|
| Design Features | This study is a multi-center, randomized, double-blind, placebo-controlled trial to assess the efficacy and safety of otilimab for the treatment of severe pulmonary COVID-19 related disease. The study population consists of hospitalized participants with new onset hypoxia requiring significant oxygen support or requiring early invasive mechanical ventilation (≤48 hours before dosing). All participants will receive standard of care as per institutional protocols, in addition to study treatment. |
| Study intervention | Otilimab 90mg or Placebo, single intravenous infusion |
| Study intervention Assignment | Participants will be randomized 1:1 by interactive response technology (IRT) in a blinded manner to receive either a blinded 1-hour infusion of otilimab 90mg or placebo IV in addition to standard of care. Participants will be assessed daily until discharge from investigator site (or Day 28, whichever is sooner), and followed up at Days 42 and 60 after randomization.<br><br>Stratification – based on the study eligibility criteria, the clinical status of the participants entering the main cohort (*i.e.* after the 20th participant) of the study will be within ordinal scale categories [redacted] and also age groups, as shown below.<br>• Hospitalized, high-flow oxygen (≥15L/min), CPAP, BIPAP, non-invasive ventilation. Age <60 years<br>• Hospitalized, high-flow oxygen (≥15L/min), CPAP, BIPAP, non-invasive ventilation. Age 60 to <70 years<br>• Hospitalized, high-flow oxygen (≥15L/min), CPAP, BIPAP, non-invasive ventilation. Age 70 to <80 years<br>• Hospitalized, intubation and mechanical ventilation. Age <60 years<br>• Hospitalized, intubation and mechanical ventilation. Age 60 to <70 years<br>• Hospitalized, intubation and mechanical ventilation. Age 70 to <80 years |
| Interim & Final Analyses | Initial Safety Cohort (first 20 participants)<br>• The first 20 participants will be from (hospitalized, high-flow oxygen ((≥15L/min), CPAP, BIPAP, non-invasive ventilation). Since this is the |

| | |
|---|---|
| | first time that otilimab has been tested in participants with severe pulmonary COVID-19 related disease, initial dosing will be staggered, and safety data reviewed as follows:<br>• The first two participants to be dosed will be randomized 1:1 (otilimab:placebo) and monitored closely. After approximately 24 hours, the clinical status of both participants will be assessed by the investigator(s) and then discussed with the GSK medical monitor. If no safety issues are identified, and the GSK medical monitor and investigator(s) agree, two further participants will be allowed to be dosed.<br>• The third and fourth participants will also be randomized 1:1 (otilimab:placebo) and monitored closely. After approximately 24 hours, the clinical status of all four participants will be assessed by the investigator(s) and then discussed with the GSK medical monitor. If no safety issues are identified, and the GSK medical monitor and investigator(s) agree, recruitment and dosing of further participants will start.<br>• The GSK Safety Review Team (SRT) will review available blinded safety data on an ongoing basis for the additional 16 participants in the initial safety cohort.<br>• Safety data will be reviewed by the Independent Data Monitoring Committee (IDMC), as outlined in the IDMC charter.<br><br>Interim Analyses<br>• Interim analyses will be used to assess futility and efficacy. Full details of timing of analyses and stopping criteria will be given in the IDMC charter.<br>• An IDMC will actively monitor in-stream interim unblinded safety and efficacy data to make recommendations to GSK as per IDMC charter. The IDMC members will include 4-6 physicians with relevant medical specialist training and one statistician. Details regarding the IDMC process will be available in relevant IDMC documents prior to the first participant's visit.<br>• A GSK Safety Review Team (SRT) will review the blinded safety data of this study at regular intervals through both initial safety and main cohort dosing. Details regarding the SRT process will be available in relevant SRT documents. The SRT will inform the IDMC if any safety signals are identified.<br><br>Primary Analyses<br>• The final analyses will be conducted once all participants have been randomised and completed to Day 28 or withdrawn.<br><br>Final Analyses<br>• The final analyses will be conducted once all participants have been randomised and completed the trial or withdrawn; or all participants randomised at the time at which success is declared following an interim analysis complete or withdrawn from the trial whichever occurs first. |

### 2. STATISTICAL HYPOTHESES / SUCCESS CRITERIA

The primary objective of this study is to compare the efficacy of otilimab 90 mg IV versus placebo in participants with severe pulmonary COVID-19 related disease.

The study will test the null hypothesis that the difference between otilimab 90 mg and placebo on the proportion of participants alive and free of respiratory failure at Day 28 is less than or equal to zero versus the alternative hypothesis that otilimab 90 mg increases the proportion of participants alive and free of respiratory failure compared with placebo at Day 28.

#### 2.1. Multiple Comparisons and Multiplicity

The study will utilize a group sequential design, using a Lan-DeMets alpha-spending function to control the type I error with three interim analyses for futility, two early in the study where futility alone will be assessed, using Pocock analogue rules; and one for efficacy, at a later time in the study and at the same time as futility, using the O'Brien-Fleming analogue rules [Lan, 1983].

The testing of secondary endpoints is adjusted for multiplicity by using the following hierarchy:

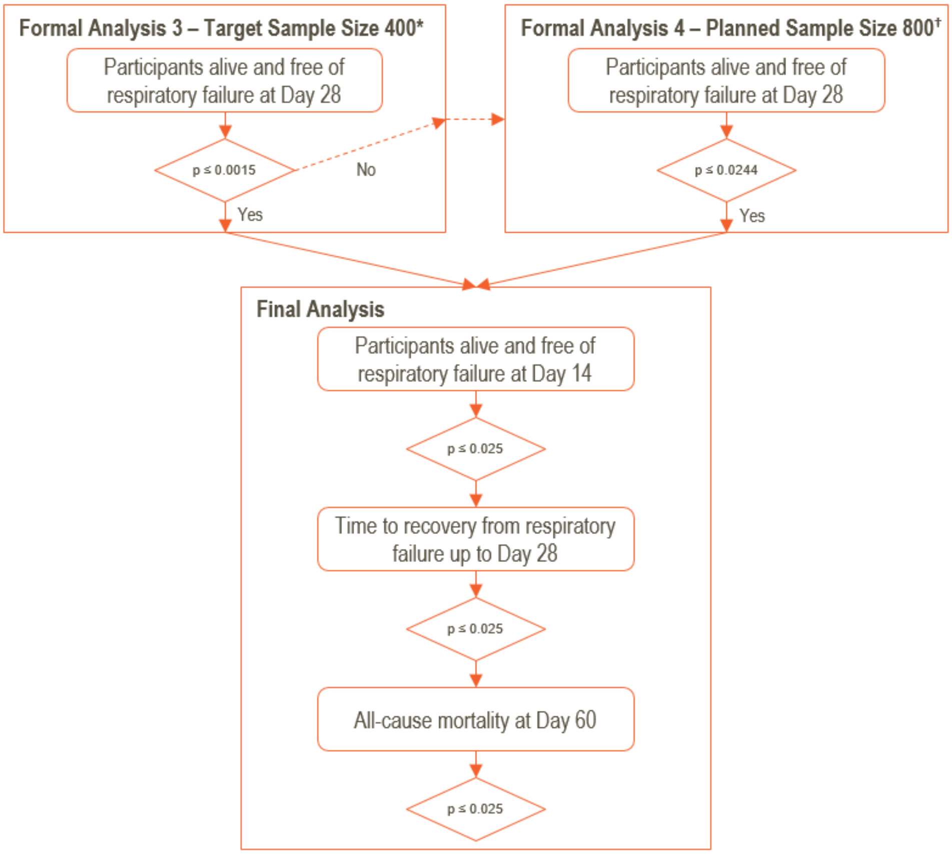

*The boundaries are based on the estimated amount of information at each interim. If the amount included in the interim analysis differs from these values, the actual boundaries will be determined based on the exact amount of information at the time of the interim analyses.

†This boundary is based on the estimated amount of information per the planned sample size of N=800, it will not be updated based on the actual sample size.

Note: The hypothesis on participants alive and free of respiratory failure at Day 14 will only be tested if the null hypothesis on participants alive and free of respiratory failure at Day 28 is rejected (*i.e*. primary endpoint is met). As such the full alpha of 0.025 is transferred down between each endpoint/hypothesis.

### 3. SAMPLE SIZE DETERMINATION

A sample size of 800 participants will provide approximately 90% power to detect a difference of 12% in the proportion of participants alive and free of respiratory failure at a one-sided 2.5% significance level and an assumed placebo response rate of 45%. The minimal detectable effect for this design is 7%.

The assumed placebo response is based on data from recent publications of studies of patients with severe pulmonary COVID-19 related disease [Sanofi, 2020].

### 4. ANALYSIS SETS

| Analysis Set | Definition / Criteria | Analyses Evaluated |
|---|---|---|
| Screened | • All participants who were screened for eligibility | • Study Population |
| Enrolled | • All participants who entered the study.<br>• Data should be reported according to the randomised treatment<br>• Note: screening failures are excluded from the enrolled analysis set as they did not enter the study. | • Study Population |
| Safety | • All participants who received study intervention.<br>• Data should be reported according to the actual treatment received<br>○ If participants receive any Otilimab then they will be summarised according to “Otilimab 90mg”, including interrupted/incomplete infusion. | • Safety<br>• Study Population |
| Modified Intent-To-Treat (MITT) | • All randomized participants who received study intervention<br>• Data should be reported according to the randomised treatment | • Efficacy |
| Pharmacokinetic (PK) | • All participants in the Safety analysis set who had at least 1 non-missing PK assessment (Non-quantifiable [NQ] values will be considered as non-missing values).<br>• Data should be reported according to the actual treatment<br>○ If participants receive any Otilimab then they will be summarised according to “Otilimab 90mg”, including interrupted/incomplete infusion. | • PK |

#### 4.1. Protocol Deviations

Important protocol deviations (including deviations related to study inclusion/exclusion criteria, conduct of the trial, patient management or patient assessment) will be summarised.

Protocol deviations will be tracked by the study team throughout the conduct of the study. These protocol deviations will be reviewed to identify those considered as important as follows:

- Data will be reviewed prior to freezing the database to ensure all important deviations are captured and categorised in the protocol deviations SDTM dataset.
- This dataset will be the basis for the summaries of important protocol deviations.

A separate listing of all inclusion/exclusion criteria deviations will also be provided. This listing will be based on data as recorded on the inclusion/exclusion page of the eCRF.

### 5. STATISTICAL ANALYSES

#### 5.1. General Considerations

##### 5.1.1. General Methodology

The study population analyses will be based on the “Enrolled”, “Screened” or “Safety” populations. The MITT Analysis Set will be used for all Efficacy analyses, and Safety Analysis Set will be used for all safety analyses, unless otherwise specified.

In the case of a difference between the stratification assigned at the time of randomization and the data collected in the eCRF:

- subgroups will be summarised based on the actual subgroup to which the participant belongs

- covariate adjustment will be based on the randomised strata. For participants in the Safety Cohort First Sentinel Pair, Safety Cohort Second Sentinel Pair or Safety Cohort Main the randomised strata will be derived as the calculated stratum at dosing.

All analyses will be adjusted for ordinal scale category at baseline (category 5 or 6 per randomised strata) and age group (treated as categorical variable defined by the levels <60 years, 60 to <70 years, 70 to <80 years per randomised strata). If the number of participants is small within a category of a covariate, then the covariate categories may be refined. If the category cannot be refined further, then the covariate may be included as a continuous measure. Confidence intervals will use the 95% levels unless otherwise specified.

Unless otherwise specified, continuous data will be summarized using descriptive statistics: n, mean, standard deviation, median, minimum and maximum. Categorical data will be summarized as the number and percentage of participants in each category.

Visit windows will be applied to Day 14, Day 28, Day 42 and Day 60, where data within ±2 days of the target day may be used if data is not recorded on the actual day.

##### 5.1.2. Baseline Definition

For all treated participants, for all endpoints and measurements the baseline value will be the latest pre-dose assessment with a non-missing value, including those from unscheduled visits. If time is not collected, Day 1 assessments that were noted to be pre-dose per the Schedule of Activities are assumed to be taken prior to first dose and used as baseline.

For all participants randomised but not dosed, for all endpoints and measurements the baseline value will be the latest assessment on or prior to the date of randomisation with a non-missing value, including those from unscheduled visits.

For ventilation status, baseline ventilation status will be derived as the ventilation at the time of dosing, and if there are multiple ventilation types being used the worst case will be assumed.

Unless otherwise stated, if baseline data is missing no derivation will be performed and baseline will be set to missing.

##### 5.1.3. Multicenter Studies

It is anticipated that patient accrual will be spread thinly across centers and summaries of data by center would unlikely be informative and will not, therefore, be provided.

##### 5.1.4. Intercurrent Events

In general, the following may be considered intercurrent events if not part of the endpoint definition:

- Interruption or infusion stopped early and not completed
- Use of additional medications or changes to standard or care
- Death
- Discharge from ICU
- Admittance to ICU
- Start of Ventilation
- Termination of Ventilation
- Independence from supplementary oxygen
- Discharge from investigator site

The applicable intercurrent events for each endpoint will be highlighted within in the endpoint definition along with the strategy for handling them.

In general, unless otherwise specified, the handling strategy for all these intercurrent events will be based on a treatment policy approach; specifically, the effects estimated will be based on initial randomized treatment arm regardless of whether the participant had experienced an intercurrent event. If possible, data will continue to be collected after the occurrence of the intercurrent event, until the participant either completes the study or withdraws from the study before completion.

##### 5.1.5. Missing Data Handling Rules

Unless otherwise stated the following rules will be applied to all Primary and Secondary endpoints:

- Missing data can occur due to study withdrawal or participants lost to follow-up before the completion of the study or due to intermittent missing values (*i.e.* data between two non-missing assessments).
- Missing data will be imputed under a missing at random (MAR) assumption using a multiple imputation (MI) model, unless otherwise specified. The MI model will include covariates; treatment, ordinal scale category at baseline, age group and baseline of the variable of interest (if appropriate). More details will be provided in Appendix 3.
- For the interim analyses, participant data will only be imputed up to their estimated current point in the trial based on the date of the data cut

##### 5.1.6. Examination of Covariates, Other Strata and Subgroups

###### 5.1.6.1. Covariates and Other Strata

The list of covariates and other strata may be used in descriptive summaries and statistical analyses, some of which may also be used for subgroup analyses. Additional covariates and other strata of clinical interest may also be considered.

- If the number of participants is small within a category of a covariate, then the covariate categories may be refined prior to unblinding the trial.
- If the category cannot be refined further, then the covariate may be included as a continuous measure.

| Category | Details |
|---|---|
| Strata | Clinical status at baseline, age group (<60 years, 60 to <70 years, 70 to <80 years) |
| Covariates | Treatment, Clinical Status at Baseline, Age Group at Baseline, and baseline of the variable of interest (if appropriate). |

###### 5.1.6.2. Examination of Subgroups

The list of subgroups may be used in descriptive summaries and statistical analyses. Additional subgroups of clinical interest may also be considered.

- If there are less than 10 participants within a subgroup, then the subgroup categories may be refined prior to unblinding the trial.
- If the category cannot be refined further, then descriptive rather than statistical comparisons may be performed for the particular subgroup.

| Subgroup | Categories |
|---|---|
| Age Group | 1. <60 years<br>2. 60 to <70 years<br>3. >=70 years |
| Clinical status at baseline | 1. Group 5 [redacted]<br>2. Group 6 [redacted] |
| Strata at baseline | 1. Clinical Status 5 [redacted] and age <60 years<br>2. Clinical Status 5 [redacted] and age 60 to <70 years<br>3. Clinical Status 5 [redacted] and age >=70 years<br>4. Clinical Status 6 [redacted] and age <60 years<br>5. Clinical Status 6 [redacted] and age 60 to <70 years<br>6. Clinical Status 6 [redacted] and age >=70 years |

#### 5.2. Primary Endpoint(s) Analyses

##### 5.2.1. Definition of Endpoint(s)

Participants are free of respiratory failure if they are in category 1, 2, 3 or 4 from the GlaxoSmithKline (GSK) modified ordinal scale adapted from World Health Organization (WHO) scale 2020 (See Table 4 - Clinical Status using Ordinal Scale).

Participants will meet the endpoint if they are alive and free of respiratory failure at Day 28.  As participants may be discharged from the investigator site prior to Day 28, analysis windows will be used to define the endpoint, further details on analysis windows will be provided in the Output and Programming Specification (OPS).

##### Table 4 Clinical Status using Ordinal Scale

**Ordinal Scale (GSK modified version, adapted from WHO, 2020 version)**

CCI - This section contained Clinical Outcome Assessment data collection questionnaires or indices, which are protected by third party copyright laws and therefore have been excluded.

[redacted]

##### 5.2.2. Main Analytical Approach

The primary endpoint will be summarized using counts and proportions of the number of participants alive and free of respiratory failure and will be analysed using a logistic regression model, adjusting for treatment, clinical status category at baseline and age group. The outcome of the model is an odds ratio for the treatment effect, a confidence interval and p-value for the odds of improvement in proportion of participants alive and free of respiratory failure at Day 28, where an odds ratio >1 indicates improvement. The corresponding difference in proportion and confidence interval will be calculated [Ge, 2011].

In addition, unadjusted relative risks and risk differences will be calculated in addition to the adjusted odds ratios. These will be presented with 95% Wald confidence intervals. Any endpoint where an odds ratio/hazard ratio above 1 indicates improvement of otilimab over placebo, the inverse odds ratio/hazard ratio/relative risk will also be presented.

In addition, a detailed summary of participant ventilation status (participant died, invasive mechanical ventilation, BIPAP, CPAP, high-flow oxygen therapy, low-flow nasal cannula/prongs, low-flow oxygen face mask, other, none) will be summarised for each day using a stacked bar chart.

###### 5.2.2.1. Decision Criteria

Guidelines for stopping the study for futility (*e.g.* lack of efficacy) or overall success (*e.g.* evidence of efficacy) based on the primary endpoint are detailed below.

Futility will formally be assessed 28 days after approximately 160 participants have been randomised and will then be assessed 28 days after approximately 280, 400 and 600 participants have been randomised. Study success will be assessed 28 days after approximately 400 and 600 participants have been randomised.

Study stopping criteria have been defined using group sequential design methodology, using a Lan-DeMets alpha-spending function to control the type I error to 2.5% one-sided, with four interim analyses for futility, using Pocock analogue rules with a beta-spend of 10%; and two for efficacy, using the O'Brien-Fleming analogue rules [Lan, 1983].

Table 5 gives the stopping boundaries based on p-values (one-sided) from the analysis and also on the Z score scale. The p-values or Z scores at each interim analysis will be plotted against the boundaries and if either the futility or efficacy success boundary is crossed the study should be recommended to stop.

The boundaries are based on the estimated amount of information at each interim. If the amount included in interim analyses 1-4 differs from these values, the actual boundaries will be determined based on the exact amount of information at the time of the interim analyses. Note: The boundary at interim 5 is based on the estimated amount of information per the planned sample size of N=800, it will not be updated based on the actual sample size.

**Table 5 Formal Stopping Boundaries for Futility and Efficacy Success**

| Stage / Formal Analysis | Target Sample Size | Efficacy Success Boundaries | | Futility Boundaries | |
|---|---|---|---|---|---|
| | | p-value | Z-Score | p-value | Z-Score |
| 1 | 160 | | | >0.6005 | <-0.2548 |
| 2 | 280 | | | >0.3938 | <0.2693 |
| 3 | 400 | <0.0015 | >2.9626 | >0.2308 | <0.7363 |
| 4 | 600 | <0.0092 | >2.3590 | >0.0744 | <1.4441 |
| 5 – Final Analysis | 800 | <0.0220 | >2.0141 | >0.0220 | <2.0141 |

As recruitment has completed prior to formal interim analysis number 4 (on approximately 600 participants), it was decided to remove this formal pre-specified analysis on approximately 600 participants and continue to the final analyses only. The final analysis boundary has been adjusted to carry over the alpha that would have been spent on the 600-participant analysis as per Table 6. Previous boundaries were not amended.

**Table 6 Final Analysis Stopping Boundaries for Futility and Efficacy Success**

| Stage / Formal Analysis | Target Sample Size | Efficacy Success Boundaries | | Futility Boundaries | |
|---|---|---|---|---|---|
| | | p-value | Z-Score | p-value | Z-Score |
| Final Analysis | 800 | <0.0244 | >1.9697 | >0.0244 | <1.9697 |

##### 5.2.3. Sensitivity Analyses

As a sensitivity to the missing data, the following sensitivity analyses will be performed.

###### 5.2.3.1. Tipping Point Analyses

For the tipping point analyses, the underlying response rate among those participants with missing response status in each arm will be tested ranging between 0 and 1. This analysis will be two-dimensional, i.e. will allow for assumptions about the assumed response rate (and thus missing outcomes) in the two arms to vary independently, furthermore they will include scenarios where dropouts on otilimab have worse outcomes than dropouts on placebo. For combinations of the assumed response rates in the two arms, the number of additional responders among participants with missing response will be imputed multiple times by drawing from a binomial distribution. The log-odds ratio and associated standard error for each imputed dataset will be calculated using a logistic regression with treatment as the only covariate and results combined using Rubin's rules to calculate the test statistic and the corresponding p-value. Results will be presented via a heatmap.

##### 5.2.4. Supplementary Analyses

As a supplementary strategy, a composite estimand may be considered for the handling of the intercurrent event of use of specific COVID-19 medications. Specifically, a participant may be considered to be a non-responder if they have taken specific COVID-19 medications post-baseline, further details will be provided in a separate technical document.

##### 5.2.5. Subgroup Analyses

Subgroup analyses of the age group and clinical status at baseline will be performed using the same methodology as the primary endpoint with the addition of a treatment by subgroup interaction. A joint test of the interaction term will be provided in the summary table. Results will be presented as per primary analyses and, in addition a forest plot of the difference in proportions will be generated.

For subgroup analyses, if there are <10 events in a subgroup then only a summary will be produced.

Additional subgroup analyses may be performed and will be described in a separate technical document.

#### 5.3. Secondary Endpoint(s) Analyses

Secondary endpoints will be analysed at the final analysis only. The estimands are described in Table 3

##### 5.3.1. Endpoint Definition

###### 5.3.1.1. All-cause Mortality at Day 60

Participants will meet the endpoint if they have died due to any cause prior to Day 60.

If the date of death is unknown refer to Section 5.1.5 for missing data handling rules.

###### 5.3.1.2. Time to All-cause Mortality up to Day 60

Defined as the time (days) from dosing to death, due to any cause, up to (and including) Day 60.

###### 5.3.1.3. Participants Alive and Free of Respiratory Failure at Day 7, 14, 42, and 60

Defined as per primary endpoint, see Section 5.2.1.

###### 5.3.1.4. Time to Recovery from Respiratory Failure up to Day 28

Defined as the time (days) from dosing to last recovery from respiratory failure up to (and including) Day 28. Participants are in respiratory failure if they are in category 5 or above from the GlaxoSmithKline (GSK) modified ordinal scale adapted from World Health Organization (WHO) scale 2020 (Table 4).
Note: if a participant has recovered from respiratory failure but then progresses back into respiratory failure on or prior to the end of follow up (Day 28) the participant has not had their last recovery from respiratory failure.

This endpoint will use a composite approach for death such that if the intercurrent event of death occurs (on or prior to Day 28), then time will be censored at end of follow up (Day 28).

###### 5.3.1.5. Participants Alive and Independent of Supplementary Oxygen at Day 7, 14, 28, 42, and 60

Participants are independent of supplementary oxygen if they are in category 1, 2 or 3 from the GlaxoSmithKline (GSK) modified ordinal scale adapted from World Health Organization (WHO) scale 2020 (See Table 4- Ordinal Scale). Alternatively, if the ordinal scale is unknown but ventilation or oxygen status is available, then the endpoint is defined as the first full day where oxygen support is not required.

Participants will meet the endpoints if they are alive and independent of supplementary oxygen at Day 7, 14, 28, 42, and 60 respectively. As participants may be discharged from investigator site prior to Day 28, analysis windows will be used to define the endpoint, further details on analysis windows will be provided in the Output and Programming Specification (OPS).

###### 5.3.1.6. Time to Last Dependence on Supplementary Oxygen up to Day 28

Defined as the time (days) from dosing to last dependence on supplementary oxygen up to (and including) Day 28. Participants are dependent on supplementary oxygen if they are in category 4 or above from the GlaxoSmithKline (GSK) modified ordinal scale adapted from World Health Organization (WHO) scale 2020 (Table 4). Alternatively, If the ordinal scale is unknown but ventilation or oxygen status is available, then the endpoint is defined as the last day up to (and including) Day 28 where oxygen support is required.
Note: if a participant is independent of supplementary oxygen but then progresses back

into dependence of supplementary oxygen on or prior to the end of follow up (Day 28) the participant has not had their last dependence on supplementary oxygen.

This endpoint will use a composite approach such that if the intercurrent event of death occurs (on or prior to Day 28), then time will be censored at end of follow up (Day 28).

###### 5.3.1.7. ICU Admission up to Day 28

Participants will meet the endpoint if they have been admitted to the ICU up to (and including) Day 28, only evaluated for participants not in ICU at time of dosing.

Participants who die prior to Day 28 without being admitted ICU will be considered to have met the endpoint (composite estimands strategy).

###### 5.3.1.8. Time to Final ICU Discharge up to Day 28

This is defined as the time (days) from dosing to when the participant is discharged from the ICU for the last time up to (and including) Day 28. It will only be evaluated for participants in the ICU at baseline.

This endpoint will use a composite approach such that if the intercurrent event of death occurs (on or prior to Day 28), then time will be censored at the end of follow-up (Day 28), including those that were discharged from ICU prior to death.

###### 5.3.1.9. Time to First Discharge from Investigator Site up to Day 60

This is defined as the time (days) from dosing to when the participant is first discharged from investigator site up to (and including) Day 60.

This endpoint will use a composite approach such that if the event has not occurred prior to the intercurrent event of death (on or prior to Day 60), then time will be censored at the end of follow-up (Day 60).

###### 5.3.1.10. Time to First Discharge to Non-Hospitalised Residence up to Day 60

This is defined as the time (days) from dosing to when the participant is discharged to a non-hospitalised residence for the first time up to (and including) Day 60.

This endpoint will use a composite approach such that if the event has not occurred prior to the intercurrent event of death occurred (on or prior to Day 60), then time will be censored at the end of follow-up (Day 60).

##### 5.3.2. Main Analytical Approach

###### 5.3.2.1. Binary Endpoints

The binary endpoints:

- All-cause mortality at Day 60
- Participants alive and free of respiratory failure at Day 7, 14, 42, and 60

- Participants alive and independent of supplementary oxygen at Day 7, 14, 28, 42, and 60
- ICU admission up to Day 28

Binary secondary endpoints will be analysed as per Section 5.2.2.

Additionally, for ICU admission, the counts and proportion of participants in ICU at baseline, entered ICU post-baseline and those that never entered ICU will be produced. In addition, the counts and proportion of participants who die prior to and post ICU admission will be produced. Note: In ICU at Baseline, Entered ICU Post Baseline and Never entered ICU are mutually exclusive, participants may only enter ICU post Baseline if they were not in ICU at Baseline.

###### 5.3.2.2. Time to Event Endpoints

The following endpoints will be analysed using time to event analyses with a follow-up to Day 60:

- Time to all-cause mortality
- Time to first discharge from investigator site
- Time to first discharge to non-hospitalised residence

The following exploratory endpoints will be analysed using time to event analyses with a follow-up to Day 28:

- Time to recovery from respiratory failure
- Time to last dependence on supplementary oxygen
- Time to final ICU discharge

For time to event endpoints where the event is a worsening participant status, participants with missing data due to study withdrawal prior to occurrence of the event will be censored at the time of study withdrawal, participants alive at the end of follow-up will be censored at the time of end of follow-up (i.e. Day 28/60).

For time to event endpoints where the event is an improvement participant status, participants with missing data due to study withdrawal prior to first occurrence of the event will be censored at the time of end of follow-up. The end of follow up will be the earliest of the nominal timepoint (i.e. Day 28/60) and their actual study day of visit. If a participant meets the endpoint at the point of withdrawal, then the participant will be assumed to have achieved the event.

The endpoint will be analysed using a Cox proportional hazards model adjusted by treatment, clinical status and age group at baseline. Ties will be broken using Efron's method. The hazard ratio of the treatment group and corresponding confidence interval and p-value will be reported. For any endpoint where a hazard ratio above 1 indicates improvement of otilimab over placebo, the inverse hazard ratio will also be presented. For model checks refer to Section 6.3.3.

Kaplan-Meier plots will be presented by treatment group. Estimates of the median time-to-event and other relevant percentiles will be derived from the unadjusted Kaplan-Meier plots and reported in a table. If 50% of participant do not meet the event definition during the study, alternative percentiles may be produced.

Kaplan-Meier plots will be presented by treatment group.

Additionally, for time to ICU discharge, the following summaries will be produced:

- For participants who are in the ICU at any point during the trial, the duration of ICU stay will be summarised separately for participants that have died and those that complete the study, it will be further split by participants ICU status (in ICU at dosing or entered ICU post-dose).
- Additionally, for all participants a count and proportion of participants with stays in the ICU of 0 days, 1-<7 days, 7-<14 days and ≥14 days will be produced.

##### 5.3.3. Sensitivity Analyses

###### 5.3.3.1. Binary Endpoints

Sensitivity analyses as per Section 5.2.3 will be performed on the binary endpoints in the hierarchy specified in Section 2.1 (Participants alive and free of respiratory failure at Day 14, All-cause mortality at Day 60). Sensitivity analyses on additional endpoints may be investigated, further details will be provided in a separate technical document.

###### 5.3.3.2. Time to Event Endpoints

As a sensitivity to the competing risk of death an alternative model will be investigated to analyse the time to event endpoints in the hierarchy specified in Section 2.1 (time to recovery from respiratory failure up to Day 28). Analysis will be performed as follows:

- Competing risks via Fine-Gray method will be conducted. The competing risk for time to recovery from respiratory failure will be death. The model will be adjusted for clinical status and age group. In this case, the hazard ratio of the treatment group and corresponding confidence interval and p-value will be reported.

Sensitivity analyses on additional endpoints may be investigated, further details will be provided in a separate technical document.

##### 5.3.4. Supplementary Analyses

Supplementary estimands as described in Section 5.2.4 may be performed.

##### 5.3.5. Subgroup Analyses

Subgroup analyses may be performed as per Section 5.2.5 and will use the methods described above.

#### 5.4. Exploratory Endpoint(s) Analyses

Exploratory endpoints may be analysed at the final analysis only.

##### 5.4.1. Definition of Endpoint(s)

###### 5.4.1.1. Invasive Mechanical Ventilation (if not previously initiated) up to Day 28

Participants will meet the endpoint if they have had invasive mechanical ventilation initiated up to (and including) Day 28. This will only be evaluated for participants who are not supported using invasive mechanical ventilation at baseline.

Participants who die on or prior to Day 28 without the initiation of invasive mechanical ventilation will be considered to have met the endpoint (composite estimands strategy).

###### 5.4.1.2. Time to Invasive Mechanical Ventilation (if not previously initiated) up to Day 28

Time from dosing (days) to first use of invasive mechanical ventilation or death up to (and including) Day 28 as defined in Section 5.4.1.1. This will only be evaluated for participants who are not supported using invasive mechanical ventilation at baseline.

This endpoint will use a composite approach for death such that for participants who die on or prior to Day 28 without the initiation of invasive mechanical ventilation, the time to invasive mechanical ventilation will be set as the time of death.

###### 5.4.1.3. Alive and Not Invasively Mechanically Ventilated up to Day 28

Participants will meet the endpoint if they are alive and not requiring invasive mechanical ventilation at Day 28. This will be evaluated only for participants supported using invasive mechanical ventilation at baseline

###### 5.4.1.4. Time to Definitive Extubation up to Day 28

For participants supported using invasive mechanical ventilation at baseline this is defined as the time from dosing (days) to final extubation from invasive mechanical ventilation up to (and including) Day 28.

This endpoint will use a composite approach for death such that if the event has not occurred prior to the intercurrent event of death (on or prior to Day 28), then time will be censored at the end of follow-up (Day 28), including those that were extubated prior to death.

###### 5.4.1.5. Recovery or Improvement of at Least 2 Points Relative to Baseline of Sequential Organ Failure Assessment (SOFA) score up to Day 28

The SOFA score will be evaluated for participants in ICU with a SOFA score greater than 2 at baseline and analysed using a composite strategy to handle the intercurrent events as below.

Participants will meet the endpoint if they have an improvement of at least 2 points relative to baseline in SOFA score (derived where lower values indicate improvement) or they have recovered (discharged from ICU or investigator site for any other reason other than advanced facilitated care).

| Intercurrent event | Response Status |
|---|---|
| Death | Non-responder |
| Discharge from ICU or Investigator Site to Advanced facilitated care | Non-responder |
| Discharge from ICU or Investigator Site for any other reason other than advanced facilitated care | Responder |

In addition, change from baseline in SOFA score will be summarized for participants in ICU at baseline up to Day 28. This will be summarised using the trimmed means approach given in Section 5.4.2.4.

For the trimmed means, if >50% of the participants in ICU at baseline experience the intercurrent events listed above at any timepoint, then the data will be summarised up to that timepoint only.

If a participant experiences multiple intercurrent events, then the worst response status will be assumed.

###### 5.4.1.6. Recovery or Improvement Relative to Baseline in Blood Oxygen Saturation ($SpO_2$)

Change from Baseline in $SpO_2$, calculated as daily average of all assessments, for participants who are not invasively mechanically ventilated at baseline. $SpO_2$ will be analysed using a composite strategy to handle the intercurrent events as below.

Participants will meet the endpoint if they have an improvement relative to baseline in $SpO_2$ (derived where higher values indicate improvement) or they have recovered (participant is discharged from investigator site for any other reason other than advanced facilitated care or participant is independent from supplementary oxygen and $SpO_2$ is missing).

| Intercurrent event | Response Status |
|---|---|
| Death | Non-responder |
| Discharge from investigator site to advanced facilitated care | Non-responder |
| Use of Invasive Mechanical Ventilation | Non-responder |
| Discharge from Investigator Site for any other reason other than advanced facilitated care | Responder |
| Independence from supplementary oxygen and missing $SpO_2$ | Responder |

In addition, change from baseline in $SpO_2$ will be summarized for participants not invasively mechanically ventilated at baseline up to Day 28. Participants who have $SpO_2$ recorded whilst on invasive mechanical ventilation will be set to missing. $SpO_2$ will be summarized for observed data and using a trimmed means approach as specified in the Section 5.4.2.4.

For the trimmed means, if >50% of the participants not invasively mechanically ventilated at baseline experience the intercurrent events listed above at any timepoint, then the data will be summarised up to that timepoint only.

If a participant experiences multiple intercurrent events, then the worst response status will be assumed.

###### 5.4.1.7. Recovery or Improvement Relative to Baseline in Concentration of Inspired Oxygen ($FiO_2$)

Change from Baseline in $FiO_2$, calculated as daily average of all assessments. $FiO_2$ will be analysed using a composite strategy to handle the intercurrent events as below.

Participants will meet the endpoint if they have an improvement relative to baseline in $FiO_2$ (derived where lower values indicate improvement) or they have recovered (participant is discharged from investigator site for any other reason other than advanced facilitated care or participant is independent from supplementary oxygen and $SpO_2$ is missing). If $FiO_2$ is not recorded but Oxygen Flow Rate is, the $FiO_2$ will be calculated.

| Intercurrent event | Response Status |
|---|---|
| Death | Non-responder |
| Discharge from investigator site to advanced facilitated care | Non-responder |
| Discharge from Investigator Site for any other reason other than advanced facilitated care | Responder |
| Independence from supplementary oxygen and missing $SpO_2$ | Responder |

In addition, change from baseline in $FiO_2$ will be summarized for participants up to Day 28. $FiO_2$ will be summarized for observed data and using a trimmed means approach as specified in the Section 5.4.2.4.

For the trimmed means, if >50% of the participants experience the intercurrent events listed above at any timepoint, then the data will be summarised up to that timepoint only.

If a participant experiences multiple intercurrent events, then the worst response status will be assumed.

###### 5.4.1.8. Recovery or Improvement Relative to Baseline in $SpO_2/FiO_2$ Ratio

The ratio of $SpO_2/FiO_2$ will be calculated for each day using date-time matched values. Change from baseline in $SpO_2$/FiO2 will be calculated as average of the daily assessments, for participants who are not invasively mechanically ventilated. It will be analysed using a composite strategy to handle the intercurrent events as described in the individual endpoint sections. If a participant is a non-responder due to an intercurrent event for either $SpO_2$ or $FiO_2$ then they will be a non-responder for the ratio. Participants will meet the endpoint if they have an improvement relative to baseline in $SpO_2/FiO_2$ ratio (derived where higher values indicate improvement) or they have recovered (the participant is discharged from investigator site for any other reason other than advanced facilitated care or the participant is independent from supplementary oxygen and $SpO_2$ is missing).

###### 5.4.1.9. Oxygen-free Days up to Day 28

A participant is considered oxygen free on a day if they meet the definition in Section 5.3.1.5.
Intermittent data is expected after discharge from investigator site, therefore if participants have discontinued oxygen and have been discharged to a non hospitalised residence, they will be considered to be oxygen-free during the intermittent timepoints.

###### 5.4.1.10. Ventilator-free Days up to Day 28

A participant is considered ventilator-free on a day if they meet the definition for being alive and not invasively mechanically ventilated in Section 5.4.1.3.
Intermittent data is expected after discharge from investigator site, therefore if participants have discontinued ventilation and have been discharged to a non hospitalised residence, they will be considered to be ventilation-free during the intermittent timepoints.

###### 5.4.1.11. Time to Resolution of Pyrexia (>48h) up to Day 28

Time from dosing (days) to first resolution of pyrexia (defined as the first day of a >48-hour period with temperature below thresholds below) up to (and including) Day 28:

| Location | Pyrexia Threshold | Core Temperature Equivalent |
|---|---|---|
| Forehead | >36.3°C | >37.8°C |
| Axilla | >36.6°C | >37.8°C |
| Sublingual region | >37.2°C | >37.8°C |
| Rectum or Tympanic membrane | >37.8°C | >37.8°C |
| Pulmonary artery branch | >37.8°C | >37.8°C |
| Bladder | >37.8°C | >37.8°C |
| Esophagus | >37.8°C | >37.8°C |
| Nasopharynx | >37.8°C | >37.8°C |
| Brain | >37.8°C | >37.8°C |
| Other | >36.3°C (Minimum Pyrexia Threshold) | >37.8°C |

Evaluated for all participants with pyrexia at baseline.

This endpoint will use a composite approach such that if the intercurrent event of death occurs (on or prior to Day 28), then time will be censored at the end of follow-up (Day 28), including those that had resolution of pyrexia prior to death.

###### 5.4.1.12. Clinical Status Assessed Using an Ordinal Scale Assessed at Days 4, 7, 14, 28, 42, and 60

Clinical status assessed using an ordinal scale assessed at Days 4, 7, 14, 28, 42 and 60 see Table 4.

###### 5.4.1.13. Time to Improvement of at Least 2 Categories Relative to Baseline in Clinical Status up to Day 60

Time from dosing (days) to final improvement of at least 2 categories relative to baseline in clinical status (see Table 4) up to (and including) Day 60. Evaluated for all participants with clinical status greater than 2 at baseline.

Note: if a participant has an improvement of at least 2 categories relative to baseline but then progresses back to a less than 2 category improvement the participant has not had their final improvement of at least 2 categories.

This endpoint will use a composite approach such that if the event has not occurred prior to the intercurrent event of death (on or prior to Day 60), then time will be censored at the end of follow-up (Day 60).

###### 5.4.1.14. Change in COVID-19 Signs and Symptoms up to Day 60

Participants change in COVID-19 signs and symptoms up to Day 60 relative to dosing will be analysed.

Unadjusted relative risks, risk differences and odds ratios will be calculated and presented with 95% confidence intervals. For odds ratios, if the proportions are below 2% in any treatment group then exact 95% confidence interval will be calculated by inverting two one-sided (equal-tail) exact tests that are based on the noncentral hypergeometric distribution. All other confidence intervals will use the Wald method. A 2-sided Fisher's Exact Test will be performed, and p-values will be presented.

##### 5.4.2. Main Analytical Approach

###### 5.4.2.1. Time to Event Endpoints

The following exploratory endpoints will be analysed using time to event analyses with a follow-up to Day 28:

- Time to invasive mechanical ventilation (if not previously initiated)
- Time to definitive extubation
- Time to resolution of pyrexia (for at least 48h)

The following exploratory endpoints will be analysed using time to event analyses with a follow-up to Day 60:

- Time to improvement of at least 2 categories relative to baseline in clinical status

The endpoints will be analysed as per Section 5.3.2.2

###### 5.4.2.2. Binary Endpoints

The following exploratory endpoints will be analysed as binary endpoints:

- Invasive mechanical ventilation (if not previously initiated)
- Alive and not invasively mechanically-ventilated
- Improvement relative to baseline in $SpO_2$, $FiO_2$ and $SpO_2/FiO_2$ ratio
- Improvement of at least 2 points relative to baseline on SOFA score

Binary secondary endpoints will be analysed as per Section 5.2.2.

Additionally, for Invasive mechanical ventilation, the counts and proportion of participants for whom prone position was, and was not, used will be produced.

###### 5.4.2.3. Ordinal Endpoints

The clinical status assessed using an ordinal scale assessed at Days 4, 7, 14, 28, 42, and 60 will be summarized using counts and proportions of the number of participants in each category of the ordinal scale and will be analysed using a proportional odds logistic regression model, adjusting for treatment, ordinal scale category at baseline and age group. The outcome of the model is an odds ratio, a confidence interval and p-value for the odds of improvement on the ordinal scale, where an odds ratio >1 indicates improvement in clinical status.

In addition a shift table from baseline to the clinical status at Day 4, 7, 14, 28, 42 and 60 including a row for improvement (defined as moving from a higher category to a lower category) will be produced and the proportion of participants in each category of the ordinal scale will be summarised at each visit using a stacked bar chart.

###### 5.4.2.4. Continuous Endpoints

The following exploratory endpoints will be analysed as continuous endpoints:

- Change of Sequential Organ Failure Assessment (SOFA) score
- Change in $SpO_2$, $FiO_2$, and $SpO_2/FiO_2$ ratio

Continuous endpoints will be summarised using the trimmed means approach (Permutt, 2017) where the proportion of data to be trimmed will be determined by the amount of missing data due to intercurrent events as defined in Section 5.1.4.

Study withdrawal will be treated as a negative event if it occurs prior to one of the listed intercurrent events. If the subject has recovered (has one of the intercurrent events listed within the endpoint definition) prior to withdrawal then they will be considered to be recovered post-withdrawal.

Missing data without reason (no accompanying intercurrent event or withdrawal) will be treated as missing at random and will not count towards the trimmed sample.

###### 5.4.2.5. Count Endpoints

The following exploratory endpoints will be analysed as count data

- Oxygen-free days
- Ventilator-free days

Summary statistics of the number of days will be presented and the data will be analysed using negative binomial regression adjusting for clinical status at baseline and age group.

##### 5.4.3. Sensitivity Analyses

No sensitivity analyses planned.

##### 5.4.4. Supplementary Analyses

No supplementary analyses planned.

##### 5.4.5. Subgroup Analyses

No subgroup analyses are planned.

#### 5.5. Safety Analyses

##### 5.5.1. Extent of Exposure

As this is a single dose study, summaries of exposure will be limited to the number of participants exposed and the number of participants with interruptions or infusion stopped early and not completed. This will be presented with participant disposition.

##### 5.5.2. Adverse Events

Adverse events analyses including the analysis of adverse events (AEs) and Serious AEs (SAEs) will be based on GSK Core Data Standards. All adverse events reported will be treatment emergent adverse events.

An overview summary of AEs, including counts and percentages of participants with

- any AE
- AEs related to study intervention
- AEs leading to permanent discontinuation of study intervention,
- study intervention related AEs leading to permanent discontinuation of study intervention,

- AEs leading to dose interruption,
- Common AEs, with common AE defined as any AE with an incidence of at least 5% in any of the treatment group
- SAEs,
- SAEs related to study intervention,
- fatal SAEs
- fatal SAEs related to study intervention

Adverse events will be coded using the standard Medical Dictionary for Regulatory Affairs (MedDRA dictionary).

A summary of number and percentage of participants with any adverse events by maximum intensity will be produced. AEs will be sorted by Preferred term (PT) in descending order. The summary will use the following algorithms for counting the participant:

- **Preferred term row**: Participants experiencing the same AE preferred term several times with different intensities will only be counted once with the maximum grade.
- **Any event row**: Each participant with at least one adverse event will be counted only once at the maximum intensity no matter how many events they have.

The frequency and percentage of AEs (all intensities) will be summarized and displayed in two ways: 1) in descending order by PT only and 2) in descending order by SOC and PT. In the SOC row, the number of participants with multiple events under the same SOC will be counted once.

A separate summary will be provided for study intervention-related AEs. A study intervention-related AE is defined as an AE for which the investigator classifies the possible relationship to study intervention as “Yes”. A worst-case scenario approach will be taken to handle missing relatedness data, *i.e.* the summary table will include events with the relationship to study intervention as ‘Yes’ or missing. The summary table will be displayed by PT only.

All SAEs will be tabulated based on the number and percentage of participants who experienced the event. Separate summaries will also be provided for study intervention-related SAEs. The summary tables will be displayed by PT.

A study intervention-related SAE is defined as an SAE for which the investigator classifies the relationship to study intervention as “Yes”. A worst-case scenario approach will be taken to handle missing data, *i.e.* the summary table will include events with the relationship to study intervention as ‘Yes’ or missing.

Unadjusted Relative risks with Wald confidence intervals based on observed frequencies, for the proportions of participants with common (>=5% in either treatment group) AEs will be calculated for otilimab versus placebo.

Relative risks will not be calculated if there are no events in either of the two treatment arms being compared. The relative risks and exact confidence interval will be plotted on log base 10 scale.

The adverse events with calculated relative risks will be presented in decreasing order as per the IDSL standard.

###### 5.5.2.1. Adverse Events of Special Interest

The following will be considered adverse events of special interest (AESI) for the purpose of analyses:

- Cytokine release syndrome (CRS)
- Serious hypersensitivity reactions
  Note: these will be adjudicated by the SRT.
- Infusion site reactions
- Neutropenia ≥ Grade 3 ($<1.0 \times 10^9$/L)
- Serious infections

Listings of event characteristics will be provided for each AESI respectively. For AESIs with more than 8 events a summary of event characteristics will be provided for each AESI respectively, including number and percentage of participants with any event, number and percentage of events, maximum intensity, outcomes and the action taken for the event. The worst-case approach will be applied at participant level for the maximum intensity, *i.e*. a participant will only be counted once as the worst case from all the events experienced by the participant.

##### 5.5.3. Additional Safety Assessments

###### 5.5.3.1. Laboratory Data

Summaries of worst-case grade increase from baseline grade will be provided for all the lab tests that are gradable by a modified CTCAE v5. These summaries will display the number and percentage of participants with a maximum post-baseline grade increasing from their baseline grade. Any increase in grade from baseline will be summarized along with any increase to a maximum grade of 3 and any increase to a maximum grade of 4. Missing baseline grade will be assumed as grade 0. For laboratory tests that are graded for both low and high values, summaries will be done separately and labelled by direction, *e.g.*, sodium will be summarized as hyponatremia and hypernatremia separately.

The CTCAE v5 grades were modified to remove grading criteria that cannot be derived programmatically with the data collected (see footnotes). The modified CTCAE v5 grades are defined as follows:

| Laboratory parameters of interest[1] | Grade | | | |
|---|---|---|---|---|
| | 1 | 2 | 3 | 4 |
| HEMOGLOBIN decreased (CTCAE term is Anemia)[2] | <LLN - 10.0 g/dL; <LLN - 6.2 mmol/L; <LLN - 100 g/L | <10.0 - 8.0 g/dL; <6.2 - 4.9 mmol/L; <100 - 80 g/L | <8.0 g/dL; <4.9 mmol/L; <80 g/L | - |
| HEMOGLOBIN increased | >0 - 2 g/dL | >2 - 4 g/dL | >4 g/dL | - |
| LEUKOCYTE (White blood cell) decreased | <LLN - 3000/mm3; <LLN - 3.0 x 10e9 /L | <3000-2000/mm3; <3.0 - 2.0 x 10e9 /L | <2000-1000/mm3; <2.0 - 1.0 x 10e9 /L | <1000/mm3; <1.0 x 10e9 /L |
| LYMPHOCYTE COUNT decreased | <LLN - 800 /mm3; <LLN - 0.8 x 10e9/L | <800 - 500 /mm3; <0.8 - 0.5 x 10e9/L | <500 - 200 /mm3; <0.5 - 0.2 x 10e9/L | <200/mm3; <0.2 x 10e9/L |
| LYMPHOCYTE COUNT increased | - | >4,000 - 20,000 /mm3; >4 - 20 x 10e9/L | >20,000 /mm3; >20 x 10e9/L | - |
| NEUTROPHIL COUNT decreased | <LLN - 1,500 /mm3; <LLN - 1.5 x 10e9/L | <1,500 - 1,000 /mm3; <1.5 - 1.0 x 10e9/L | <1,000 - 500 /mm3; <1.0 - 0.5 x 10e9/L | <500 /mm3; <0.5 x 10e9/L |
| PLATELET COUNT decreased | <LLN - 75,000 /mm3; <LLN - 75.0 x 10e9/L | <75,000 - 50,000 /mm3; <75.0 - 50.0 x 10e9/L | <50,000 - 25,000 /mm3; <50.0 - 25.0 x 10e9/L | <25,000 /mm3; <25.0 x 10e9/L |
| AST (SGOT) | >ULN - 3.0 x ULN if baseline was normal; 1.5 - 3.0 x baseline if baseline was abnormal | >3.0 - 5.0 x ULN if baseline was normal; >3.0 - 5.0 x baseline if baseline was abnormal | >5.0 - 20.0 x ULN if baseline was normal; >5.0 - 20.0 x baseline if baseline was abnormal | >20.0 x ULN if baseline was normal; >20.0 x baseline if baseline was abnormal |
| ALP | >ULN - 2.5 x ULN if baseline was normal; 2.0 - 2.5 x baseline if baseline was abnormal | >2.5 - 5.0 x ULN if baseline was normal; >2.5 - 5.0 x baseline if baseline was abnormal | >5.0 - 20.0 x ULN if baseline was normal; >5.0 - 20.0 x baseline if baseline was abnormal | >20.0 x ULN if baseline was normal; >20.0 x baseline if baseline was abnormal |

| Laboratory parameters of interest[1] | Grade | | | |
|---|---|---|---|---|
| | 1 | 2 | 3 | 4 |
| ALT (SGPT) | >ULN - 3.0 x ULN if baseline was normal; 1.5 - 3.0 x baseline if baseline was abnormal | >3.0 - 5.0 x ULN if baseline was normal; >3.0 - 5.0 x baseline if baseline was abnormal | >5.0 - 20.0 x ULN if baseline was normal; >5.0 - 20.0 x baseline if baseline was abnormal | >20.0 x ULN if baseline was normal; >20.0 x baseline if baseline was abnormal |
| BLOOD BILIRUBIN increased | >ULN - 1.5 x ULN if baseline was normal; > 1.0 - 1.5 x baseline if baseline was abnormal | >1.5 - 3.0 x ULN if baseline was normal; >1.5 - 3.0 x baseline if baseline was abnormal | >3.0 - 10.0 x ULN if baseline was normal; >3.0 - 10.0 x baseline if baseline was abnormal | >10.0 x ULN if baseline was normal; >10.0 x baseline if baseline was abnormal |
| ALBUMIN (Hypo-albuminemia) | <LLN - 3 g/dL; <LLN - 30 g/L | <3 - 2 g/dL; <30 - 20 g/L | <2 g/dL; <20 g/L | - |
| APTT Activated partial thromboplastin time prolonged | >ULN - 1.5 x ULN | >1.5 - 2.5 x ULN | >2.5 x ULN | - |
| CALCIUM (Hypercalcemia) | Corrected serum calcium of >ULN - 11.5 mg/dL; >ULN - 2.9 mmol/L; Ionized calcium >ULN - 1.5 mmol/L | Corrected serum calcium of >11.5 - 12.5 mg/dL; >2.9 - 3.1 mmol/L; Ionized calcium >1.5 - 1.6 mmol/L | Corrected serum calcium of >12.5 - 13.5 mg/dL; >3.1 - 3.4 mmol/L; Ionized calcium >1.6 - 1.8 mmol/L | Corrected serum calcium of >13.5 mg/dL; >3.4 mmol/L; Ionized calcium >1.8 mmol/L |
| CALCIUM (Hypocalcemia) | Corrected serum calcium of <LLN - 8.0 mg/dL; <LLN - 2.0 mmol/L; Ionized calcium <LLN - 1.0 mmol/L | Corrected serum calcium of <8.0 - 7.0 mg/dL; <2.0 - 1.75 mmol/L; Ionized calcium <1.0 - 0.9 mmol/L | Corrected serum calcium of <7.0 - 6.0 mg/dL; <1.75 - 1.5 mmol/L; Ionized calcium <0.9 - 0.8 mmol/L | Corrected serum calcium of <6.0 mg/dL; <1.5 mmol/L; Ionized calcium <0.8 mmol/L |
| CPK increased | >ULN - 2.5 x ULN | >2.5 x ULN - 5 x ULN | >5 x ULN - 10 x ULN | >10 x ULN |
| CREATININE increased | >ULN - 1.5 x ULN | >1.5 - 3.0 x baseline; >1.5 - 3.0 x ULN | >3.0 x baseline; >3.0 - 6.0 x ULN | >6.0 x ULN |

| Laboratory parameters of interest[1] | Grade | | | |
|---|---|---|---|---|
| | 1 | 2 | 3 | 4 |
| GGT increased | >ULN - 2.5 x ULN if baseline was normal; 2.0 - 2.5 x baseline if baseline was abnormal | >2.5 - 5.0 x ULN if baseline was normal; >2.5 - 5.0 x baseline if baseline was abnormal | >5.0 - 20.0 x ULN if baseline was normal; >5.0 - 20.0 x baseline if baseline was abnormal | >20.0 x ULN if baseline was normal; >20.0 x baseline if baseline was abnormal |
| GLUCOSE (Hypoglycemia) | <LLN - 55 mg/dL; <LLN - 3.0 mmol/L | <55 - 40 mg/dL; <3.0 - 2.2 mmol/L | <40 - 30 mg/dL; <2.2 - 1.7 mmol/L | <30 mg/dL; <1.7 mmol/L |
| LDH (Blood lactate dehydrogenase increased) | >ULN | - | - | - |
| POTASSIUM (Hyperkalemia) | >ULN – 5.5 mmol/L | >5.5 – 6.0 mmol/L; | >6.0 – 7.0 mmol/L | >7.0 mmol/L |
| POTASSIUM (Hypokalemia)[3] | <LLN – 3.0 mmol/L | | <3.0 – 2.5 mmol/L | <2.5 mmol/L |
| SODIUM (Hypernatremia) | >ULN – 150 mmol/L | >150 - 155 mmol/L | >155 - 160 mmol/L | >160 mmol/L |
| SODIUM (Hyponatremia)[4] | <LLN - 130 mmol/L | - | 120-129 mmol/L | <120 mmol/L |
| INR increased[5] | >1.2 – 1.5 | >1.5 – 2.5 | >2.5 | - |
| Eosinophils[6] | >ULN and >Baseline | - | - | - |

1. Removal of cardiac troponin I or T increased gradings. Specifically Grade 1 indicated by levels above the upper limit of normal and below the level of myocardial infarction as defined by the manufacturer and Grade 3 indicated by levels consistent with myocardial infarction as defined by the manufacturer
2. Removal of Grade 3 defined as transfusion indicated by transfusion and grade 4 indicated by Life-threatening consequences or urgent intervention indicated
3. Removal of Grade 2 defined as symptomatic with <LLL – 3.0 mmol/L
4. Removal of Grade 2 defined as 125-129 mmol/L and asymptomatic, Grade 3 is re-defined as 120-129 mmol/L i.e. conservatively including the grade 2 criteria of "125-129 mmol/L and asymptomatic" into the Grade 3 criteria of "120-124 mmol/L regardless of symptoms; 125-129 mmol/L symptomatic" as asymptomatic/symptomatic is not collected.
5. Removal of Grade 1 indicated by >1 – 1.5 x baseline if on anticoagulation; Grade 2 indicated by >1.5 – 2.5 x baseline if on anticoagulation; and Grade 3 indicated by >2.5 x baseline if on anticoagulation.
6. Removal of Grade 3 defined as steroids initiated

Note: The grading will be rederived for any laboratory parameters with modifications from the CTCAE V5

For lab tests that are not gradable by modified CTCAE v5, summaries of worst-case changes from baseline with respect to normal range will be generated. Decreases to low, changes to normal or no changes from baseline, and increases to high will be summarized for the worst-case post-baseline. If a participant has a decrease to low and an increase to high during the same time interval, then the participant is counted in both the "Decrease to Low" categories and the "Increase to High" categories.

Separate summary tables for haematology, and chemistry laboratory tests will be produced. Liver function laboratory tests will be included with chemistry lab tests.

In addition, if any AE of CRS symptoms is reported a summary table of hematology, CRP, D-dimer and ferritin will be produced.

Summaries of hepatobiliary laboratory events will be provided in addition to what has been described above. Possible Hy's law cases defined as any elevated alanine aminotransferase (ALT) ≥ 3 × upper limit of normal (ULN) and total bilirubin ≥ 2 × ULN (>35% direct bilirubin) or ALT ≥ 3 × ULN and INR > 1.5 will be presented in a plot of maximum ALT vs maximum total bilirubin.

###### 5.5.3.2. Vital Signs

Summaries of worst-case systolic blood pressure (SBP), diastolic blood pressure (DBP) and heart rate relative to Potential Clinical Importance (PCI) post-baseline relative to baseline will be provided. This will include all post-baseline assessments.

Potential Clinical Importance ranges are defined as follows:

| Vital Sign Parameter | Units | Clinical Concern Range | |
|---|---|---|---|
| | | Lower | Upper |
| **Absolute** | | | |
| Systolic blood pressure | mmHg | <80 | >170 |
| Diastolic blood pressure | mmHg | <45 | >110 |
| Heart Rate | bpm | <40 | >100 |
| **Change from Baseline** | | | |
| Systolic blood pressure | mmHg | ≥30↓ | ≥30↑ |
| Diastolic blood pressure | mmHg | ≥20↓ | ≥20↑ |

Worst case summaries will display the number and percentage of participants with changes in the absolute values "To Low", "To w/in Range or No Change" or "To High" post baseline relative to baseline. Participants will be reported in both the "To Low" and "To High" categories if both of these changes are observed post-baseline. Participants will only be reported in the "To w/in Range or No Change" category if they experience no "To Low" or "To High" changes post-baseline. The summaries will also display the number and percentage of participants with "PCI Increase from Baseline", "PCI Decrease from Baseline" or "No PCI Change from Baseline". Participants will be reported in the "PCI Increase from Baseline" (/"PCI Increase from Baseline") categories if the upper (/lower) change from baseline clinical concern range value is observed, respectively. Participants will be reported in both the "PCI Increase from Baseline" and "PCI Decrease from Baseline" categories if both of these changes are observed post-baseline. Participants will only be reported in the "No PCI Change from Baseline" category if they experience no "PCI Increase from Baseline" or "PCI Decrease from Baseline" changes post-baseline.

In addition, summaries of change from baseline in systolic blood pressure (SBP), diastolic blood pressure (DBP), respiratory rate and temperature over Days 1 to 28 will be provided.

###### 5.5.3.3. ECG

A summary of the number and percentage of participants with ECG findings will be summarized by treatment. The ECG findings to be summarized are the ECG interpretation and clinical significance of abnormal ECGs. Participants with missing baseline values will be excluded from this summary.

#### 5.6. Other Analyses

##### 5.6.1. Pharmacokinetics (PK)

Pharmacokinetic concentrations will be listed and summarised by visit.

##### 5.6.2. Population Pharmacokinetics

Sparse PK samples will be analysed using population PK approach. Otilimab PK data from this study will be combined with PK data after IV administration from 4 studies in healthy volunteers, RA and MS adult participants. The PK parameters reported will be clearance, steady-state volume of distribution and AUC.

Further details will be provided in a supplemental analyses plan.

##### 5.6.3. Pharmacodynamics

Pharmacodynamic biomarkers measures including free GM-CSF, key markers of inflammation including, but not limited to CRP, serum ferritin and inflammatory cytokines as appropriate will be listed and summarised by visit.

Further details will be provided in a supplemental analyses plan.

##### 5.6.4. Pharmacokinetics/Pharmacodynamics

Exposure-response relationship for key efficacy, safety and PD endpoints will be explored graphically and if data permits (range of otilimab concentrations observed in this study is wide enough) will be followed by model-based analysis.

Further details will be provided in a supplemental analyses plan.

#### 5.7. Interim Analyses

Details of the interim analyses and required outputs are provided in the IDMC Charter and IDMC Statistical Analyses Plan.

### 6. SUPPORTING DOCUMENTATION

#### 6.1. Appendix 1 Abbreviations and Trademarks

##### 6.1.1. List of Abbreviations

| Abbreviation | Description |
|---|---|
| AE | Adverse Event |
| AESI | Adverse Event of Special Interest |
| ANCOVA | Analyses of Covariance |
| AUC | Area Under the Curve |
| BIPAP | Bilevel Positive Airway Pressure |
| CL | Clearance |
| COVID-19 | Corona Virus Disease 2019 |
| CPAP | Continuous Positive Airway Pressure |
| CRP | C-Reactive Protein |
| CRS | Cytokine Release Syndrome |
| CTCAE | Common Terminology Criteria for Adverse Events |
| DBP | Diastolic Blood Pressure |
| ECG | Electrocardiogram |
| eCRF | Electronic Case Record Form |
| FCS | Fully Conditional Specification |
| $FiO_2$ | Concentration of Inspired Oxygen |
| FU | Follow-up |
| GM-CSF | Granulocyte-macrophage colony-stimulating factor |
| GSK | GlaxoSmithKline |
| ICH | International Conference on Harmonization |
| ICU | Intensive Care Unit |
| IDMC | Independent Data Monitoring Committee |
| IDSL | Integrated Data Standards Library |
| IND | Investigational New Drug |
| IRT | Interactive Response Technology |
| IV | Intravenous |
| MAR | Missing at random |
| MCMC | Markov Chain Monte Carlo |
| MI | Multiple imputation |
| MITT | Modified Intent-To-Treat |
| MS | Multiple Sclerosis |
| OPS | Output and Programming Specification |
| PCI | Potential Clinical Importance |
| PD | Pharmacodynamic |
| PK | Pharmacokinetic |
| PT | Preferred Term |
| RA | Rheumatoid Arthritis |
| RMST | Restricted Mean Survival Time |
| SAE | Serious Adverse Event |

| Abbreviation | Description |
|---|---|
| SAP | Statistical Analysis Plan |
| SBP | Systolic Blood Pressure |
| SDTM | Study Data Tabulation Model |
| SOC | System Organ Class |
| SOFA | Sequential Organ Failure Assessment |
| $SpO_2$ | Blood Oxygen Saturation |
| SRT | Safety Review Team |
| ULN | Upper Limit of Normal |
| WHO | World Health Organisation |

##### 6.1.2. Trademarks

| Trademarks of the GlaxoSmithKline Group of Companies |
|---|
| None |

| Trademarks not owned by the GlaxoSmithKline Group of Companies |
|---|
| None |

#### 6.2. Appendix 2: Changes to Protocol-Planned Analyses

Changes to protocol-planned analyses include:

- The number of interim analyses in the group sequential design has been reduced to three interims for futility and one for efficacy. This is due to recruitment being completed prior to formal interim analyses number 4 (on approximately 600 participants), hence it was decided to remove this formal analysis on approximately 600 participants and continue to the final analyses only. The final analyses boundary has been adjusted to carry over the alpha that would have been spent on the 600-participant analysis. The hierarchy has been updated accordingly but previous boundaries were not amended.
- The study design has been updated such that the final analyses on the primary endpoint will be conducted once all participants have been randomised and completed to Day 28 or withdrawn.
- The proposed hierarchy for testing secondary endpoints, refer to Section 2 for full details.
- Updates to secondary endpoint related to hospital discharge, refer to Section 5.3.1.9 and Section 5.3.1.10 for full details.
- For exploratory endpoints on improvement in SOFA, Blood Oxygen Saturation (SpO2), Concentration of Inspired Oxygen (FiO2) and SpO2/FiO2 ratio, the endpoints have been clarified to include recovery as part of composite strategy.
- The exploratory endpoint “Time to Improvement of at Least 2 Categories Relative to Baseline on an Ordinal Scale up to Day 60” has been updated to “Time to Improvement of at Least 2 Categories Relative to Baseline in Clinical Status up to Day 60” for consistency over the use of clinical status rather than ordinal scale.

#### 6.3. Appendix 3: Statistical Modelling

##### 6.3.1. Multiple Imputation

Multiple imputation will be utilized to impute data that is missing following withdrawal of the participant from the study. Each data type will use one set of imputations with all endpoints that depend on that data type being derived from the intermediate dataset.

The statistical model used for the multiple imputation data generation will use one of two approaches depending on data type:

- Continuous endpoints will use the Markov Chain Monte Carlo (MCMC) method with adjustment for covariates.
- Binary and ordinal endpoints will use a two-stage approach:
    1. First intermittent missing data will be imputed using MCMC followed by adaptive rounding [Carpenter, 2012] step to create a monotone structure.
    2. Second a monotone logistic regression imputation will be performed to impute data post-study withdrawal, this model will include the analyses covariates. If logistic regression step fails to converge the following back-up options including:
        - i. A discriminant function [Brand, 1999] may be used in place of logistic.
        - ii. MCMC with adaptive rounding.

A sufficient number of imputations will be performed to ensure the stability of the estimates, the current plan is 10,000 imputation. The results of analysis from each complete imputed dataset will be combined using Rubin's rule. Table 7 details the models, covariates and the seed to be used for the analyses.

For subgroup analyses the multiple imputation model will be identical to the main endpoint analyses and the interaction term will be added into the analysis step. The chi-squared test statistic of the interaction term will be pooled using the procedure of Rubin (1987) and Li (1991).

**Table 7	Multiple Imputation Specifications**

| Endpoint | Model | Covariates | Post Baseline<br><br>Timepoints Included in Prediction Model | Initial Seed |
|---|---|---|---|---|
| Participants alive and free of respiratory failure at Day 7, 14, 28, 42, and 60. | Monotone Binary Logistic Regression | Treatment, Clinical Status at Baseline, Age Group at Baseline | Days 7,14,28,42 and 60 | 3764 |
| All-cause mortality at Day 60. | Monotone Binary Logistic Regression | Treatment, Clinical Status at Baseline, Age Group at Baseline | Days 7,14,28,42 and 60 | 1734 |
| Participants alive and independent of supplementary oxygen at Day 7, 14, 28, 42, and 60 | Monotone Binary Logistic Regression | Treatment, Clinical Status at Baseline, Age Group at Baseline | Days 7,14,28,42 and 60 | 9849 |
| Admission to ICU up to Day 28 | Monotone Binary Logistic Regression | Treatment, Clinical Status at Baseline, Age Group at Baseline | Days 4, 7, 10, 14 and 28 | 6935 |
| If the number of participants is small within a category of a covariate, then the covariate categories may be refined. If the category cannot be refined further, then the covariate may be included as a continuous measure. | | | | |

The seeds were generated using the following code:

```
DATA seeds;
  DO i=1 to 4;
    seed=int(10000*ranuni(214094));
    OUTPUT;
  END;
RUN;
```

If additional seeds are required, then the initial seed as specified will be incremented by 1.

###### 6.3.1.1. Adaptive Rounding

The adaptive rounding algorithm [Carpenter, 2012] is:

1. For binary variable $j$ in imputed dataset $k = 1, \dots, K$, let $\bar{Y}_{j,k}$ denote the mean of the observed (binary) and imputed (continuous) values.
2. Construct the threshold $c_{j,k} = \bar{Y}_{j,k} - \Phi^{-1}(\bar{Y}_{j,k})\sqrt{\bar{Y}_{j,k}(1 - \bar{Y}_{j,k})}$
3. In imputed data set $k$, re-code continuous imputed values of the binary variable $Y_j$ according to the following rule: $Y_{i,j} \leq c_{j,k}$ becomes $Y_{i,j} = 0$, and $Y_{i,j} > c_{j,k}$ becomes $Y_{i,j} = 1$.

##### 6.3.2. Proportional Odds Regression Model Checking

To assess the assumption of proportional odds for the clinical status logistic regressions will be performed for each binary grouping:

$$Clinical\ Status \leq \ j\ vs\ Clinical\ Status > j),\ \ for\ j = 1, \dots 7$$

and for each covariate separately (treatment, clinical status and age group at baseline).

The coefficients of a given covariate will be tested for equality across the binary logistic regressions using a Wald test. This test being anti-conservative, the proportionality of the odds will be also be assessed graphically.

##### 6.3.3. Time to Event Model Checking

For the Cox proportional hazards model, the proportional hazards assumption will be assessed prior to model fitting using the following methods:

- Plot of log(-log(survival)) versus log(time) by treatment group: A non-parallel pattern is an indication of violation of the proportional hazard assumption.
- Plot of Schoenfeld residuals versus time for continuous covariates: A non-zero slope is an indication of a violation of the proportional hazard assumption.
- Evaluation of time-dependency of treatment effect by adding an interaction term of treatment and time in the Cox model. If the interaction term is significant ($p< 0.05$), it is considered that the proportional hazards assumption is violated.

If one or more of the procedures above demonstrates clear violation of the proportional hazards assumption, it will be considered the proportional hazards assumption does not hold.

##### 6.3.4. Count Data Model Checking

Distributional assumptions underlying the model used for analysis will be checked. If there are any departures from the distributional assumptions, alternative models may be explored as sensitivity analyses.

### 7. REFERENCES

Brand JPL. Development, Implementation, and Evaluation of Multiple Imputation Strategies for the Statistical Analysis of Incomplete Data Sets. Ph.D. thesis, Erasmus University 1999.

Carpenter JR, Kenward MG. Multiple Imputation and its Applications. Wiley 2012.

Ge, Murray, Meyer, *et al*. Covariate-adjusted difference in proportions from clinical trials using logistic regression and weighted risk differences. Drug Information Journal 2011;45:481-93

GSK Document Number: 2020N436091_02. Study ID: 214094. A randomized, double-blind, placebo-controlled, study evaluating the efficacy and safety of otilimab IV in patients with severe pulmonary COVID-19 related disease. Effective Date 02-JUL-2020.

Lan KKG, DeMets DL. Discrete sequential boundaries for clinical trials. Biometrika 1983;70:659-63.

Li, K.-H., Meng, X.-L., Raghunathan, T.E., and Rubin, D.B. Significance levels from repeated p-values with multiply-imputed data. Statistica Sinica, 1991;1:65-92

Permutt T, Li F. Trimmed means for symptom trials with dropouts. Pharm Stat 2017;16:20-8.

Rubin D.B. Multiple Imputation for Nonresponse in Surveys. John Wiley and Sons 1987.

Sanofi. Press release. https://www.sanofi.com/en/media-room/press-releases/2020/2020-04-27-12-58-00.

Van Buuren, S. Multiple Imputation of Discrete and Continuous Data by Fully Conditional Specification. Statistical Methods in Medical Research 2007;16:219–242.

WHO. https://www.who. int/blueprint/priority-diseases/key-action/COVID-19_Treatment_Trial_Design_Master_Protocol_synopsis_Final_18022020.pdf, 2020.
