## Supplementary material for "A Randomized Trial of Otilimab in Severe COVID-19 Pneumonia (OSCAR)": Protocol

### TITLE PAGE

**Protocol Title:** A randomized, double-blind, placebo-controlled, study evaluating the efficacy and safety of otilimab IV in patients with severe pulmonary COVID-19 related disease.

**Protocol Number**: 214094

| | |
|---|---|
| **Compound Number or Name:** | GSK3196165 (otilimab) |

**Study Name:** OSCAR (**O**tilimab in **S**evere **C**OVID-19 **R**elated Disease).

**Brief Title:** Investigating otilimab in patients with severe pulmonary COVID-19 related disease.

**Study Phase:** Phase 2

**Sponsor Name and Legal Registered Address:**

GlaxoSmithKline Research & Development Limited
980 Great West Road
Brentford
Middlesex, TW8 9GS
UK

**Regulatory Agency Identifying Number(s):**

**IND:** 149754

**EudraCT:** 2020-001759-42

**Approval Date:** 02-JUL-2020

Copyright 2020 the GlaxoSmithKline group of companies. All rights reserved. Unauthorized copying or use of this information is prohibited.

#### SPONSOR SIGNATORY

PPD

Dr Jatin Patel

Digitally signed by Dr Jatin Patel
Date: 2020.07.02 11:13:55 +01'00'

July 2nd 2020

Dr. Jatin Patel
Clinical Development Leader
Clinical Science (II and Fibrosis)

**Date**

**Medical Monitor Name and Contact Information** can be found in the Study Reference Manual

### PROTOCOL AMENDMENT SUMMARY OF CHANGES TABLE

| DOCUMENT HISTORY | | |
|---|---|---|
| **Document** | **Date** | **DNG Number** |
| Amendment 2 | 02-JUL-2020 | 2020N436091_02 |
| Amendment 1 | 18-MAY-2020 | 2020N436091_01 |
| Original Protocol | 30-APR-2020 | 2020N436091_00 |

**Amendment 2:** 02-JUL-2020

This amendment is considered to be non-substantial based on the criteria set forth in Article 10(a) of Directive 2001/20/EC of the European Parliament and the Council of the European Union.

**Overall Rationale for the Amendment:** modifications to the protocol in response to regulatory and ethics committee feedback, and clarifications based on investigator feedback. The main changes are (1) clarification that Day 1 pre-dose assessments do not need to be repeated if Screening and Randomization are within 24 hours; (2) expedited reporting of cytokine release syndrome (CRS) as an AESI; (3) clarification that organ transplant patients are excluded per Exclusion Criteria #11; (4) clarification of conditions by which convalescent plasma is permitted or not during the study; (5) removed specific mention of chloroquine and hydroxychloroquine; (6) expanded the null hypothesis; (7) consent may be collected before the 48h screening window.

| Section # and Name | Description of Change | Brief Rationale |
|---|---|---|
| **Section 1.3. Schedule of Activities** | Revised text that if Screening and Randomization are within 24 hours rather than the same day, there is no need to repeat Day 1 pre-dose assessments, (except Ordinal Scale and pre-infusion vital signs), and that Randomization and Dosing should be as close as possible.<br>Added note that consent may be collected before the 48h screening window. | Investigator's feedback. |
| **Section 2.3.1. Risk Assessment and Section 7.4.7.1. Cytokine Release Syndrome** | Added statement that in the event that new onset of CRS is suspected, the medical monitor should be notified immediately and under no circumstance should this exceed 24 hours, and the event must also be reported within 24 hours as an AE or an SAE. | Regulatory request. |
| **Section 4.4** | Definition of a participant who completes the study updated to include participants who die prior to Day 60. | Clarification of definition. |
| **Section 5.2. Exclusion Criteria #11** | Added note that participants with an organ transplant are excluded. | Ethics Committee request. |

| Section # and Name | Description of Change | Brief Rationale |
|---|---|---|
| **Section 5.2. Exclusion Criteria #13** | Added conditions by which convalescent plasma is not permitted during the study. | Investigator's feedback. |
| **Section 5.2. Exclusion Criteria #19** | Hemoglobin ≤9 g/dL. | Error in units. |
| **Section 6.6. Concomitant Therapy** | Added conditions by which convalescent plasma is permitted. | Investigator's feedback. |
| **Section 6.6.2. Medication/Treatment PERMITTED During the Study** | Removed specific mention of chloroquine and hydroxychloroquine. | No longer expected to be widely used following clinical trials results. |
| **Section 7.2.4. Ventilation status** | Added statement to record if the participant was in the prone or semi-prone position at any time whilst mechanically ventilated. | Investigator's feedback. |
| **Section 7.2.6. Ordinal Scale** | Added note for additional organ support. | Clarification. |
| **Section 7.3.1. Physical Examinations** | Clarification that physical exam results will not be recorded in the eCRF. | Investigator's feedback. |
| **Section 8.1. Statistical Hypotheses** | Expanded the null hypothesis. | Clarification of the null hypothesis. |
| **Section 9.1.3. Informed Consent Process** | Added note that consent may be collected before the 48h screening window. | Clarification. |
| **All sections** | Other minor, grammatical and typographical corrections to improve readability. | |

### 1. PROTOCOL SUMMARY

#### 1.1. Synopsis

A randomized, double-blind, placebo-controlled, study evaluating the efficacy and safety of otilimab IV in patients with severe pulmonary COVID-19 related disease.

**Brief Title:**

Investigating otilimab in patients with severe pulmonary COVID-19 related disease.

**Rationale:**

The aim of this study is to evaluate the benefit-risk of a single infusion of otilimab in the treatment of patients with severe pulmonary COVID-19 related disease.

**Objectives and Endpoints:**

| Objectives | Endpoints |
|---|---|
| **Primary** | |
| • To compare the efficacy of otilimab 90 mg IV versus placebo | • Participants alive and free of respiratory failure at Day 28 |
| **Secondary** | |
| • To compare the efficacy of otilimab 90 mg IV versus placebo | • All-cause mortality at Day 60<br>• Time to all-cause mortality up to Day 60<br>• Participants alive and free of respiratory failure at Day 7, 14, 42, and 60<br>• Time to recovery from respiratory failure up to Day 28<br>• Participants alive and independent of supplementary oxygen at Day 7, 14, 28, 42, and 60<br>• Time to last dependence on supplementary oxygen up to Day 28<br>• Admission to ICU up to Day 28<br>• Time to final ICU discharge up to Day 28<br>• Time to final hospital discharge up to Day 28 |
| • To compare the safety and tolerability of otilimab 90 mg IV versus placebo | • Occurrence of adverse events (AEs) [up to Day 60]<br>• Occurrence of serious adverse events (SAEs) [up to Day 60] |
| **Exploratory** | |
| • To compare the efficacy of otilimab 90 mg IV versus placebo | **Other endpoints up to Day 28**<br>• Invasive mechanical ventilation (if not previously initiated)<br>• Time to invasive mechanical ventilation (if not previously initiated)<br>• Alive and not invasively mechanically-ventilated<br>• Time to definitive extubation<br>• Improvement of at least 2 points relative to baseline of Sequential Organ Failure Assessment (SOFA) score |

| Objectives | Endpoints |
|---|---|
| | • Improvement relative to baseline in $SpO_2$, $FiO_2$, and $SpO_2/FiO_2$ ratio<br>• Oxygen-free days<br>• Ventilator-free days<br>• Time to resolution of pyrexia (for at least 48h)<br>**Other endpoints up to Day 60**<br>• Clinical status assessed using an ordinal scale[1] assessed at Days 4, 7, 14, 28, 42, and 60<br>• Time to improvement of at least 2 categories relative to baseline on an ordinal scale[1]<br>• Change in COVID-19 signs and symptoms |
| • To determine the pharmacokinetics (PK) profile of otilimab | **PK Endpoints up to Day 14**<br>• Otilimab apparent clearance (CL/F) and other PK parameters as appropriate using sparse PK sampling |
| • To determine:<br> • Exposure-response.<br> • Pharmacodynamic (PD) biomarkers<br> • Changes in key markers of inflammation | **PD Endpoints up to Day 28**<br>• Exposure-response relationship for key efficacy, safety and PD endpoints<br>• Key markers of inflammation including, but not limited to CRP, serum ferritin and inflammatory cytokines as appropriate |

[1]**Ordinal Scale (GSK modified version, adapted from WHO, 2020 version)**

CCI - This section contained Clinical Outcome Assessment data collection questionnaires or indices, which are protected by third party copyright laws and therefore have been excluded.

[redacted]

**Overall Design:**

This study is a multi-center, randomized, double-blind, placebo-controlled trial to assess the efficacy and safety of otilimab for the treatment of severe pulmonary COVID-19 related disease. The study population consists of hospitalized participants with new onset hypoxia requiring significant oxygen support or requiring invasive mechanical ventilation (≤48 hours before dosing). All participants will receive standard of care as per institutional protocols, in addition to study treatment.

**Brief Summary:**

Participants will be recruited from two mutually-exclusive categories within the ordinal scale:

CCI - This section contained Clinical Outcome Assessment data collection questionnaires or indices, which are protected by third party copyright laws and therefore have been excluded.

[redacted]

The first 20 participants (initial safety cohort) will be from category 5 CCI [redacted]

Since this is the first time that otilimab has been tested in participants with severe pulmonary COVID-19 related disease, initial dosing will be staggered, and safety data reviewed. Unless a safety signal is identified during the ongoing safety review by the GSK safety review team (SRT) and periodic review by the Independent Data Monitoring Committee (IDMC), enrolment of the main cohort of participants will be initiated, and the enrolment rate will be capped to control randomization.

**Number of Participants:**

A maximum of approximately 800 participants will be randomly assigned to study intervention. Any participant who receives study intervention will be considered evaluable.

**Intervention Groups and Duration:**

Participants will be randomized 1:1 by interactive response technology (IRT) in a blinded manner to receive either a blinded 1-hour infusion of otilimab 90 mg or placebo IV in addition to standard of care. Participants will be assessed daily until Discharge (or Day 28, whichever is sooner), and followed up at Days 42 and 60 after randomization.

**Independent Data Monitoring Committee:** Yes

#### 1.2. Schema

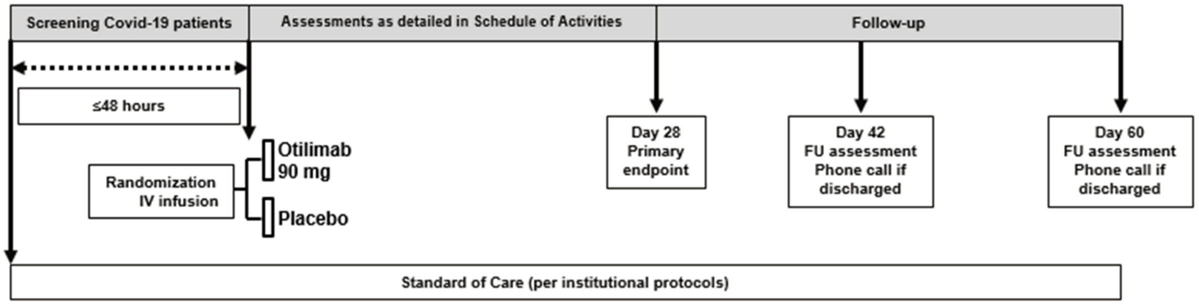

#### 1.3. Schedule of Activities (SoA)

See Section 7 for further details of study procedures listed in the SoA.

| Assessments | Screening[2)] | Day 1 | Days 2-7 | Day 14[4)] | Day 28[5)] | Discharge / Early Withdrawal[7)] | Follow-up Phone call (D42 & 60 / Early Withdrawal)[6), 7)] | Notes |
|---|---|---|---|---|---|---|---|---|
| Informed consent[3)] | X | | | | | | | |
| Eligibility assessment | X | | | | | | | |
| Thoracic CT scan or chest X-ray | X | | | | | | | Chest X-ray or CT scan must be available at Screening to confirm a diagnosis of pneumonia and eligibility. Record in the eCRF if a CT scan or chest X-ray is done between Screening and Discharge. |
| Demography | X | | | | | | | |
| Medical history | X | | | | | | | At Screening, record the signs and symptoms of COVID-19 at admission, as detailed in the eCRF. |
| Daily clinical features of COVID-19 (for non-mechanically ventilated participants) | | ←=============== Daily[1)] ===============→ | | | | | X | Daily review of COVID-19 signs and symptoms, as detailed in the eCRF. |
| Full physical examination | X | | | | | X | | |
| 12-lead ECG | X | X | | | | X | | **Day 1 - record before dosing.** |
| Weight | X | | | | | | | |
| Vital Signs: blood pressure, pulse, respiratory rate and temperature | X | ←=============== Daily[1)] ===============→ | | | | | | **Day 1 – measure pre-infusion and approximately 1 hour post-infusion**. Measure daily AM (at about same time daily), as per standard of care (SoC). |
| $SpO_2$ (for participants not on invasive mechanical ventilation) | X | ←=============== Daily[1)] ===============→ | | | | | | **Day 1 - record before dosing.** Measure daily AM and PM (at about same times daily), as per SoC. |
| Concentration of inspired oxygen ($FiO_2$) and / or oxygen flow rate (L/min) | X | ←=============== Daily[1)] ===============→ | | | | | | **Day 1 - record before dosing.** Measure daily AM and PM (at about same times daily), as per SoC. |
| Ventilation status, ICU admission and discharge, and ICU supportive care | X | ←=============== Daily[1)] ===============→ | | | | | X | Includes ventilation status (method of non-invasive ventilation or mechanical ventilation). Record participant admission or discharge from the ICU. Record oxygen treatment if used at home. |
| Ordinal scale | X | ←=============== Daily[1)] ===============→ | | | | | X | **Day 1 - record category <u>before randomization</u> (for stratification).** Record daily AM (at about same time daily). |
| SOFA score (ICU participants **ONLY**) | X | ←=============== Daily[1)] ===============→ | | | | | | **Day 1 - record before dosing.** Record daily AM (at about same time daily), as per SoC. |
| SAE/AE review | X | ←=============== Daily[1)] ===============→ | | | | | X | At Screening, only SAEs related to study participation or GSK concomitant medication will be recorded. |
| Concomitant medication review | X[8)] | ←=============== Daily[1)] ===============→ | | | | | X | |

| Assessments | Screening[2)] | Day 1 | Days 2-7 | Day 14[4)] | Day 28[5)] | Discharge / Early Withdrawal[7)] | Follow-up Phone call (D42 & 60 / Early Withdrawal)[6), 7)] | Notes |
|---|---|---|---|---|---|---|---|---|
| Survival follow-up | | | | | | X | X | |
| **Laboratory Assessments** | | | | | | | | |
| Local Laboratory tests[9)] | X | X | X[11)] | X | X | X | | **Day 1 - collect before dosing.** Only if not discharged. Refer to Table 3 for the list of laboratory assessments required. Collected at any time during the day, as convenient. |
| Pregnancy test | X | | | | | X | (verbal confirmation on Day 60) | Local urine or serum test at Screening and Discharge. Verbal confirmation of pregnancy status will be obtained at follow-up phone call. |
| PD sample (cytokines)[9)] | X[10)] | | X[11)] | | | | | Only if not discharged. Collected at any time during the day, as convenient. Sample collection time must be recorded. |
| PK sample | | X[12)] | X[13)] | X | | | | Only if not discharged. Samples after Day 1 can be collected at any time during the day, as convenient. Sample collection time must be recorded. |
| **Treatment Administration** | | | | | | | | |
| Randomization | | X | | | | | | |
| IV dosing with otilimab or placebo | | X | | | | | | 1-hour IV infusion. Record start and end times of the infusion. Randomization and Dosing should be as close as possible. |

1) Assessment to be conducted every day from Day 1 until Discharge (or until Day 28 if the participant remains hospitalized).
2) Up to 48 h before randomization. If Screening and Randomization are within 24h, there is no need to repeat Day 1 pre-dose assessments (except Ordinal Scale and pre-infusion vital signs).
3) Consent may be collected before the 48h screening window if the investigator considers that the participant is likely to become eligible, and wants to ensure that consent is given by the participant (rather than an LAR).
4) If the participant is discharged before Day 13, the Day 14 and Day 28 visits will be replaced with a phone call (±2 days). If the participant is discharged on Day 13, the Day 14 visit is not required, and the Day 28 visit will be a phone call (±2 days). When visits are replaced with a phone call, complete the same assessments as the Day 42 or 60 phone call.
5) If the participant is discharged between Day 15 and Day 27, the Day 28 visit will be replaced with a phone call (±2 days). When visits are replaced with a phone call, complete the same assessments as the Day 42 or Day 60 phone call.
6) Follow-up phone calls at Day 42 (±2 days) and Day 60 (±2 days). If the participant has not been discharged, these will be visits.
7) See Section 6.7 for information on Early Withdrawal.
8) At Screening, prior medications will be recorded from the 14 days before Screening.
9) If a cytokine release syndrome (CRS) is suspected (see Section 7.4.7.1), the following samples should be taken as soon as possible after suspected CRS onset and again 24 h later: Hematology, CRP, D-dimer, ferritin, PD sample (cytokines).
10) Sample can be collected anytime from Screening up to start of infusion.
11) Days 2, 4 and 7 only.
12) Immediately after infusion.
13) Days 2 and 7 only.

### 2. INTRODUCTION

Granulocyte-macrophage colony stimulating factor (GM-CSF) is a hemopoietic growth factor and a key mediator of tissue inflammation, that signals through the GM-CSFα/β receptor complex. GM-CSF has multiple cellular sources, including monocytes, macrophages, T cells, B cells, neutrophils and tissue resident cells [Wicks, 2016]. GM-CSF exerts multiple pro-inflammatory effects on myeloid cells including macrophages, monocytes and neutrophils. Pre-clinical models and human clinical trials have demonstrated that neutralizing GM-CSF or blocking the GM-CSFRα subunit can be efficacious in inflammatory diseases such as rheumatoid arthritis. Otilimab is a fully human anti-GM-CSF monoclonal antibody (mAb) that has been well tolerated by subjects and demonstrates anti-inflammatory activity in RA patients (phase 3 clinical trials ongoing).

Emerging data from COVID-19 patients indicate that GM-CSF may play an important role in the acute hyper-inflammatory state associated with severe stages of illness. Treatment with an anti-GM-CSF receptor (anti-GM-CSFR) mAb (mavrilimumab) was well tolerated, reduced fever and improved oxygenation in 6 patients experiencing a steep decline of pulmonary status due to COVID-19 pneumonia. All patients were reported to respond to treatment, and 3 out of the 6 patients were discharged within 5 days, further supporting the scientific rationale to evaluate the efficacy of otilimab in these patients.

#### 2.1. Study Rationale

A subset of patients with SARS-CoV-2 infection develop pulmonary complications and may transition to a form of acute respiratory distress syndrome (ARDS) and the most severe stage of illness, which manifests as an extra-pulmonary, systemic hyper-inflammation syndrome [Siddiqi, 2020]. Markers of systemic inflammation appear to be elevated at this disease stage and studies have shown that inflammatory cytokines such as IL-2, IL-7, G-CSF, MCP-1, MIP-1α and TNFα were significantly elevated, particularly in COVID-19 patients in requiring ICU treatment compared with those not requiring ICU treatment [Huang, 2020]. Elevated levels of IL-6 have also been detected in COVID-19 patients and have been correlated with adverse patient outcomes [Coomes, 2020].

In healthy subjects, GM-CSF is not detectable in the circulation, however, GM-CSF was detectable in plasma of some COVID-19 patients whether they received ICU care or not [Huang, 2020]. In a separate study, increased numbers of GM-CSF-expressing monocytes, T -cells, B cells and NK cells were found in the blood of COVID-19 ICU and non-ICU patients compared with healthy controls. The COVID-19 ICU patient group also had increased numbers of $CD4^+$ T cells co-expressing GM-CSF and IFNγ, as well as increased numbers of $GM\text{-}CSF^+$ $CD8^+$ T cells compared with non-ICU patients [Zhou, 2020].

GM-CSF is well known to drive monocytes/macrophages into proinflammatory phenotypes that are primed to produce inflammatory cytokines including TNFα, IL-1β and IL-6 [Hamilton, 2019; Wicks, 2016]. The observation that GM-CSF neutralization prevented IL-6, IL-8 and MCP-1 cytokine release by monocytes in a co-culture CAR T-cell assay [Sachdeva, 2019], and cytokine release in a humanized CAR T-cell xenograft mouse model [Sterner, 2019], suggests a role for GM-CSF as a driver of hyper-inflammation and cytokine release syndrome (CRS).

Therefore, by blocking GM-CSF signaling, an anti-GM-CSF mAb such as otilimab is hypothesized to work “upstream” of IL-6 in the hyper-inflammatory processes triggered by SARS-CoV-2 infection and will directly inhibit proinflammatory activities of monocytes/macrophages (*e.g.* by reducing TNFα, IL-1β and IL-6 production) that perpetuate inflammation and cause tissue damage.

Otilimab, an anti-GM-CSF mAb, is currently in phase 3 development (ContRAst program) for rheumatoid arthritis (RA) following phase 2 results, which showed a positive efficacy and safety profile based on a large clinically-driven response for joint improvement and pain.

In addition, lenzilumab (KB003), another anti-GM-CSF mAb dosed IV over 20 weeks was generally well tolerated in a placebo-controlled study in inadequately controlled asthma patients, with no safety signals of concern and no increase in infection rate [Molfino, 2016]. Moreover, there were no dose limiting toxicities identified and no treatment emergent, study drug-related, grade 3 or 4 toxicities reported in a phase 1 study of lenzilumab dosed in 15 patients with chronic myelomonocytic leukemia [Patnaik, 2020].

Otilimab has not previously been tested in patients with severe pulmonary COVID-19 related disease. This study aims to show that otilimab improves the respiratory status of patients with severe pulmonary COVID-19 related disease compared with standard of care. In order to understand risk and benefit in a stepwise fashion, the study has a leading cohort with detailed safety assessments to provide early evidence of safety prior to the larger cohort of patients to provide further confirmatory efficacy and safety data that may facilitate early access to patients.

#### 2.2. Background

##### 2.2.1. Role of GM-CSF in inflammation

Originally identified as a hematopoietic growth factor, GM-CSF is now recognized as a key driver of tissue inflammation that exerts multiple pro-inflammatory effects on myeloid cells. The cellular sources of GM-CSF include T cells, B cells, monocytes, macrophages, neutrophils and tissue resident cells [Wicks, 2016] and GM-CSF signals via the GM-CSFα/β receptor complex which is expressed on myeloid cells. GM-CSF promotes survival and polarizes monocytes/macrophages into pro-inflammatory phenotypes that are primed to produce pro-inflammatory cytokines including TNFα, IL-1β and IL-6 [Hamilton, 2019]. GM-CSF also extends neutrophil survival, primes the neutrophil oxidative burst, enhances phagocytosis and promotes neutrophil adhesion to endothelial cells and promotes neutrophil trafficking to sites of inflammation [Wicks, 2016].

Therapeutic inhibition of GM-CSF signaling by mavrilimumab in RA subjects has been shown to reduce circulating serum IL-6 levels and indirectly suppress T-cell activation [Guo, 2018]. Efficacy and safety data from clinical trials in rheumatoid arthritis (RA) patients indicate that inhibition of GM-CSF or its receptor is anti-inflammatory and well tolerated with long-term dosing [Burmester, 2018; Taylor, 2019].

##### 2.2.2. Scientific rationale to inhibit GM-CSF in COVID-19 patients

Some patients with severe acute respiratory syndrome–coronavirus-2 (SARS-CoV-2) infection develop pulmonary complications and may transition to acute respiratory distress syndrome (ARDS) and the most severe stage of illness, which manifests as an extra-pulmonary, systemic hyper-inflammation syndrome. It is widely proposed that intervention with immunomodulatory therapies to reduce this hyper-inflammatory state could be beneficial, particularly in the later and more severe stages of COVID-19 disease [Mehta, 2020; Siddiqi, 2020].

COVID-19 pneumonia patients have serological markers associated with macrophage activation syndrome and the limited COVID-19 post mortem data available shows prominent alveolar edema, hyalonosis (intra-alveolar proteinosis) and fibrin deposition with pneumocytes viral cytopathic change and immune cell infiltration [McGonagle, 2020]. Pulmonary post-mortem findings from 38 COVID-19 cases from Italy have shown that inflammatory infiltrate was composed by macrophages in alveolar lumens and lymphocytes mainly in the interstice [Carsana, 2020]. Therefore, in later stages of severe disease suppressing hyper-inflammatory processes (“cytokine storm syndrome” [Mehta, 2020]) with an immunomodulator would be important to limit further lung damage, prevent respiratory failure, shock, and multi-organ failure.

In health, GM-CSF is not detectable in the circulation, however, GM-CSF was detectable in plasma of some COVID-19 patients whether they received ICU care or not [Huang, 2020]. In a separate study, increased numbers of GM-CSF-expressing monocytes, T-cells, B cells and NK cells were found in the blood of COVID-19 ICU and non-ICU patients compared with healthy controls. The COVID-19 ICU patient group also had increased numbers of $CD4^+$ T cells co-expressing GM-CSF and IFNγ, as well as increased numbers of $GM\text{-}CSF^+$ $CD8^+$ T cells compared with non-ICU patients [Zhou, 2020].

COVID-19 patients have increased numbers of circulating GM-CSF+ monocytes compared to healthy controls. ICU treated patients also have increased numbers of $IL\text{-}6^+$ monocytes and $CD14^+CD16^+$ monocytes compared with non-ICU patients [Zhou, 2020]. A small study has also shown that pro-inflammatory monocyte-derived macrophages dominate over resident alveolar macrophages in the lungs of patients with severe COVID-19, whereas resident alveolar macrophages predominated in the lungs of mild/recovering COVID-19 patients [Liao, 2020]. The existence of these monocyte-derived macrophages suggests that activated monocytes infiltrate the injured lung from the blood, release inflammatory cytokines and may differentiate into activated macrophages. It has been proposed that in severe or critical COVID-19 cases, since the lung barrier integrity is interrupted, monocytes and neutrophils are chemotactic to the infection site to clear exudates with virus particles and infected cells, resulting in uncontrolled inflammation. In this process, because of the substantial reduction and dysfunction of lymphocytes, the adaptive immune response cannot be effectively initiated, and the uncontrolled virus infection leads to more macrophage infiltration and a further worsening of lung injury [Li, 2020]. GM-CSF is well known to drive monocytes/macrophages into proinflammatory phenotypes that are primed to produce inflammatory cytokines including IL-6, TNF and IL-1β [Hamilton, 2019; Wicks, 2016].

The observation that GM-CSF neutralization prevented IL-6, IL-8 and MCP-1 cytokine release by monocytes in a co-culture CAR T-cell assay [Sachdeva, 2019], and that GM-CSF neutralization prevented human VEGF, IP10 and IL-1Rα cytokine release and murine IP10 and MCP-1 chemokine release in a humanized CAR T-cell xenograft mouse model [Sterner, 2019], suggests a role for GM-CSF as a driver of hyper-inflammation and cytokine release syndrome (CRS).

As well as GM-CSF, other markers of systemic inflammation are elevated in hospitalized COVID-19 patients [Huang, 2020]. Plasma levels of IL-1β, IL-1Rα, IL-6, IL-8, IL-9, IL-10, IFNγ, IP-10, MIP-1β, PDGF, and VEGF concentrations were higher in both ICU patients and non-ICU COVID-19 patients than in healthy controls. Plasma levels of IL-2, IL-7, G-CSF, MCP-1, MIP-1α and TNFα were significantly elevated in COVID-19 patients requiring ICU treatment compared with those not requiring ICU treatment [Huang, 2020]. Elevated levels of IL-6 are reported in COVID-19 patients and have been correlated with adverse patient outcomes [Coomes, 2020].

These observations support the hypothesis that hyper-inflammation leads to poor outcome in hospitalized patients, transition to ICU/ventilator and death due to cytokine storm and organ damage. In these patients there is a strong rationale that inhibiting hyper-inflammatory processes with targeted immunomodulatory approaches would be beneficial [Mehta, 2020; McGonagle, 2020].

##### 2.2.3. Timing of GM-CSF inhibition in COVID patients

GM-CSF is not produced in most healthy tissues, but local alveolar production of GM-CSF is necessary for mature alveolar macrophage differentiation and lung homeostasis function over the long term. Complete genetic or auto-antibody ablation of GM-CSF (or its receptor) eventually results in pulmonary alveolar proteinosis (PAP), however this is likely not to be a concern in severe pulmonary COVID-19 related disease since PAP takes many months/years to develop [Sakagami, 2010; Trapnell, 2019], and has not been observed following long-term inhibition of GM-CSF signaling with an anti-GM-CSF mAb (mavrilimumab) treatment in RA patients [Burmester, 2018].

Increased levels of GM-CSF in bronchoalveolar lavage (BAL) fluids on Day 1 and 3 of diagnosis with ARDS was associated with improved survival [Matute-Bello, 2000], and a second study of 29 critically-ill patients within 24 hours of diagnosis with ventilator-associated pneumonia (VAP) demonstrated that higher GM-CSF/TGFβ1 BAL ratios correlated with increased patient survival [Overgaard, 2015]. However, administration of recombinant GM-CSF by intravenous (IV) infusion daily for 14 days evaluated in a randomized phase II trial in 130 patients with acute lung injury (ALI)/ARDS had no effect on ventilator-free days, mortality or organ failure [Paine, 2012].

Intranasal administration of an anti-GM-CSFRα or anti-GM-CSF monoclonal antibody inhibited cell influx and inflammation in the lungs of mice challenged with LPS

[Alessandris, 2019; Bozinovski, 2002]. Neutralization of GM-CSF was shown to dose-dependently suppress neutrophilic inflammation, inhibit TNF mRNA expression, suppress matrix metalloprotease 9 (MMP9) levels and activity, and reduce macrophage replication [Bozinovski, 2002; Bozinovski, 2004].

Conversely, preclinical studies assessing influenza A infection in mice deficient in GM-CSF or its receptor have demonstrated a more severe viral infection in the absence of pulmonary GM-CSF signaling; whereas intranasal administration of GM-CSF to wildtype mice or GM-CSF overexpression in the airways of GM-CSF knockout mice reduced mortality following influenza A infection [Halstead, 2015; Halstead, 2018; Huang, 2011]. However, it has been speculated that at high influenza viral titer, persistent activation of GM-CSF as induced by highly pathogenic influenza strains, GM-CSF may contribute to injurious inflammation.[Halstead, 2015].

Thus, local lung expression of GM-CSF in the early stages of COVID-19 related pulmonary disease in patients may help control viral titer and to maintain the epithelial lung barrier. As the disease progresses to hypoxia, controlling the viral load is thought to be less important, whereas suppressing the systemic hyper-inflammatory process ("cytokine storm syndrome" [Mehta, 2020]) is much more important to limit further lung damage, prevent respiratory failure, shock, and multi-organ failure. Therefore, once hypoxia occurs in COVID-19 patients, it is reasonable to propose that reducing systemic hyper-inflammatory processes with targeted immunomodulatory approaches may be most beneficial [Mehta, 2020].

To support this, there have been several reports of anti-IL-6 mAbs having some benefit in small studies of patients with severe pulmonary COVID-19 related disease where beneficial clinical response has been inferred by outcomes such as reduction in fever, oxygen requirements, C-reactive protein (CRP), radiological pulmonary abnormalities [Roche, 2020; Xu, 2020], or need for ventilation [Hermine, 2020]. Moreover, review of interim data from an ongoing large phase 3 study of sarilumab led to a recommendation by the IDMC to continue only in the more advanced "critical" group whose outcomes had improved and discontinuing the less advanced "severe" group, without any new safety findings [Sanofi, 2020].

By blocking GM-CSF signaling, an anti-GM-CSF mAb such as otilimab is hypothesized to work "upstream" of IL-6 in the hyper-inflammatory processes triggered by SARS-CoV-2 infection and will directly inhibit proinflammatory activities of monocytes/macrophages (*e.g.* by reducing IL-6, TNF and IL-1 production) that perpetuate inflammation and cause tissue damage. In addition, as described previously, early experience in treating COVID-19 patients with severe pulmonary disease with an anti-GM-CSF receptor mAb (mavrilimumab) has been well tolerated and demonstrated clinical benefit through fever reduction and improved oxygenation.

#### 2.3. Benefit/Risk Assessment

The potential risk assessment and mitigation strategy for the administration of otilimab in this protocol is outlined below.

##### 2.3.1. Risk Assessment

More detailed information about the known and expected benefits and risks and reasonably expected adverse events of otilimab may be found in the Investigator's Brochure (IB). The assessment of potential risks and mitigation strategy for the administration of otilimab in this protocol is outlined below.

| Potential Risk of Clinical Significance | Summary of Data/Rationale for Risk | Mitigation Strategy |
|---|---|---|
| **Study Intervention(s) Otilimab** | | |
| Cytokine release syndrome (CRS) | CRS is a systemic inflammatory response caused by the release of inflammatory cytokines, such as IL-6, tumor necrosis factor alpha (TNFα), by lymphocytes (B cells, T cells, and/or natural killer cells) and/or myeloid cells (macrophages, dendritic cells, and monocytes). This can lead to a constellation of clinical symptoms including fever, flu-like symptoms, hypotension, to severe inflammatory syndrome leading to widespread organ dysfunction, even death. It is not expected that the inhibition of GM-CSF signaling, by administration of otilimab, would carry the risk of directly inducing CRS.<br>**Pre-Clinical Data**<br>GM-CSF neutralization in CAR T-cell assays or *in vivo* models inhibited cytokine release [Sachdeva, 2019; Sterner, 2019]. *In vitro* otilimab showed variable results, with evidence of cytokine elevation at elevated levels of otilimab (50 and 100 µg/mL). Primate studies demonstrated no evidence of CRS with concentrations at $C_{max}$ of 2,690 µg/mL (54x | **Mitigation:**<br>• To mitigate the uncertainty around safety and tolerability of otilimab IV, dosing will occur in an initial safety cohort of 20 non-mechanically ventilated participants, and these participants will be closely monitored and dosed in a staggered manner (see Section 4.1).<br>• Unless a safety signal is identified during the ongoing safety review by the GSK SRT and periodic review by the IDMC, enrolment of the main cohort of participants will be initiated.<br>• Careful dose selection.<br>• Exclusion of participants who do not require oxygen therapy, and estimated GFR ≤30 mL/min/1.73m$^2$ (Section 5.2, Exclusion Criteria).<br>**Monitoring:**<br>• All participants will be hospitalized during infusion and will be closely monitored for signs and symptoms of CRS (defined as a worsening of the CTCAE CRS Grade, Section 7.4.7.1. |

| Potential Risk of Clinical Significance | Summary of Data/Rationale for Risk | Mitigation Strategy |
|---|---|---|
| | higher than anticipated human levels) and substantial safety margin for AUCs.<br>**Clinical Data:**<br>There has been no evidence of cytokine release or symptoms suggestive of CRS in any human study with otilimab. PK data from IV administration of absolute doses of otilimab 90 mg or higher is available from a total of 46 HVs and patients, with a maximum concentration of 56 µg/mL in one HV. | Cytokine Release Syndrome) within 6 hours of infusion.<br>• CRS is categorized as an AESI.<br>• Evidence of CRS that occurs within 6 hours of infusion and demonstrated as worsening of CRS.<br>• In the event of CRS, investigators should follow institutional guidelines for symptom management.<br>• In the event that new onset of CRS is suspected, the medical monitor must be notified immediately and under no circumstance can this exceed 24 hours, and the event must also be reported within 24 hours as an AE or an SAE. |
| Serious hypersensitivity reactions | There is a potential risk of hypersensitivity reactions, including anaphylaxis, during and following the administration of protein-based products, such as otilimab.<br>**Clinical Data:**<br>No serious allergic or acute systemic reactions have been observed to date in the clinical development program. | **Mitigation:**<br>• Exclusion of participants with a history of allergic reaction, including anaphylaxis to any previous treatment with an anti-GM-CSF therapy (Section 5.2 Exclusion Criteria)<br>**Monitoring:**<br>• Serious hypersensitivity reactions are categorized as AESIs.<br>• Hypersensitivity to be managed appropriately per local guidelines/medical judgement<br>• All participants hospitalized during infusion and will undergo monitoring for hypersensitivity reactions |
| Infusion site reactions | Infusions may be associated with local reactions (*e.g*. swelling, induration, pain).<br>**Non-clinical Data:**<br>No macroscopic or microscopic changes indicative of local injection site reactions were observed following SC administration in cynomolgus monkeys. | **Monitoring:**<br>• Infusion site reactions are categorized as AESIs<br>• Monitor for infusion site reactions throughout study. |

| Potential Risk of Clinical Significance | Summary of Data/Rationale for Risk | Mitigation Strategy |
|---|---|---|
| | **Clinical Data:**<br>Injection/infusion site reactions have been reported in completed Phase II studies with otilimab and were non-serious and of mild to moderate intensity. | |
| Neutropenia ≥ Grade 3 (<1.0 x $10^9$/L) | Although there is a perceived theoretical risk that GM-CSF blockade may affect maturation of leukocytes and their precursors, mice lacking GM-CSF do not develop neutropenia or show any major perturbation of hematopoiesis [Stanley, 1994].<br>**Clinical Data:**<br>Neutropenia has been observed in completed studies of otilimab; however, no clinically significant cases have been observed. | **Mitigation:**<br>Exclusion of participants with ANC <1.5 x $10^9$/L (neutropenia ≥ Grade 2) (Section 5.2 Exclusion Criteria)<br>**Monitoring:**<br>• Neutropenia Grade 3 (<1.0 x $10^9$/L) is categorized as an AESI.<br>• Full blood count (with differential) performed throughout hospitalization. |
| Infections | Immune-modulating biologic drugs are associated with an increased risk of serious and opportunistic infections. Because of the role of GM-CSF in anti-infective immunity, otilimab also has the potential to increase the risk of infection. GM-CSF has been noted to play a role in helping the lung fight infection and recover from any injury. Therefore, it is possible that otilimab, by blocking GM-CSF, may affect the ability of the lungs to fight lung infection or properly recover from infection<br>**Non-clinical Data:**<br>Studies in knock-out mice showed that GM-CSF deficiency affects the ability of mice to control infection when infected *with M. tuberculosis* or pulmonary group B streptococcus [Paine, 2001; LeVine, 1999].<br>No changes in peripheral blood populations (lymphocytes, neutrophils, monocytes, eosinophils or basophils), phagocytic activity of peripheral blood | **Eligibility Criteria**<br>Exclusion of participants with:<br>• **Untreated** systemic bacterial, fungal, viral, or other infection (other than SARS-CoV-2).<br>• Known active tuberculosis (TB), history of untreated or incompletely treated active or latent TB, suspected or known extrapulmonary TB<br>• Known HIV regardless of immunological status<br>• Known HBsAg and/or anti-HCV positive<br>• Absolute neutrophil count (ANC) <1.5 x $10^9$/L (neutropenia ≥ Grade 2)<br>• Received monoclonal antibody therapy (*e.g.* tocilizumab, sarilumab) within the past 3 months prior to randomization, including intravenous immunoglobulin, or planned to be received during the study<br>• Received immunosuppressant therapy including but not limited to cyclosporin, azathioprine, tacrolimus, mycophenolate, JAK inhibitors (*e.g.* |

| Potential Risk of Clinical Significance | Summary of Data/Rationale for Risk | Mitigation Strategy |
|---|---|---|
| | polymorphonuclear cells (investigational endpoint in the 26 week study), T-cell dependent B-cell primary or secondary response, or circulating cytokine levels (26 week study) were observed.<br>**Clinical Data:**<br>Based on the mechanism of action of otilimab an increased risk of infection including TB, fungal and opportunistic infections could be expected for anti-GM-CSF treatment, because of the role of GM-CSF in anti-infective immunity.<br>One HV in study MSC-1000 experienced septic shock secondary to pneumonia 29 days after receiving a single dose of 1.5 mg/kg but recovered after treatment with antibiotics.<br>In Phase II completed studies, no significant infections or opportunistic infections were reported. | baricitinib, tofacitinib, upadacitinib) within the last 3 months prior to randomization or planned to be received during the study<br>• Currently receiving chronic oral corticosteroids for a non-COVID-19 related condition in a dose higher than prednisone 10 mg or equivalent per day.<br>**Monitoring:**<br>• Serious and opportunistic infections are categorized as AESIs.<br>• Participants will be hospitalized under direct medical supervision and closely monitored for signs of infection as part of SoC with appropriate diagnostic tests and antibiotic treatment, as necessary.<br>• Routine laboratory assessments will be obtained throughout hospitalization. |

AESI=Adverse Event of Special Interest, ANC= Absolute Neutrophil Concentration, AUC=Area Under the Curve, CAR=Chimeric Antigen Receptor, $C_{max}$=Maximum Concentration, CRS=Cytokine Release Syndrome, CTCAE=Common Terminology Criteria for Adverse Events, HV=Healthy Volunteer, GFR=Glomerular Filtration Rate, IDMC=Independent Data Monitoring Committee, SC=Subcutaneous.

##### 2.3.2. Benefit Assessment

Otilimab is currently in Phase 3 clinical development for the treatment of RA, at weekly doses of 90 and 150 mg SC for up to 12 months in pivotal studies and thereafter eligible for treatment in a long-term extension study.

In completed studies (IV and SC), cumulatively 447 patients and healthy volunteers (HV) have received otilimab (326 RA patients, 25 multiple sclerosis (MS) patients, 22 hand osteoarthritis patients, and 74 HVs).

Although otilimab has not been studied before in this acutely ill population, there is biologic rationale which supports the use of otilimab in the treatment of COVID-19 patients with pulmonary complications.

GM-CSF can act as a proinflammatory cytokine that promotes myeloid cell survival and polarizes monocytes/macrophages into pro-inflammatory phenotypes that are primed to produce pro-inflammatory cytokines including TNFα, IL-1β and IL-6. GM-CSF also extends neutrophil survival, primes the oxidative burst, enhances phagocytosis and promotes neutrophil adhesion to endothelial cells and neutrophil trafficking to sites of inflammation.

COVID-19 ICU patients have increased circulating numbers of IL-6$^{+}$ monocytes, CD14$^{+}$CD16$^{+}$ monocytes and GM-CSF$^{+}$IFNγ$^{+}$ helper T cells compared with non-ICU COVID-19 patients. Once hypoxia occurs, stopping the systemic hyper-inflammatory processes to limit further damage to lung and other organs would the most important therapeutic approach. Preclinical data suggest that elevated levels of GM-CSF in the lung could be more beneficial in the early stages of disease and may play a role in disease resolution by helping control viral titer, restore or maintain the lung barrier function, and return to tissue homeostasis. As the disease progresses, suppressing the systemic hyper-inflammatory processes would be a priority and where otilimab could be beneficial. Therefore, a single dose of otilimab in patients with pneumonia and oxygenation impairment, would be expected to inhibit inflammation and still allow sufficient GM-CSF levels in the lung during the recovery phase.

##### 2.3.3. Overall Benefit: Risk Conclusion

The overall benefit-risk assessment takes into account the potential benefit of otilimab treatment through GM-CSF blockade and mitigation of the hyper-inflammatory response.

This hyper-inflammatory response associated with severe SARS-CoV-2 infection can result in severe or fatal pulmonary disease and multi-organ failure.

With the planned safety monitoring measures around the expected low risk of CRS and overall favorable safety profile to date, the overall benefit-risk assessment for the patient population proposed for this study is considered positive.

### 3. OBJECTIVES AND ENDPOINTS

| Objectives | Endpoints |
|---|---|
| **Primary** | |
| • To compare the efficacy of otilimab 90 mg IV versus placebo | • Participants alive and free of respiratory failure at Day 28 |
| **Secondary** | |
| • To compare the efficacy of otilimab 90 mg IV versus placebo | • All-cause mortality at Day 60<br>• Time to all-cause mortality up to Day 60<br>• Participants alive and free of respiratory failure at Day 7, 14, 42, and 60<br>• Time to recovery from respiratory failure up to Day 28<br>• Participants alive and independent of supplementary oxygen at Day 7, 14, 28, 42, and 60<br>• Time to last dependence on supplementary oxygen up to Day 28<br>• Admission to ICU up to Day 28<br>• Time to final ICU discharge up to Day 28<br>• Time to final hospital discharge up to Day 28 |
| • To compare the safety and tolerability of otilimab 90 mg IV versus placebo | • Occurrence of adverse events (AEs) [up to Day 60]<br>• Occurrence of serious adverse events (SAEs) [up to Day 60] |
| **Exploratory** | |
| • To compare the efficacy of otilimab 90 mg IV versus placebo | **Other endpoints up to Day 28**<br>• Invasive mechanical ventilation (if not previously initiated)<br>• Time to invasive mechanical ventilation (if not previously initiated)<br>• Alive and not invasively mechanically-ventilated<br>• Time to definitive extubation<br>• Improvement of at least 2 points relative to baseline of Sequential Organ Failure Assessment (SOFA) score<br>• Improvement relative to baseline in $SpO_2$, $FiO_2$, and $SpO_2/FiO_2$ ratio<br>• Oxygen-free days<br>• Ventilator-free days<br>• Time to resolution of pyrexia (for at least 48h)<br>**Other endpoints up to Day 60**<br>• Clinical status assessed using an ordinal scale[1] assessed at Days 4, 7, 14, 28, 42, and 60<br>• Time to improvement of at least 2 categories relative to baseline on an ordinal scale[1]<br>• Change in COVID-19 signs and symptoms |
| • To determine the pharmacokinetics (PK) profile of otilimab | **PK Endpoints up to Day 14**<br>• Otilimab apparent clearance (CL/F) and other PK parameters as appropriate using sparse PK sampling |

| Objectives | Endpoints |
|---|---|
| • To determine:<br>• Exposure-response.<br>• Pharmacodynamic (PD) biomarkers<br>• Changes in key markers of inflammation | **PD Endpoints up to Day 28**<br>• Exposure-response relationship for key efficacy, safety and PD endpoints<br>• Key markers of inflammation including, but not limited to CRP, serum ferritin and inflammatory cytokines as appropriate |

[1]**Ordinal Scale (GSK modified version, adapted from WHO, 2020 version)**

CCI - This section contained Clinical Outcome Assessment data collection questionnaires or indices, which are protected by third party copyright laws and therefore have been excluded.

[redacted]

The primary estimand is the odds ratio between otilimab versus placebo in the proportion of participants alive and free of respiratory failure at Day 28 in participants with severe pulmonary COVID-19 related disease regardless of use of additional medications or changes to standard of care.

### 4. STUDY DESIGN

#### 4.1. Overall Design

This study is a multi-center, randomized, double-blind, placebo-controlled trial to assess the efficacy and safety of otilimab for the treatment of severe pulmonary COVID-19 related disease. The study population consists of hospitalized participants with new onset hypoxia requiring significant oxygen support or requiring invasive mechanical ventilation (≤48 hours before dosing). All participants will receive standard of care as per institutional protocols, in addition to study treatment. Use of drugs or treatments specifically intended for COVID-19 related disease are permitted only if according to local hospital/institutional policies, and if not excluded in Section 5.2 Exclusion Criteria Prior/Concomitant therapies. Any concerns related to eligibility in respect of medications or treatments taken for COVID-19 related disease should be discussed with the GSK medical monitor(s). See Section 6.6.

After completion of screening assessments, participants will be treated with a single IV dose of otilimab 90 mg on Day 1, assessed daily while in hospital until Discharge (or Day 28, whichever is sooner), and followed up at Days 42 and 60 after randomization.

Participants will be recruited from two mutually-exclusive categories within the ordinal scale:

CCI - This section contained Clinical Outcome Assessment data collection questionnaires or indices, which are protected by third party copyright laws and therefore have been excluded.

[redacted]

Participants will initially be randomized 1:1 by interactive response technology (IRT) in a blinded manner to receive either a 1-hour infusion of otilimab 90 mg or placebo IV in addition to standard of care.

##### 4.1.1. Initial Safety Cohort (first 20 participants)

The first 20 participants will be from category 5 above (hospitalized, high-flow oxygen (≥15L/min), CPAP, BiPAP, non-invasive ventilation). Since this is the first time that otilimab has been tested in participants with severe pulmonary COVID-19 related disease, initial dosing will be staggered, and safety data reviewed as follows:

- The first two participants to be dosed will be randomized 1:1 (otilimab:placebo) and monitored closely. After approximately 24 hours, the clinical status of both participants will be assessed by the investigator(s) and then discussed with the GSK medical monitor. If no safety issues are identified, and the GSK medical monitor and investigator(s) agree, two further participants will be allowed to be dosed.
- The third and fourth participants will also be randomized 1:1 (otilimab:placebo) and monitored closely. After approximately 24 hours, the clinical status of all four participants will be assessed by the investigator(s) and then discussed with the GSK medical monitor. If no safety issues are identified, and the GSK medical monitor and investigator(s) agree, recruitment and dosing of further participants will start.
- The GSK Safety Review Team (SRT) will review available blinded safety data on a weekly basis for the additional 16 participants in the initial safety cohort.
- Safety data will be reviewed by the Independent Data Monitoring Committee (IDMC), as outlined in the IDMC charter (refer to Section 9.1.5 for more information about the IDMC).

Unless a safety signal is identified during the ongoing safety review by the GSK SRT and evaluation by the IDMC, enrolment of the main cohort of participants (both categories 5 and 6) will be initiated.

##### 4.1.2. Main Cohort (after 20 participants)

After the initial safety cohort, enrolment rate will initially be capped to control randomization. For the first 200 participants, a weekly maximum randomization cap will be set using the study IRT system, as follows:

- Participant 21 to 100 – randomization cap of 20 participants / week
- Participant 101 to 200 – randomization cap of 50 participants / week

After 200 participants, it is planned that the randomization cap will be removed. The randomization caps will be subject to revision by the IDMC at the scheduled data review meetings (as outlined in the IDMC Charter).
**Stratification** – based on the study eligibility criteria, the clinical status of the participants entering the main cohort (*i.e.* after the 20th participant) of the study will be within the two ordinal scale categories 5 and 6 and also age groups, as follows:

- Hospitalized, high-flow oxygen (≥15L/min), CPAP, BiPAP, non-invasive ventilation. Age <60 years
- Hospitalized, high-flow oxygen (≥15L/min), CPAP, BiPAP, non-invasive ventilation. Age 60 to <70 years
- Hospitalized, high-flow oxygen (≥15L/min), CPAP, BiPAP, non-invasive ventilation. Age 70 to <80 years
- Hospitalized, intubation and mechanical ventilation. Age <60 years
- Hospitalized, intubation and mechanical ventilation. Age 60 to <70 years
- Hospitalized, intubation and mechanical ventilation. Age 70 to <80 years

The rationale for these stratification factors is to obtain a balanced population across the treatment arms with respect to the severity of the disease and age group which are both associated with increased mortality [Grasselli, 2020; Wu, 2020].

Further, the target percentage of participants in category 6 (Hospitalized, intubation and mechanical ventilation) will be no more than 60% of the study population. However, this target may be revised based on ongoing IDMC reviews (see Section 9.1.5).

#### 4.2. Scientific Rationale for Study Design

A multicenter, randomized, double-blind, placebo-controlled trial is a well-established strategy to evaluate efficacy and safety of investigational medicinal products.

The study will utilize a group sequential design with interim analyses that intends to identify early signs of clinical efficacy if responses are higher than expected and to avoid the use of ineffective therapeutics in severely or critically ill participants with severe pulmonary COVID-19 related disease. In addition, frequent in-stream reviews of safety data will be performed to identify early signs of harm and stop the study if these are identified. This design allows for the assessment of efficacy and safety as well as evaluating the benefit:risk.

#### 4.3. Justification for Dose

##### 4.3.1. Otilimab

CCI
[redacted]

CCI

[redacted]

###### 4.3.2. Placebo

There is no established treatment for severe pulmonary COVID-19 related disease, hence a placebo control is required to distinguish the safety and tolerability of otilimab from the background signs and symptoms of severe pulmonary COVID-19 related disease and to evaluate its potential benefit on clinical outcomes.

#### 4.4. End of Study Definition

A participant is considered to have completed the study if he/she has completed all visits of the study through to Day 60 or has died prior to Day 60. The end of the study is defined as the date of the last contact of the last participant in the study.

### 5. STUDY POPULATION

Prospective approval of protocol deviations to recruitment and enrolment criteria, also known as protocol waivers or exemptions, is not permitted.

#### 5.1. Inclusion Criteria

Participants are eligible to be included in the study only if all of the following criteria apply:

| **AGE** |
|---|
| 1. Age ≥18 years and ≤79 years at the time of obtaining informed consent. |
| **TYPE OF PARTICIPANT AND DISEASE CHARACTERISTICS** |
| 2. Participants must:<br>a. have positive SARS-CoV-2 result (any validated test*, e.g.* RT-PCR [performed on an appropriate specimen; *e.g.* respiratory tract sample])<br>b. **AND** be hospitalized due to diagnosis of pneumonia (chest X-ray or computerized tomography [CT] scan consistent with COVID-19)<br>c. **AND** be developing new onset of oxygenation impairment requiring any of the following:<br>1. high-flow oxygen (≥15L/min)<br>2. non-invasive ventilation (*e.g.* CPAP, BiPAP)<br>3. mechanical ventilation ≤48h prior to dose<br>d. **AND** have increased biological markers of systemic inflammation (either CRP >ULN[1] or serum ferritin >ULN[1]). |

[1]According to local reference range. Repeat CRP or ferritin tests are permitted.

| **SEX** |
|---|
| 3. No gender restriction.<br>4. Female participants must meet and agree to abide by the contraceptive criteria detailed in Appendix 4. Contraceptive use by women should be consistent with local regulations regarding the methods of contraception for those participating in clinical studies.<br>• A female participant is eligible to participate if she is not pregnant or breastfeeding, and one of the following conditions applies:<br>○ Is a woman of non-childbearing potential (WONCBP) as defined in Section 9.4: Contraceptive and Barrier Guidance.<br>OR<br>○ Is a woman of childbearing potential (WOCBP) and using a contraceptive method that is highly effective, with a failure rate of <1%, as described in Section 9.4 during the study intervention period and for at least 60 days after the last dose of study intervention (sexual abstinence is acceptable if it is the participant's normal practice). |

| |
|---|
| ○ If not consistently on a highly effective method of contraception (Section 9.4) during hospitalization, the participant must agree to a highly effective contraception plan if discharged before Day 60.<br>○ The investigator should evaluate potential for contraceptive method failure (*e.g.* noncompliance, recently initiated) in relation to the first dose of study intervention.<br>○ A WOCBP must have a negative highly sensitive pregnancy test (urine or serum as required by local regulations) at hospital admission or before the first dose of study intervention. See Section 7.3.5 Pregnancy Testing (additional requirements for pregnancy testing during and after study intervention).<br>○ The investigator is responsible for review of medical history, menstrual history, and recent sexual activity to decrease the risk for inclusion of a woman with an early undetected pregnancy. |

| **INFORMED CONSENT** |
|---|
| 5. Capable of giving written informed consent as described in Section 9.1.3. If participants are not capable of giving written informed consent, alternative consent procedures will be followed as detailed in Section 9.1.3. |

#### 5.2. Exclusion Criteria

Participants are excluded from the study if any of the following criteria apply:

| RELATED TO SEVERE PULMONARY COVID-19 RELATED DISEASE |
|---|
| 1. Progression to death is imminent and inevitable within the next 48 hours, irrespective of the provision of treatments, in the opinion of the investigator.<br>2. Multiple organ failure according to the investigator's judgement or a Sequential Organ Failure assessment (SOFA score) >10 if in the ICU.<br>3. Extracorporeal membrane oxygenation (ECMO) hemofiltration/dialysis, or high-dose (>0.15µg/kg/min) noradrenaline (or equivalent) or more than one vasopressor. |

| CONCOMITANT MEDICAL CONDITIONS |
|---|
| 4. Current serious or uncontrolled medical condition (*e.g.* significant pulmonary disease [such as severe COPD or pulmonary fibrosis], heart failure [NYHA class III or higher], significant renal dysfunction, acute myocardial infarction or acute cerebrovascular accident within the last 3 months) or abnormality of clinical laboratory tests that, in the investigator's judgment, precludes the participant's safe participation in and completion of the study.<br>5. **Untreated** systemic bacterial, fungal, viral, or other infection (other than SARS-CoV-2).<br>6. Known active tuberculosis (TB), history of untreated or incompletely treated active or latent TB, suspected or known extrapulmonary TB.<br>7. Known HIV regardless of immunological status.<br>8. Known HBsAg and/or anti-HCV positive.<br>9. Currently receiving radiotherapy, chemotherapy or immunotherapy for malignancy. |

| PRIOR MEDICATIONS/TREATMENTS |
|---|
| 10. Received monoclonal antibody therapy (*e.g.* tocilizumab, sarilumab) within the past 3 months prior to randomization, including intravenous immunoglobulin, or planned to be received during the study.<br>11. Received immunosuppressant therapy including but not limited to cyclosporin, azathioprine, tacrolimus, mycophenolate, JAK inhibitors (*e.g.* baricitinib, tofacitinib, upadacitinib) within the last 3 months prior to randomization or planned to be received during the study.<br>Note: Participants with an organ transplant are therefore excluded (except patients with corneal transplants not requiring immunosuppression).<br>12. History of allergic reaction, including anaphylaxis to any previous treatment with an anti-GM-CSF therapy.<br>13. Received COVID-19 convalescent plasma within 48 hours of randomization.<br>Note: Participants who have received COVID-19 convalescent plasma but continue to worsen in the 48 hours after infusion of the convalescent plasma, in the opinion of the investigator, will become eligible for the study. |

| PROHIBITED MEDICATIONS |
|---|
| 14. Currently receiving chronic oral corticosteroids for a non-COVID-19 related condition in a dose higher than prednisone 10 mg or equivalent per day. |
| 15. Treatment with an investigational drug within 30 days of randomization. |

| PRIOR/CONCURRENT CLINICAL STUDY EXPERIENCE |
|---|
| 16. Participating in other drug clinical trials, including for COVID-19. |

| DIAGNOSTIC ASSESSMENTS |
|---|
| 17. Aspartate aminotransferase (AST) or alanine aminotransferase (ALT) >5x upper limit of normal (ULN). |
| 18. Platelets <50,000/$mm^{3.}$ |
| 19. Hemoglobin ≤9 g/dL. |
| 20. Absolute neutrophil count (ANC) <1.5 x $10^9$/L (neutropenia ≥ Grade 2). |
| 21. Estimated GFR ≤30 mL/min/1.73$m^2$. |

| OTHER CRITERIA |
|---|
| 22. Pregnant or breastfeeding females. |

#### 5.3. Screen Failures

Screen failures are defined as participants who consent to participate in the clinical study but are not subsequently randomized. A minimal set of screen failure information is required to ensure transparent reporting of screen failure participants to meet the Consolidated Standards of Reporting Trials (CONSORT) publishing requirements and to respond to queries from regulatory authorities. Minimal information includes demography, screen failure details, eligibility criteria, any protocol deviations and any protocol deviations and any serious adverse events (SAEs).

Individuals who do not meet the criteria for participation in this study may be rescreened once. Rescreened participants should be assigned a new participant number for every screening/rescreening event.

### 6. STUDY INTERVENTION(S) AND CONCOMITANT THERAPY

Study intervention is defined as any investigational intervention(s) or placebo intended to be administered to a study participant according to the study protocol.

#### 6.1. Study Intervention(s) Administered

CCI [redacted]

CCI

[redacted]

#### 6.2. Preparation/Handling/Storage/Accountability

- The site unblinded pharmacist must confirm appropriate temperature conditions have been maintained during transit for all study intervention received and any discrepancies are reported and resolved before use of the study intervention.
- Only participants enrolled in the study may receive study intervention. All study interventions must be stored in a secure, environmentally controlled, and monitored (manual or automated) area in accordance with the labelled storage conditions with access limited to the investigator and authorized site staff.
- The investigator, institution, or the head of the medical institution (where applicable) is responsible for study intervention accountability, reconciliation, and record maintenance (*i.e.* receipt, reconciliation, and final disposition records).
- Further guidance and information for the final disposition of unused study intervention are provided in the SRM.
- Under normal conditions of handling and administration, study intervention is not expected to pose significant safety risks to site staff. Take adequate precautions to avoid direct eye or skin contact and the generation of aerosols or mists. In the case of unintentional occupational exposure notify the monitor, Medical Monitor and/or GSK study contact.
- A Material Safety Data Sheet (MSDS)/equivalent document describing occupational hazards and recommended handling precautions either will be provided to the investigator, where this is required by local laws, or is available upon request from GSK.

#### 6.3. Measures to Minimize Bias: Randomization and Blinding

All participants will be centrally randomized using IRT. Before the study is initiated, the log in information and directions for the IRT will be provided to each site. Study intervention will be administered at the time-point detailed in the SoA (Section 1.3). Returned study intervention must not be re-dispensed.

The IRT will be programmed with blind-breaking instructions. In case of an emergency, the investigator has the sole responsibility for determining if unblinding of a participant’s intervention assignment is warranted. Participant safety must always be the first consideration in making such a determination. If a participant’s intervention assignment is unblinded GSK must be notified within 24 hours after breaking the blind. The date and reason that the blind was broken must be recorded in the source documentation and case report form, as applicable.

Participants will be randomized in a ratio of 1:1 to receive study intervention (Section 1.2, Section 4.1 and Section 6.1). Investigators will remain blinded to each participant’s assigned study intervention throughout the course of the study. To maintain this blind, all participants will receive a single IV injection. An unblinded pharmacist (or unblinded designee) will be responsible for the dispensation of the study intervention and will endeavor to ensure that there are no differences in time taken to dispense between the different intervention arms. The IV bag should be prepared away from view of the blinded site staff and the participant. Every IV injection must be labelled to maintain the blind (refer to SRM).

Unblinded monitors and in the event of a Quality Assurance audit, the auditor(s) will be allowed access to unblinded study intervention records at the site(s) to verify that randomization/dispensing has been done accurately.

A participant may continue in the study if that participant's intervention assignment is unblinded.

GSK's Pharma Safety staff may unblind the intervention assignment for any participant with an SAE. If the SAE requires that an expedited regulatory report be sent to one or more regulatory agencies, a copy of the report, identifying the participant's intervention assignment, may be sent to investigators in accordance with local regulations and/or GSK policy.

#### 6.4. Study Intervention Compliance

Participants will receive study intervention directly from the investigator or designee, under medical supervision. The date and time of the dose administered will be recorded in the source documents. The dose of study intervention and study participant identification will be confirmed at the time of dosing by a member of the study site staff other than the person administering the study intervention.

#### 6.5. Treatment of Overdose

For this study, any dose of otilimab greater than the 90 mg dose indicated in this study or used more frequently than permitted (otilimab 90 mg single IV infusion) will be considered an overdose.

No specific treatment is recommended for an overdose of otilimab, and the investigator should treat as clinically indicated.

In the event of a potential overdose, the investigator should:

- Contact the Medical Monitor immediately.
- Closely monitor the participant for adverse event (AE)/serious adverse event (SAE) and laboratory abnormalities.
- Contact the unblinded monitor with the quantity and duration of the overdose to be documented.

#### 6.6. Concomitant Therapy

At Screening, information will be collected on any medications the participant has taken in the 14 days before screening.

Any medication or vaccine (including over-the-counter or prescription medicines, vitamins, and/or herbal supplements) that the participant is receiving at the time of enrolment or receives during the study must be recorded along with:

- reason for use
- dates of administration including start and end dates
- dosage information including dose and frequency

The Medical Monitor should be contacted if there are any questions regarding concomitant or prior therapy.

Participants who have received COVID-19 convalescent plasma but continue to worsen in the 48 hours after infusion of the convalescent plasma, in the opinion of the investigator, will become eligible for the study.

Due to the effect of cytokines on the CYP450 enzymes (Section 2.3) initiation or discontinuation of study intervention may have a clinically relevant effect for CYP substrates with a narrow therapeutic index *e.g.* warfarin and theophylline should be monitored.

Investigators should exercise caution when study intervention is co-administered with CYP3A4 substrate drugs where decrease in effectiveness is undesirable, *e.g.*, oral contraceptives, lovastatin, atorvastatin, *etc.*

Aside from those medications specified in Exclusion Criteria (Section 5.2) or Section 6.6.1, all regular medications that are required by the participant will be allowed. All concomitant medications (and treatments, such as convalescent plasma) should be recorded in the eCRF.

##### 6.6.1. Medication NOT Permitted During the Study

Any monoclonal antibody therapy (*e.g.* tocilizumab, sarilumab), including intravenous immunoglobulin.

Immunosuppressant therapy including but not limited to cyclosporin, azathioprine, tacrolimus, mycophenolate, JAK inhibitors (*e.g.* baricitinib, tofacitinib, upadacitinib).

Chronic oral corticosteroids (for a non-COVID-19 related condition) in a dose higher than prednisone 10 mg or equivalent per day.

##### 6.6.2. Medication/Treatment PERMITTED During the Study

The participant's regular medication.

Treatment required for standard of care (*e.g.* analgesia, antipyretic, sedation).

Medications/treatments required for ICU care.

Additional medications/treatments of COVID-19 related disease (*e.g.* remdesivir, favipiravir, convalescent plasma) are permitted only if according to local hospital/institutional policies, and not part of a clinical trial. Any concerns regarding the acceptability of potential treatments should be discussed with the GSK medical monitor(s).

#### 6.7. Participant Discontinuation/Withdrawal from the Study

- A participant may withdraw from the study at any time at his/her own request, at the request of their legally authorized representative (LAR) or may be withdrawn at any time at the discretion of the investigator for safety, behavioral, or compliance reasons. This is expected to be uncommon.
- At the time of discontinuing from the study, if possible, a Withdrawal visit (same as Discharge visit) should be conducted (for hospitalized participants) or a Withdrawal phone call (same as Follow-up phone call) should be conducted (for non-hospitalized participants), as shown in the SoA.
- If the participant withdraws consent or the LAR requests that the participant is withdrawn for disclosure of future information, the sponsor may retain and continue to use any data collected before such a withdrawal of consent.
- If a participant withdraws from the study or the LAR requests that the participant is withdrawn, he/she may request destruction of any samples taken and not tested, and the investigator must document this in the site study records.

#### 6.8. Lost to Follow Up

A participant will be considered lost to follow-up if he or she is unable to be contacted by the study site after discharge from hospital. Every effort must be made to reduce missing data.

- The site must attempt to contact the participant as soon as possible and counsel the participant on the importance of maintaining the assigned study schedule and ascertain whether or not the participant wishes to and/or should continue in the study.
- Before a participant is deemed lost to follow up, the investigator or designee must make every effort to regain contact with the participant (where possible, 3 telephone calls and, if necessary, a certified letter to the participant’s last known mailing address or local equivalent methods). These contact attempts should be documented in the participant’s medical record.
- Should the participant continue to be unreachable, he/she will be considered to have withdrawn from the study.

Discontinuation of specific sites or of the study as a whole is described in Section 9.1.9.

### 7. STUDY ASSESSMENTS AND PROCEDURES

This section lists the parameters of each planned study assessment. Participants will mainly be evaluated using standard of care investigations. In addition to standard care, data will be collected for the Ordinal Scale and the Daily Clinical Features of COVID-19, and blood samples will be collected for PK and PD analysis (collected as detailed in the SoA, Section 1.3).

- Study procedures and their timing are summarized in the SoA (Section 1.3).
- As detailed in the SoA (Section 1.3), follow-up phone calls must be made within ±2 days of the scheduled time-point, but every effort must be made to complete the phone call on the scheduled day. This is particularly important if the Day 28 time-point is a phone call, because this is the study primary endpoint.
- Every effort must be made to reduce missing data throughout the study.
- Protocol waivers or exemptions are not allowed.
- Urgent study-related safety concerns should be discussed with the GSK medical monitor immediately upon occurrence or awareness to determine if the participant should continue or in exceptional circumstances withdraw from the study.
- Adherence to the study design requirements, including those specified in the SoA, is essential and required for study conduct.
- All screening evaluations must be completed and reviewed to confirm that potential participants meet all eligibility criteria. The investigator will maintain a screening log to record details of all participants screened and to confirm eligibility or record reasons for screening failure, as applicable.
- Procedures conducted as part of the participant's routine clinical management (*e.g.*, blood count) and taken before obtaining consent may be utilized for screening or baseline purposes provided the procedure met the protocol-specified criteria and was performed within the time frame defined in the SoA.
- The maximum amount of blood collected from each participant over the duration of the study for the purpose of meeting the objectives will not exceed 200 mL. This does not include any standard of care assessments.
- Repeat or unscheduled samples may be taken for safety reasons or for technical issues with the samples.

#### 7.1. Entry/Screening Assessments

##### 7.1.1. Demography & Medical History

The following demographic parameters will be captured: year of birth, gender and ethnicity/race. If it is not possible to collect at Screening, demography can be collected at any point during the study.

We are collecting details of ethnicity in this study since there have been reports from several counties that ethnic minority groups have been disproportionately affected by either the incidence or outcomes of COVID-19 related disease. In line with ICH E5 principles, this information is therefore paramount to the assessment of the ethnic sensitivity of otilimab in this patient population.

To the extent practical given the participant's condition, a full medical history, including cardiovascular risk, will be taken during Screening or as information becomes available at any point during the study. This should record concurrent medical conditions and concomitant medications.

At Screening, information will be collected on any medications the participant has taken in the 14 days before Screening.

##### 7.1.2. Chest X-ray or CT Scan

A chest X-ray or computerized tomography (CT) scan must be available at Screening to confirm a diagnosis of pneumonia and eligibility. If another chest X-ray or CT scan is done after Screening and before Discharge, this status should be recorded in the eCRF.

#### 7.2. Efficacy Assessments

**General Note**: Research staff are asked to liaise with clinical staff managing the participant to ensure that all study relevant data is collected efficiently and accurately and that study related assessments are not performed soon after or before those required for participant clinical management. Hence, all study mandated physical examination assessments, measurements of vital signs and obtaining blood for laboratory tests / point of care tests should be performed in coordination with the clinical staff looking after the participant.

##### 7.2.1. Daily clinical features of COVID-19

Signs and symptoms of COVID-19 will be collected for non-mechanically ventilated participants. A detailed list of signs and symptoms of COVID-19 at admission will be recorded at Screening, and an abbreviated list of signs and symptoms of COVID-19 will be reviewed daily (as outlined in the eCRF).

##### 7.2.2. $SpO_2$ (for participants not on invasive mechanical ventilation)

$SpO_2$ should be measured at the timepoints defined in the SoA (Section 1.3). Where possible, this should be measured "at rest" *i.e.* not undergone a procedure or handling for at least 5 minutes.

##### 7.2.3. Concentration of inspired oxygen ($FiO_2$)

Concentration of inspired oxygen ($FiO_2$) and/or oxygen flow rate (L/min) should be measured at the timepoints defined in the SoA (Section 1.3).

##### 7.2.4. Ventilation status

Ventilation status (invasive or non-invasive ventilation), mask or nasal catheter oxygen will be collected at times outlined in the SoA (Section 1.3). For mechanically ventilated participants, indicate if the participant was in the prone or semi-prone position at any time whilst mechanically ventilated. If the participant is discharged from hospital with oxygen support, record oxygen treatment at subsequent phone call follow-ups (at times outlined in the SoA (Section 1.3)).

##### 7.2.5. ICU Supportive Care

For participants in ICU, additional information will be collected on the use of supportive care: inotropes/vasopressors, renal replacement therapy (RRT) and extracorporeal membrane oxygenation (ECMO) should be recorded at the timepoints defined in the SoA (Section 1.3).

##### 7.2.6. Ordinal Scale

The mutually exclusive ordinal scale presented in Table 2, adapted from the WHO scale 2020, will be used to assess clinical improvement at the timepoints indicated in the SoA (Section 1.3).

**Table 2 Ordinal Scale (GSK modified version, adapted from WHO, 2020 version)**

CCI - This section contained Clinical Outcome Assessment data collection questionnaires or indices, which are protected by third party copyright laws and therefore have been excluded.

[redacted]

##### 7.2.7. Sequential Organ Failure Assessment (SOFA) Scores

The SOFA score is a tool for grading organ dysfunction in critically ill participants during their admission to ICU. Scores range from 0 to 4 for each component and represent the extent of dysfunction or failure [Vincent, 1996].

The SOFA score is made up of the following components:

- The Respiratory System component of the SOFA score will be assessed using $PaO_2/FiO_2$ (mmHg) and will require an arterial blood gas measurement.
- The Nervous System component of the SOFA score will be determined using the Glasgow Coma Scale (GCS). If possible, GCS should be assessed / completed during a sedation holiday. If not, then the highest score of the last 24-hour period should be used. For participants who are intubated and ventilated, the GCS can be omitted from the SOFA score calculation.

- The Cardiovascular System component of the SOFA score can be determined by Mean Arterial Pressure (MAP) measurement or administration of vasopressors.
- The Liver, Renal, and Coagulation components of the SOFA score will be determined by measurement of total bilirubin, serum creatinine, and platelet count respectively. The highest score of the last 24-hour period for each measurement should be used.

The individual parameters will not be captured in the eCRF (unless they are captured for another reason) – only the overall score calculated at the site will be recorded.

SOFA scores will be determined at the time points stipulated in SoA (Section 1.3).

#### 7.3. Safety Assessments

Planned time points for all safety assessments are provided in the SoA (Section 1.3) – where possible, these should be aligned with standard of care.

##### 7.3.1. Physical Examinations

- A full physical examination will include, at a minimum, assessments of the Cardiovascular, Respiratory, Gastrointestinal and Neurological systems. Results will not be recorded in the eCRF.

##### 7.3.2. Vital Signs

- Temperature, pulse rate, respiratory rate, blood pressure will be assessed. Single measurements will be used at all timepoints as detailed in the SoA.
- At Screening only, body weight should either be measured, established from the medical records *e.g.* last 3 months and /or reported / estimated at the bedside.
- The method used to measure temperature will be as per standard of care but must be recorded in the eCRF.

##### 7.3.3. Electrocardiograms

- Single 12-lead ECG will be obtained as outlined in the SoA (see Section 1.3).
- Each ECG report must be reviewed by the Principal Investigator (PI) or his qualified designee. Each ECG must be initialled and dated by the PI or his/her qualified designee reviewing the report.

##### 7.3.4. Clinical Safety Laboratory Assessments

- See Appendix 2 for the list of clinical laboratory tests to be performed and to the SoA for the timing and frequency.
- The investigator must review the laboratory report, document this review, and record any clinically significant changes occurring during the study as an AE. The laboratory reports must be filed with the source documents.

- Abnormal laboratory findings associated with the underlying disease are not considered clinically significant, unless judged by the investigator to be more severe than expected for the participant's condition.
- All laboratory tests with values considered clinically significantly abnormal (during participation in the study until the follow up visit) should be repeated until the values return to normal or baseline or are no longer considered significantly abnormal by the investigator or medical monitor. If clinically significant values do not return to normal/baseline within a period of time judged reasonable by the investigator, the etiology should be identified and the sponsor notified.
- If laboratory values from non-protocol specified laboratory tests are performed and require a change in participant management or are considered clinically significant by the investigator (*e.g*. SAE), then the results must be recorded.

##### 7.3.5. Pregnancy Testing

- Pregnancy testing (urine or serum as required by local regulations) should be conducted at Screening to confirm eligibility as per Section 5.1.
- Pregnancy testing (urine or serum as required by local regulations) should be conducted at Discharge. If the participant has been discharged before Day 60, the Day 60 phone call will include a verbal confirmation about pregnancy status.
- Additional serum or urine pregnancy tests may be performed, as determined necessary by the investigator or required by local regulation, to establish the absence of pregnancy at any time during the participant's time in the study.

#### 7.4. Adverse Events (AEs), Serious Adverse Events (SAEs) and Other Safety Reporting

The definitions of AEs or SAEs can be found in Section 9.3.

AEs will be reported by the participant (or, when appropriate, by a caregiver, surrogate, or the participant's legally authorized representative).

The investigator and any qualified designees are responsible for detecting, documenting, and reporting events that meet the definition of an AE or SAE and remain responsible for following up all AEs that are serious, considered related to the study intervention or the study, or that caused the participant to discontinue the study.

The method of recording, evaluating, and assessing causality of AEs and SAEs and the procedures for completing and transmitting SAE reports are provided in Section 9.3.

##### 7.4.1. Time Period and Frequency for Collecting AE and SAE Information

- All AEs and SAEs will be collected from the start of intervention until the follow-up visit at the time points specified in the SoA (Section 1.3). However, any SAEs assessed as related to study participation (*e.g*., study intervention, protocol-mandated procedures, invasive tests, or change in existing therapy) or related to a GSK product will be recorded from the time a participant consents to participate in the study.

- Medical occurrences that begin before the start of study intervention but after obtaining informed consent will be recorded as Medical History/Current Medical Conditions not as AEs.
- All SAEs will be recorded and reported to the sponsor or designee immediately and under no circumstance should this exceed **24 hours**, as indicated in Appendix 3. The investigator will submit any updated SAE data to the sponsor within **24 hours** of it being available.
- Investigators are not obligated to actively seek information on AEs or SAEs after the conclusion of the study participation. However, if the investigator learns of any SAE, including a death, at any time after a participant has been discharged from the study, and he/she considers the event to be reasonably related to the study intervention or study participation, the investigator must promptly notify the sponsor.

##### 7.4.2. Method of Detecting AEs and SAEs

Care will be taken not to introduce bias when detecting AE and/or SAE. Open-ended and non-leading verbal questioning of the participant is the preferred method to inquire about AE occurrence.

##### 7.4.3. Follow-up of AEs and SAEs

After the initial AE/SAE report, the investigator is required to proactively follow each participant at subsequent visits/contacts. All SAEs, and non-serious AEs of special interest (as defined in Section 7.4.7), will be followed until the event is resolved, stabilized, otherwise explained, or the participant is lost to follow-up (as defined in Section 6.8). Further information on follow-up procedures is given in Appendix 3.

##### 7.4.4. Regulatory Reporting Requirements for SAEs

- Prompt notification by the investigator to the sponsor of an SAE is essential so that legal obligations and ethical responsibilities towards the safety of participants and the safety of a study intervention under clinical investigation are met.
- The sponsor has a legal responsibility to notify both the local regulatory authority and other regulatory agencies about the safety of a study intervention under clinical investigation. The sponsor will comply with country-specific regulatory requirements relating to safety reporting to the regulatory authority, Institutional Review Boards (IRB)/Independent Ethics Committees (IEC), and investigators.
- An investigator who receives an investigator safety report describing an SAE or other specific safety information (*e.g.*, summary or listing of SAEs) from the sponsor will review and then file it along with the Investigator's Brochure and will notify the IRB/IEC, if appropriate according to local requirements.

##### 7.4.5. Pregnancy

- Details of all pregnancies in female participants will be collected after the start of study intervention and until follow-up visit (Day 60).
- If a pregnancy is reported, the investigator will record pregnancy information on the appropriate form and submit it to GSK within **24 hours** of learning of the pregnancy in a female participant. While pregnancy itself is not considered to be an AE or SAE, any pregnancy complication or elective termination of a pregnancy for medical reasons will be reported as an AE or SAE.
- Abnormal pregnancy outcomes (*e.g.*, spontaneous abortion, fetal death, stillbirth, congenital anomalies, ectopic pregnancy) are considered SAEs and will be reported as such.
- The participant will be followed to determine the outcome of the pregnancy. The investigator will collect follow-up information on the participant and the neonate, and the information will be forwarded to the sponsor.
- Any post-study pregnancy-related SAE considered reasonably related to the study intervention by the investigator will be reported to the sponsor as described in Section 7.4.4. While the investigator is not obligated to actively seek this information in former study participants, he or she may learn of an SAE through spontaneous reporting.

##### 7.4.6. Cardiovascular and Death Events

For any cardiovascular events detailed in Appendix 3 and all deaths, whether or not they are considered SAEs, specific Cardiovascular (CV) and Death sections of the eCRF will be required to be completed. These sections include questions regarding cardiovascular (including sudden cardiac death) and non-cardiovascular death.

The CV eCRFs are presented as queries in response to reporting of certain CV MedDRA terms. The CV information should be recorded in the specific cardiovascular section of the eCRF within one week of receipt of a CV Event data query prompting its completion.

The Death eCRF is provided immediately after the occurrence or outcome of death is reported. Initial and follow-up reports regarding death must be completed within one week of when the death is reported.

##### 7.4.7. Adverse Events of Special Interest

The potential risks of otilimab are discussed in Section 2.3.

Adverse events of special interest (AESI) include:

- Cytokine release syndrome (CRS)
- Serious hypersensitivity reactions
- Infusion site reactions
- Neutropenia $\geq$ Grade 3 ($<1.0 \times 10^9$/L)

- Serious infections

###### 7.4.7.1. Cytokine Release Syndrome

Cytokine-associated toxicity, also known as cytokine release syndrome (CRS), is a non–antigen-specific toxicity that occurs as a result of strong immune activation. The magnitude of immune activation typically required to mediate clinical benefit using modern immunotherapies exceeds levels of immune activation that occurs in more natural settings. As immune-based therapies have become more potent, CRS is becoming increasingly recognized.

Symptomatology associated with CRS and the severity of symptoms varies greatly but may be quite mild, and management can be complicated by inter-current conditions in these participants. This may be particularly the case in patients with severe pulmonary COVID-19 related disease.

Potentially life-threatening severe complications of CRS include cardiac dysfunction, acute respiratory distress syndrome, neurologic toxicity, renal and/or hepatic failure, and disseminated intravascular coagulation. Of particular concern is cardiac dysfunction, which can be rapid onset and severe, but is typically reversible.

In the event of CRS, investigators should follow institutional guidelines for symptom management. Additional guidance can be found in the published literature [Lee, 2014].

For this study, CRS is defined as a worsening of the CRS Grading System (below) **within 6 hours** after study drug infusion.

In the event that CRS is suspected (defined as a worsening of the CRS Grading System within 6 hours of drug administration) and is thought to be caused by otilimab by the investigator, additional blood samples should be taken (see Section 1.3 and Section 9.2). These blood samples should be taken as soon as possible after suspected CRS onset and then again 24 hours later. The following blood samples should be taken:

- PD (cytokine) sample
- Hematology
- CRP
- D-dimer
- Ferritin

In the event that new onset of CRS is suspected, the medical monitor must be notified immediately and under no circumstance can this exceed 24 hours, and the event must also be reported within 24 hours as an AE or an SAE. The investigator will submit any updated data related to the CRS event (including completion of any AE or SAE CRF forms) to the sponsor within 24 hours of it being available. Additional CRF forms related to the CRS event should then be completed as soon as possible.

**CRS Grading System**[1]

| Grade | Toxicity |
|---|---|

CCI - This section contained Clinical Outcome Assessment data collection questionnaires or indices, which are protected by third party copyright laws and therefore have been excluded.

[redacted]

[1]Adapted from Lee, 2014
[2]Grades CCI [redacted] refer to CTCAE v5.0 grading

#### 7.5. Pharmacokinetics

- Blood samples (approximately 3.5 mL) will be collected for measurement of serum concentrations of otilimab as specified in the SoA (Section 1.3).
- The actual date and time (24-hour clock time) of each blood sample collection will be recorded.
- Collection, processing, storage and shipping procedures are provided in the central laboratory manual.
- Intervention concentration information that may/would unblind the study will not be reported to investigative sites or blinded personnel until the study has been unblinded.

#### 7.6. Genetics

Genetics are not evaluated in this study.

#### 7.7. Pharmacodynamics

- Blood samples will be collected to help understand the disease, otilimab target levels and response to otilimab in COVID-19 related disease, as specified in the SoA (Section 1.3).
- Biomarkers may include, but are not limited to: inflammatory cytokines, such as IL-6; growth factors, such as GM-CSF; and other molecules that are indicative of hyper-inflammation.

- The actual date and time (24-hour clock time) of each blood sample collection will be recorded.
- Collection, processing, storage and shipping procedures are provided in the central laboratory manual.

GSK may store samples for up to 15 years after the end of the study to achieve study objectives. Additionally, with participants' consent, samples may be used for further research by GSK or others such as universities or other companies to contribute to the understanding of SARS-CoV-2 infection, associated COVID-19 disease or other diseases, the development of related or new treatments or research methods.

#### 7.8. Immunogenicity Assessments

Antibodies to otilimab will not be assessed as part of this study, since this is a single dose study.

#### 7.9. Health Economics/Medical Resource Utilization and Health Economics

Health Economics/Medical Resource Utilization and Health Economics parameters will not be evaluated in this study.

### 8. STATISTICAL CONSIDERATIONS

#### 8.1. Statistical Hypotheses

The primary objective of this study is to compare the efficacy of otilimab 90 mg IV versus placebo in participants with severe pulmonary COVID-19 related disease.

The study will test the null hypothesis that the difference between otilimab 90 mg and placebo on the proportion of participants alive and free of respiratory failure at Day 28 is less than or equal to zero versus the alternative hypothesis that otilimab 90 mg increases the proportion of participants alive and free of respiratory failure compared with placebo at Day 28.

#### 8.2. Sample Size Determination

Otilimab and placebo will be randomized in a 1:1 ratio.

The study will utilize a group sequential design, using a Lan-DeMets alpha-spending function to control the type I error with four interim analyses for futility, two early in the study where futility alone will be assessed, using Pocock analogue rules; and two for efficacy, at later times in the study and at the same time as futility, using the O'Brien-Fleming analogue rules [Lan, 1983]. A sample size of 800 participants will provide approximately 90% power to detect a difference of 12% in the proportion of participants alive and free of respiratory failure at a one-sided 2.5% significance level and an assumed placebo response rate of 45%. The minimal detectable effect for this design is 7%.

Full details of the decision criteria and interim analysis plan will be given in the IDMC charter and analysis plan.

The assumed placebo response is based on data from recent publications of studies of patients with severe pulmonary COVID-19 related disease [Sanofi, 2020].

#### 8.3. Analysis Sets

The primary population will be the modified intent to treat (MITT) population, defined as all participants who were randomized and received study drug. This population will be used for all data summaries and analyses. Any participant who receives study intervention will be considered evaluable.

Further details of analysis sets will be provided in the Analysis Plan.

#### 8.4. Statistical Analyses

Full details of all analyses will be described in the Analysis Plan.

This section is a summary of the planned statistical analyses for the primary endpoint.

##### 8.4.1. General Considerations

In the case of a difference between the stratification assigned at the time of randomization and the data collected in the eCRF, the analyses will be performed based on the calculated stratum at randomization. All analyses will be adjusted for ordinal scale category at baseline (category 5 or 6) and age group (treated as categorical variable defined by the levels <60 years, 60 to <70 years, 70 to <80 years). Confidence intervals will use the 95% levels unless otherwise specified.

##### 8.4.2. Primary Endpoint(s)

The primary endpoint is whether or not a participant is alive and free of respiratory failure at Day 28.

Participants are considered alive and free of respiratory failure if they are in category 1, 2, 3 or 4 from the GlaxoSmithKline (GSK) modified version ordinal scale adapted from World Health Organization (WHO) scale 2020 (Section 7.2.6. Table 2 - Ordinal Scale).

The primary estimand is the odds ratio between otilimab versus placebo in the proportion of participants alive and free of respiratory failure at Day 28 in participants with severe pulmonary COVID-19 related disease regardless of use of additional medications or changes to standard of care. No other intercurrent events are expected and the strategy for analysis will be to use the treatment policy strategy, with data analyzed as collected. It is expected missing data will be minimal and most will be lost to follow up data after discharge and this along with intermittent data will be imputed using multiple imputation under the missing at random assumption. Every effort will be made to reduce missing data. Supplemental estimand strategies and missing data sensitivities may be performed and further details will be provided in the analysis plan.

The primary endpoint will be summarized using counts and proportions of the number of participants alive and free of respiratory failure and will be analyzed using a logistic regression model, adjusting for ordinal scale category at baseline and age group. The outcome of the model is an odds ratio, and a confidence interval for the odds of improvement in proportion of participants alive and free of respiratory failure at Day 28.

The analysis of the primary endpoint at the in-stream interim analyses will be the same as that described for the primary endpoint above.

##### 8.4.3. Secondary Endpoint(s)

Full details of all analysis methods for the secondary endpoints will be provided in the analysis plan.

##### 8.4.4. Exploratory Endpoint(s)

Full details of all analysis methods for the exploratory endpoints will be provided in the analysis plan.

##### 8.4.5. Safety Analysis

For safety data, no formal hypotheses will be tested. Occurrence of AEs, SAEs, and AESIs, including laboratory tests, vital signs, and 12-lead ECGs will be displayed in the form of listings, frequencies, summary statistics, graphs, and statistical analyses where appropriate. Interpretation will be aided by clinical expertise. Full details, including example outputs, will be documented in the analysis plan.

#### 8.5. Interim Analysis

Interim analyses will be used to assess safety, futility and efficacy. Safety stopping rules will be put in place for the IDMC to stop the study if there is an excess risk of mortality. Full details of timing of analyses and all stopping criteria will be given in the IDMC charter.

An IDMC will actively monitor in-stream interim unblinded safety and efficacy data to make recommendations to GSK as per IDMC charter. The IDMC members will include 4-6 physicians with relevant medical specialist training and one statistician. Details regarding the IDMC process will be available in relevant IDMC documents prior to the first participant's visit.

A GSK Safety Review Team (SRT) will review the blinded safety data of this study at regular intervals through both initial safety and main cohort dosing. Details regarding the SRT process will be available in relevant SRT documents. The SRT will inform the IDMC if any safety signals are identified.

### 9. SUPPORTING DOCUMENTATION AND OPERATIONAL CONSIDERATIONS

#### 9.1. Appendix 1: Regulatory, Ethical, and Study Oversight Considerations

##### 9.1.1. Regulatory and Ethical Considerations

- This study will be conducted in accordance with the protocol and with:
  - Consensus ethical principles derived from international guidelines including the Declaration of Helsinki and Council for International Organizations of Medical Sciences (CIOMS) International Ethical Guidelines
  - Applicable International Conference on Harmonisation (ICH) Good Clinical Practice (GCP) Guidelines
  - Applicable laws and regulations
- The protocol, protocol amendments, informed consent form (ICF), IB and other relevant documents (*e.g.*, advertisements) must be submitted to an IRB/IEC by the investigator and reviewed and approved by the IRB/IEC before the study is initiated.
- Any amendments to the protocol will require IEC/IRB approval before implementation of changes made to the study design, except for changes necessary to eliminate an immediate hazard to study participants.
- Protocols and any substantial amendments to the protocol will require health authority approval prior to initiation except for changes necessary to eliminate an immediate hazard to study participants.
- The investigator will be responsible for the following:
  - Providing written summaries of the status of the study to the IRB/IEC annually or more frequently in accordance with the requirements, policies, and procedures established by the IRB/EC
  - Notifying the IRB/IEC of SAE or other significant safety findings as required by IRB/IEC procedures
  - Providing oversight of the conduct of the study at the site and adherence to requirements of 21 CFR, ICH guidelines, the IRB/IEC, European regulation 536/2014 for clinical studies (if applicable), European Medical Device Regulation 2017/745 for clinical device research (if applicable), and all other applicable local regulations

##### 9.1.2. Financial Disclosure

Investigators and sub-investigators will provide the sponsor with sufficient, accurate financial information as requested to allow the sponsor to submit complete and accurate financial certification or disclosure statements to the appropriate regulatory authorities.

Investigators are responsible for providing information on financial interests during the course of the study and for 1 year after completion of the study.

##### 9.1.3. Informed Consent Process

Given that this trial is being conducted in participants with COVID-19, and given that some of the participants will be mechanically ventilated upon enrolment, alternative procedures to obtain informed consent will be used. Participants may be consented in three ways, as described below. The EMA *Guidance on the Management of Clinical Trials during the COVID-19 (Coronavirus) pandemic* (Version 2, 27-Mar-20) [EMA, 2020] and the FDA *Guidance on Conduct of Clinical Trials of Medical Products during the COVID-19 Pandemic* (updated 16-Apr-20) [FDA, 2020] have been considered.

**1. Participant is enrolled by signing the informed consent.**
If the participant is capable of understanding and signing the informed consent (paper or eConsent), the below points will apply:

- The investigator or his/her representative will explain the nature of the study to the participant and answer all questions regarding the study.
- The medical record must include a statement that written (or electronic) informed consent was obtained before the participant was enrolled in the study and the date the written consent was obtained. The authorized person obtaining informed consent must also sign the informed consent form (ICF). If paper consent is used, and there is an infection risk to the person obtaining consent, the trial participant and the person obtaining consent may sign and date separate ICFs. All relevant records should be archived in the Investigator Site File. A correctly signed and dated ICF should be obtained from the trial participant as soon as possible.
- Consent may be collected before the 48h screening window if the investigator considers that the participant is likely to become eligible, and wants to ensure that consent is given by the participant rather than a legally authorized representative (LAR).

**2. Participant is enrolled by consenting orally.**
If the participant is capable of understanding, but not able to provide written consent (paper or eConsent), informed consent can be obtained orally, the below points will apply:

- The investigator or his/her representative will explain the nature of the study to the participant and answer all questions regarding the study.
- Oral consent must be given in the presence of an impartial witness. In such cases, the witness is required to sign and date the ICF (paper or electronic) and the investigator is expected to record how the impartial witness was selected. All relevant records should be archived in the Investigator Site File. A correctly signed and dated ICF should be obtained from the trial participant as soon as possible.

**3. Participant is enrolled by his/her legally authorized representative (LAR) (written or oral).**

If the participant is unable to provide informed consent, the investigator or delegate can obtain consent from an LAR. If consent is obtained from an LAR, the following points will apply:

- The investigator or his/her representative will explain the nature of the study to the LAR and answer all questions regarding the study.
- Best efforts will be undertaken to obtain written consent (paper, email or eConsent) from an LAR. However, if the LAR is not able to provide written or electronic consent for logistical reasons, informed consent can be obtained orally from the LAR. As above for oral participant consent, oral LAR consent must be given to the investigator in the presence of an impartial witness. In such cases, the witness is required to sign and date the ICF and the investigator is expected to record how the impartial witness was selected. A correctly signed and dated ICF should be obtained from the LAR as soon as possible.
- The investigator has responsibility for applying local laws in the matter of who has the capacity to consent and who qualifies as an LAR of a potential participant.
- An LAR is defined as an individual or juridical or other body authorized under applicable law to consent, on behalf of a prospective participant, to the participant's participation in the clinical trial.
- An LAR can be a member of the participant's family, next-of-kin or trusted person previously designated by the participant.
- As soon as is practically possible following a participant regaining capacity, participants will be asked to provide informed consent to remain in the study. If they decline, then they will be withdrawn from the study. The participant will decide if they want to allow samples already collected to be used or request for them to be discarded if not yet analyzed.
- All relevant records should be archived in the Investigator Site File.

**Documentation of Informed consent process**

The management of the informed consent process will be documented in the medical source notes. This will include which process was used to obtain consent. The following should be noted in regard to the informed consent process:

- If consent is provided orally by the participant or LAR in the presence of an impartial witness, the witness name, time and date of witnessing must be recorded in medical source notes. The investigator must also document how the impartial witness was selected.
- If consent is provided by LAR, name, time and date of LAR and name of the personnel at site that collected this information.
- Participants/LAR must be informed that their participation is voluntary. Participants and/or LAR will be required to sign electronically or on paper (or agree orally) to a statement of informed consent that meets the requirements of 21

CFR 50, local regulations, ICH guidelines, Health Insurance Portability and Accountability Act (HIPAA) requirements, where applicable, and the IEC or study center.

A copy of all signed ICF(s) must be provided to the participant or the participant's LAR.

GSK (alone or working with others) may use participant's coded study data and samples and other information to carry out this study; understand the results of this study; learn more about otilimab or about the study disease; publish the results of these research efforts; work with government agencies or insurers to have otilimab approved for medical use or approved for payment coverage.

The ICF contains a separate section that addresses the use of participant data and remaining samples for optional further research. The investigator or authorized designee will inform each participant or their LAR of the possibility of further research not related to the study/disease. Participants or their LAR will be told that they are free to refuse to participate and may withdraw their consent at any time and for any reason during the storage period. A separate signature (participant or LAR) will be required to document agreement to allow any participant data and/or remaining leftover samples to be used for further research not related to the study/disease. Participants or their LAR who decline further research will tick the corresponding "No" box.

##### 9.1.4. Data Protection

- Participants will be assigned a unique identifier by the sponsor. Any participant records or datasets that are transferred to the sponsor will contain the identifier only; participant names or any information which would make the participant identifiable will not be transferred.
- The participant must be informed that his/her personal study-related data will be used by the sponsor in accordance with local data protection law. The level of disclosure must also be explained to the participant who will be required to give consent for their data to be used as described in the informed consent.
- The participant must be informed that his/her medical records may be examined by Clinical Quality Assurance auditors or other authorized personnel appointed by the sponsor, by appropriate IRB/IEC members, and by inspectors from regulatory authorities.

##### 9.1.5. Committees Structure

An Independent Data Monitoring Committee (IDMC) comprised of clinical experts external to GSK will review unblinded data at defined timepoints during the study. If deemed appropriate by the IDMC, or upon request by GSK or investigators, additional timepoints for review may be added.

Details of the structure and function of the IDMC, and analysis plan for IDMC reviews, are outlined in the IDMC Charter, which is available upon request.

In addition to the IDMC, the GSK SRT will review blinded safety data at a minimum of a weekly basis to ensure participant safety , which includes safety signal detection at any time during the study. Details of this are included in Section 8.5.

In the unlikely event that the IDMC cannot be convened for a scheduled meeting, the GSK Independent Safety Review Committee (iSRC) will review the unblinded outputs (as outlined in the IDMC Charter).

##### 9.1.6. Dissemination of Clinical Study Data

- Where required by applicable regulatory requirements, an investigator signatory will be identified for the approval of the clinical study report. The investigator will be provided reasonable access to statistical tables, figures, and relevant reports and will have the opportunity to review the complete study results at a GSK site or other mutually-agreeable location.
- GSK will also provide all investigators who participated in the study with a summary of the study results and will tell the investigators what treatment their participants received. The investigator(s) is/are encouraged to share the summary results with the study participants, as appropriate.
- Under the framework of the SHARE initiative, GSK intends to make anonymized participant-level data from this trial available to external researchers for scientific analyses or to conduct further research that can help advance medical science or improve patient care. This helps ensure the data provided by trial participants are used to maximum effect in the creation of knowledge and understanding. Requests for access may be made through www.clinicalstudydatarequest.com.
- GSK will provide the investigator with the randomization codes for their site only after completion of the full statistical analysis.
- The procedures and timing for public disclosure of the protocol and results summary and for development of a manuscript for publication for this study will be in accordance with GSK Policy.
- GSK intends to make anonymized patient-level data from this trial available to external researchers for scientific analyses or to conduct further research that can help advance medical science or improve patient care. This helps ensure the data provided by trial participants are used to maximum effect in the creation of knowledge and understanding
- A manuscript will be progressed for publication in the scientific literature if the results provide important scientific or medical knowledge.

##### 9.1.7. Data Quality Assurance

- All participant data relating to the study will be recorded on printed or electronic CRF unless transmitted to the sponsor or designee electronically (*e.g.*, laboratory data). The investigator is responsible for verifying that data entries are accurate and correct by physically or electronically signing the CRF.
- Guidance on completion of CRFs will be provided in the eCRF Guidelines.

- Quality tolerance limits (QTLs) will be pre-defined in the QTL Plan to identify systematic issues that can impact participant safety and/or reliability of study results. These pre-defined parameters will be monitored during and at the end of the study and all deviations from the QTLs and remedial actions taken will be summarized in the clinical study report.
- The investigator must permit study-related monitoring, audits, IRB/IEC review, and regulatory agency inspections and provide direct access to source data documents.
- Monitoring details describing strategy including definition of study critical data items and processes (e.g., risk-based initiatives in operations and quality such as Risk Management and Mitigation Strategies and Analytical Risk-Based Monitoring), methods, responsibilities and requirements, including handling of noncompliance issues and monitoring techniques (central, remote, or on-site monitoring) are provided in the Monitoring Plan.
- The sponsor or designee is responsible for the data management of this study including quality checking of the data.
- The sponsor assumes accountability for actions delegated to other individuals (e.g., Contract Research Organizations).
- Records and documents, including signed ICF, pertaining to the conduct of this study must be retained by the investigator for 25 years from the issue of the final Clinical Study Report (CSR)/ equivalent summary unless local regulations or institutional policies require a longer retention period. No records may be destroyed during the retention period without the written approval of the sponsor. No records may be transferred to another location or party without written notification to the sponsor.

##### 9.1.8. Source Documents

- Source documents provide evidence for the existence of the participant and substantiate the integrity of the data collected. Source documents are filed at the investigator's site.
- Data reported on the CRF or entered in the eCRF that are transcribed from source documents must be consistent with the source documents or the discrepancies must be explained. The investigator may need to request previous medical records or transfer records, depending on the study. Also, current medical records must be available.
- Definition of what constitutes source data and its origin can be found in the Source Data Acknowledgment.
- The investigator must maintain accurate documentation (source data) that supports the information entered in the CRF.
- Study monitors will perform ongoing monitoring (remote or on-site) as per the Monitoring Plan to confirm that data entered into the CRF by authorized site personnel are accurate, complete, and verifiable from source documents; that the safety and rights of participants are being protected; and that the study is being

conducted in accordance with the currently approved protocol and any other study agreements, ICH GCP, and all applicable regulatory requirements.

##### 9.1.9. Study and Site Start and Closure

GSK or designee reserves the right to close the study site or terminate the study at any time for any reason at the sole discretion of GSK. Study sites will be closed upon study completion. A study site is considered closed when all required documents and study supplies have been collected and a study-site closure visit has been performed.

The investigator may initiate study-site closure at any time, provided there is reasonable cause and sufficient notice is given in advance of the intended termination.

Reasons for the early closure of a study site by the sponsor or investigator may include but are not limited to:

For study termination:

- If the study is prematurely terminated or suspended, the sponsor shall promptly inform the investigators, the IECs/IRBs, the regulatory authorities, and any contract research organization(s) used in the study of the reason for termination or suspension, as specified by the applicable regulatory requirements. The investigator shall promptly inform the participant and should assure appropriate participant therapy and/or follow-up
- Discontinuation of further study intervention development

For site termination:

- Failure of the investigator to comply with the protocol, the requirements of the IRB/IEC or local health authorities, the sponsor's procedures, or GCP guidelines
- Inadequate or no recruitment of participants (evaluated after a reasonable amount of time) by the investigator

##### 9.1.10. Publication Policy

- The results of this study may be published or presented at scientific meetings. If this is foreseen, the investigator agrees to submit all manuscripts or abstracts to the sponsor before submission. This allows the sponsor to protect proprietary information and to provide comments.
- The sponsor will comply with the requirements for publication of study results. In accordance with standard editorial and ethical practice, the sponsor will generally support publication of multicenter studies only in their entirety and not as individual site data. In this case, a coordinating investigator will be designated by mutual agreement.
- Authorship will be determined by mutual agreement and in line with International Committee of Medical Journal Editors authorship requirements.

#### 9.2. Appendix 2: Clinical Laboratory Tests

The tests detailed in Table 3 will be performed by the local laboratory.

Protocol-specific requirements for inclusion or exclusion of participants are detailed in Section 5 of the protocol.

Additional tests may be performed at any time during the study as determined necessary by the investigator or required by local regulations.

**Table 3 Protocol-Required Safety Laboratory Tests**

| Laboratory Assessments[1] | Parameters | | |
|---|---|---|---|
| **Hematology** | Hemoglobin | RBC Indices: | WBC count (differential) absolute and/or %: |
| | Hematocrit | MCV | Neutrophils |
| | Platelet Count | MCH | Lymphocytes |
| | RBC Count | %Reticulocytes | Monocytes |
| | | | Eosinophils<br>Basophils |
| **Coagulation** | APTT / APTR | PT | INR |
| **Clinical Chemistry** | Urea (BUN) | Phosphate | AST / SGOT |
| | Creatinine | Troponin I[4] | ALT / SGPT |
| | Glucose[2] | D-dimer | Gamma-GT |
| | Potassium | Ferritin[5] | Total bilirubin |
| | Sodium | CRP | Direct bilirubin |
| | Calcium | LDH | ALP[7] |
| | Lactate[3] | Procalcitonin[5, 6] | Total protein |
| | Creatine kinase | ESR[5] | Albumin |
| **Pregnancy** | Highly sensitive serum or urine human chorionic gonadotropin (hCG) pregnancy test (as needed for women of childbearing potential) | | |
| **CRS Event** | If a CRS is suspected, the following samples should be taken as soon as possible after suspected CRS on-set **and** again 24 h later: Hematology, CRP, D-dimer, ferritin, PD (cytokine sample – see Section 1.3) | | |

1. Hematology, Coagulation and Clinical Chemistry testing at the following time-points (unless otherwise indicated below): Screening, Day 1, Day 2, Day 4, Day 7, Day 14, Day 28 and Discharge
2. Record glucose fasting / non-fasting status
3. Can be measured from arterial blood gas test
4. If troponin I is not available locally, troponin T can be reported
5. To be measured at Screening, Day 7 and Day 14 only
6. Procalcitonin to be measured if the test is available locally
7. If alkaline phosphatase is elevated, consider fractionating

#### 9.3. Appendix 3: AEs and SAEs: Definitions and Procedures for Recording, Evaluating, Follow-up, and Reporting

##### 9.3.1. Definition of AE

| **AE Definition** |
|---|
| • An AE is any untoward medical occurrence in a clinical study participant, temporally associated with the use of a study intervention, whether or not considered related to the study intervention.<br>• NOTE: An AE can therefore be any unfavorable and unintended sign (including an abnormal laboratory finding), symptom, or disease (new or exacerbated) temporally associated with the use of a study intervention. |

| **Events Meeting the AE Definition** |
|---|
| • Any abnormal laboratory test results (hematology, clinical chemistry, or urinalysis) or other safety assessments (*e.g.*, ECG, radiological scans, vital signs measurements), including those that worsen from baseline, considered clinically significant in the medical and scientific judgment of the investigator (*i.e.*, not related to progression of underlying disease).<br>• Exacerbation of a chronic or intermittent pre-existing condition including either an increase in frequency and/or intensity of the condition.<br>• New conditions detected or diagnosed after study intervention administration even though it may have been present before the start of the study.<br>• Signs, symptoms, or the clinical sequelae of a suspected intervention- intervention interaction.<br>• Signs, symptoms, or the clinical sequelae of a suspected overdose of either study intervention or a concomitant medication. Overdose per se will not be reported as an AE/SAE unless it is an intentional overdose taken with possible suicidal/self-harming intent. Such overdoses should be reported regardless of sequelae.<br>• "Lack of efficacy" or "failure of expected pharmacological action" per se will not be reported as an AE or SAE. Such instances will be captured in the efficacy assessments. However, the signs, symptoms, and/or clinical sequelae resulting from lack of efficacy will be reported as AE or SAE if they fulfil the definition of an AE or SAE. |

| **Events NOT Meeting the AE Definition** |
|---|
| • Any clinically significant abnormal laboratory findings or other abnormal safety assessments which are associated with the underlying disease, unless judged by the investigator to be more severe than expected for the participant's condition. |

- The disease/disorder being studied or expected progression, signs, or symptoms of the disease/disorder being studied, unless more severe than expected for the participant's condition.
- Medical or surgical procedure (*e.g.*, endoscopy, appendectomy): the condition that leads to the procedure is the AE.
- Situations in which an untoward medical occurrence did not occur (social and/or convenience admission to a hospital).
- Anticipated day-to-day fluctuations of pre-existing disease(s) or condition(s) present or detected at the start of the study that do not worsen.

##### 9.3.2. Definition of SAE

| **An SAE is defined as any serious adverse event that, at any dose:** |
|---|
| **a. Results in death** |
| **b. Is life-threatening**<br>The term 'life-threatening' in the definition of 'serious' refers to an event in which the participant was at risk of death at the time of the event. It does not refer to an event, which hypothetically might have caused death, if it were more severe. |
| **c. Requires inpatient hospitalization or prolongation of existing hospitalization**<br>• In general, hospitalization signifies that the participant has been admitted (usually involving at least an overnight stay) at the hospital or emergency ward for observation and/or treatment that would not have been appropriate in the physician's office or outpatient setting. Complications that occur during hospitalization are AE. If a complication prolongs hospitalization or fulfils any other serious criteria, the event is serious. When in doubt as to whether "hospitalization" occurred or was necessary, the AE should be considered serious.<br>• Hospitalization for elective treatment of a pre-existing condition that did not worsen from baseline is not considered an AE. |
| **d. Results in persistent or significant disability/incapacity**<br>• The term disability means a substantial disruption of a person's ability to conduct normal life functions.<br>• This definition is not intended to include experiences of relatively minor medical significance such as uncomplicated headache, nausea, vomiting, diarrhea, influenza, and accidental trauma (*e.g.* sprained ankle) which may interfere with or prevent everyday life functions but do not constitute a substantial disruption. |
| **e. Is a congenital anomaly/birth defect** |
| **f. Other situations:**<br>• Medical or scientific judgment should be exercised by the investigator in deciding whether SAE reporting is appropriate in other situations such as significant medical events that may jeopardize the participant or may require medical or surgical intervention to prevent one of |

the other outcomes listed in the above definition. These events should usually be considered serious.

- Examples of such events include invasive or malignant cancers, intensive treatment for allergic bronchospasm, blood dyscrasias, convulsions, acute deterioration of liver function, or development of intervention dependency or intervention abuse.

##### 9.3.3. Definition of Cardiovascular Events

| Cardiovascular Events (CV) Definition: |
|---|
| Investigators will be required to fill out the specific CV event page of the eCRF for the following AEs and SAEs:<br>• Myocardial infarction/unstable angina<br>• Congestive heart failure<br>• Arrhythmias<br>• Valvulopathy<br>• Pulmonary hypertension<br>• Cerebrovascular events/stroke and transient ischemic attack<br>• Peripheral arterial thromboembolism<br>• Deep venous thrombosis/pulmonary embolism<br>• Revascularization |

##### 9.3.4. Recording and Follow-Up of AE and SAE

| AE and SAE Recording |
|---|
| • When an AE/SAE occurs, it is the responsibility of the investigator to review all documentation (*e.g*. hospital progress notes, laboratory, and diagnostics reports) related to the event.<br>• The investigator will then record all relevant AE/SAE information.<br>• It is **not** acceptable for the investigator to send photocopies of the participant's medical records to GSK in lieu of completion of the GSK required form.<br>• There may be instances when copies of medical records for certain cases are requested by GSK. In this case, all participant identifiers, with the exception of the participant number, will be redacted on the copies of the medical records before submission to GSK.<br>• The investigator will attempt to establish a diagnosis of the event based on signs, symptoms, and/or other clinical information. Whenever possible, the diagnosis (not the individual signs/symptoms) will be documented as the AE/SAE. |

| **Assessment of Intensity** |
|---|
| The investigator will make an assessment of intensity for each AE and SAE reported during the study and assign it to 1 of the following categories:<br>• Mild: An event that is easily tolerated by the participant, causing minimal discomfort and not interfering with everyday activities.<br>• Moderate: An event that causes sufficient discomfort and interferes with normal everyday activities.<br>• Severe: An event that prevents normal everyday activities. An AE that is assessed as severe should not be confused with an SAE. Severe is a category utilized for rating the intensity of an event; and both AE and SAE can be assessed as severe.<br>• An event is defined as 'serious' when it meets at least 1 of the predefined outcomes as described in the definition of an SAE, NOT when it is rated as severe. |
| **Assessment of Causality** |
| • The investigator is obligated to assess the relationship between study intervention and each occurrence of each AE/SAE.<br>• A "reasonable possibility" of a relationship conveys that there are facts, evidence, and/or arguments to suggest a causal relationship, rather than a relationship cannot be ruled out.<br>• The investigator will use clinical judgment to determine the relationship.<br>• Alternative causes, such as underlying disease(s), concomitant therapy, and other risk factors, as well as the temporal relationship of the event to study intervention administration will be considered and investigated.<br>• The investigator will also consult the Investigator's Brochure (IB) and/or Product Information, for marketed products, in his/her assessment.<br>• For each AE/SAE, the investigator **must** document in the medical notes that he/she has reviewed the AE/SAE and has provided an assessment of causality.<br>• There may be situations in which an SAE has occurred, and the investigator has minimal information to include in the initial report to GSK. However, **it is very important that the investigator always make an assessment of causality for every event before the initial transmission of the SAE data to** GSK**.**<br>• The investigator may change his/her opinion of causality in light of follow-up information and send an SAE follow-up report with the updated causality assessment.<br>• The causality assessment is one of the criteria used when determining regulatory reporting requirements. |

| **Follow-up of AE and SAE** |
|---|
| • The investigator is obligated to perform or arrange for the conduct of supplemental measurements and/or evaluations as medically indicated or as requested by GSK to elucidate the nature and/or causality of the AE or SAE as fully as possible. This may include additional |

| |
|---|
| laboratory tests or investigations, histopathological examinations, or consultation with other health care professionals.<br>• If a participant dies during participation in the study or during a recognized follow-up period, the investigator will provide GSK with a copy of any post-mortem findings including histopathology.<br>• New or updated information will be recorded in the originally submitted documents.<br>• **The investigator will submit any updated SAE data to GSK within 24 hours of receipt of the information.** |

##### 9.3.5. Reporting of SAE to GSK

| SAE Reporting to GSK via Electronic Data Collection Tool |
|---|
| • The primary mechanism for reporting SAE to GSK will be the electronic data collection tool.<br>• **If the electronic system is unavailable, then the site will use the paper SAE data collection tool (see next section) to report the event within 24 hours.**<br>• The site will enter the SAE data into the electronic system as soon as it becomes available.<br>• The investigator or medically-qualified sub-investigator must show evidence within the eCRF (*e.g.*, check review box, signature, *etc.*) of review and verification of the relationship of each SAE to investigational product/study participation (causality) within 72 hours of SAE entry into the eCRF.<br>• After the study is completed at a given site, the electronic data collection tool will be taken off-line to prevent the entry of new data or changes to existing data.<br>• If a site receives a report of a new SAE from a study participant or receives updated data on a previously reported SAE after the electronic data collection tool has been taken off-line, then the site can report this information on a paper SAE form (see next section) or to the SAE coordinator by telephone.<br>• Contacts for SAE reporting can be found in Study Reference Manual. |

| SAE Reporting to GSK via Paper Data Collection Tool |
|---|
| • Facsimile transmission of the SAE paper data collection tool is the preferred method to transmit this information to the SAE coordinator.<br>• In rare circumstances and in the absence of facsimile equipment, notification by telephone is acceptable with a copy of the SAE data collection tool sent by overnight mail or courier service.<br>• Initial notification via telephone does not replace the need for the investigator to complete and sign the SAE data collection tool within the designated reporting time frames.<br>• Contacts for SAE reporting can be found in Study Reference Manual. |

#### 9.4. Appendix 4: Contraceptive and Barrier Guidance

##### 9.4.1. Definitions:

**Woman of Childbearing Potential (WOCBP)**

Women in the following categories are considered WOCBP (fertile):

1. Following menarche
2. From the time of menarche until becoming post-menopausal unless permanently sterile (see below)

**Notes**:

- A postmenopausal state is defined as no menses for 12 months without an alternative medical cause.
  - Females on HRT and whose menopausal status is in doubt will be required to use one of the non-estrogen hormonal highly effective contraception methods if they wish to continue their HRT during the study.
- Permanent sterilization methods (for the purpose of this study) include:
  - Documented hysterectomy
  - Documented bilateral salpingectomy
  - Documented bilateral oophorectomy
- For individuals with permanent infertility due to an alternate medical cause other than the above, (*e.g*., Mullerian agenesis, androgen insensitivity, gonadal dysgenesis), investigator discretion should be applied to determining study entry.

- **Note:** Documentation can come from the site personnel's review of the participant's medical records, medical examination, or medical history interview.
-

If fertility is unclear (*e.g*., amenorrhea in adolescents or athletes) and a menstrual cycle cannot be confirmed before first dose of study intervention, additional evaluation should be considered.

##### 9.4.2. Contraception Guidance:

| |
|---|
| • **CONTRACEPTIVES[a] ALLOWED DURING THE STUDY INCLUDE:** |
| • **Highly Effective Methods[b] That Have Low User Dependency** *Failure rate of <1% per year when used consistently and correctly.* |
| • Implantable progestogen-only hormone contraception associated with inhibition of ovulation[c] |
| • Intrauterine device (IUD) |
| • Intrauterine hormone-releasing system (IUS)[c] |

| |
|---|
| • Bilateral tubal occlusion |
| • Azoospermic partner (vasectomized or due to a medical cause)<br>• Azoospermia is a highly effective contraceptive method provided that the partner is the sole sexual partner of the woman of childbearing potential and the absence of sperm has been confirmed. If not, an additional highly effective method of contraception should be used. Spermatogenesis cycle is approximately 90 days.<br>•<br>Note: documentation of azoospermia for a male participant can come from the site personnel's review of the participant's medical records, medical examination, or medical history interview |
| • **Highly Effective Methods[b] That Are User Dependent** *Failure rate of <1% per year when used consistently and correctly.* |
| • Combined (estrogen- and progestogen-containing) hormonal contraception associated with inhibition of ovulation[c]<br>o oral<br>o intravaginal<br>o transdermal<br>o injectable |
| • Progestogen-only hormone contraception associated with inhibition of ovulation[c]<br>o oral<br>o injectable |
| • Sexual abstinence<br>• Sexual abstinence is considered a highly effective method only if defined as refraining from heterosexual intercourse during the entire period of risk associated with the study intervention and is the participant's normal practice. The reliability of sexual abstinence needs to be evaluated in relation to the duration of the study and the preferred and usual lifestyle of the participant. |
| a. Contraceptive use by men or women should be consistent with local regulations regarding the use of contraceptive methods for those participating in clinical studies.<br>b. Failure rate of <1% per year when used consistently and correctly. Typical use failure rates differ from those when used consistently and correctly.<br>c. Male condoms must be used in addition to hormonal contraception. If locally required, in accordance with Clinical Trial Facilitation Group (CTFG) guidelines, acceptable contraceptive methods are limited to those which inhibit ovulation as the primary mode of action.<br><br>Note: Periodic abstinence (calendar, sympto-thermal, post-ovulation methods), withdrawal (coitus interruptus), spermicides only, and lactational amenorrhea method (LAM) are not acceptable methods of contraception. Male condom and female condom should not be used together (due to risk of failure from friction). |

### 9.5. Appendix 5: Abbreviations and Trademarks

| | |
|---|---|
| Ab | Antibody |
| ADE | Adverse Device Effect |
| AE(s) | Adverse event(s) |
| AESI | Adverse event of special interest |
| ALI | Acute Lung Injury |
| ALP | Alkaline phosphatase |
| ALT | Alanine transaminase |
| ANC | Absolute Neutrophil Count |
| APTT/APTR | Activated Partial Thromboplastin Time/Activated Partial Thromboplastin Time Ratio |
| ARDS | Acute Respiratory Distress Syndrome |
| AST | Aspartate transaminase |
| AUC | Area Under the Curve |
| BAL | Bronchoalveolar Lavage |
| BiPAP | Bilevel Positive Airway Pressure |
| BUN | Blood Urea Nitrogen |
| CAR | Chimeric Antigen Receptor |
| CFR | Code of Federal Regulations |
| CIOMS | Council for International Organizations of Medical Sciences |
| CL/F | Clearance |
| $C_{max}$ | Maximum Concentration |
| CONSORT | Consolidated Standards of Reporting Trials |
| COVID-19 | Coronavirus Disease 2019 |
| CPAP | Continuous Positive Airway Pressure |
| CRF | Case Report Form |
| CRP | C-reactive protein |
| CRS | Cytokine Release Syndrome |
| CSR | Clinical Study Report |
| CT | Computed Tomography |
| CTCAE | Common terminology criteria for Adverse Events |
| CTFG | Clinical Trial Facilitation Group |
| CV | Cardiovascular |
| ECG | Electrocardiogram |
| ECMO | Extracorporeal Membrane Oxygen |
| eCRF | Electronic case report form |
| EMA | European Medicines Agency |
| ESR | Erythrocyte sedimentation rate |
| FDA | Food and Drug Administration |
| $FiO_2$ | Concentration of Inspired Oxygen |
| FU | Follow-up |
| GCP | Good Clinical Practice |
| GCS | Glasgow Coma Scale |
| GFR | Glomerular filtration rate |
| GM-CSF | Granulocyte-macrophage colony stimulating factor |

| | |
|---|---|
| GM-CSFR | Granulocyte-macrophage colony stimulating factor receptor |
| GSK | GlaxoSmithKline |
| h | Hour |
| HBsAg | Hepatitis B-surface Antigen |
| hCG | Human chorionic gonadotropin |
| HF | Human Factors |
| HIPAA | Health Insurance Portability and Accountability Act |
| HIV | Human Immunodeficiency Virus |
| HRT | Hormone replacement therapy |
| HV | Healthy volunteer |
| IB | Investigator’s brochure |
| ICF | Informed consent form |
| ICH | International Conference on Harmonisation |
| ICU | Intensive Care Unit |
| IDMC | Independent Data Monitoring Committee |
| IEC | Institutional ethics committee |
| IFNγ | Interferon Gamma |
| IFU | Instructions For Use |
| IL | Interleukin |
| IMP | Investigational medicinal product |
| INR | International normalized ratio |
| IRB | Institutional review board |
| IRT | Interactive Response Technology |
| iSRC | Independent Safety Review Committee |
| ITT | Intent to Treat |
| IUD | Intrauterine device |
| IUS | Intrauterine hormone-releasing system |
| IV | Intravenous |
| JAK | Janus Kinase |
| L | Liter |
| LAM | Lactational amenorrhea method |
| LAR | Legally Authorized Representative |
| LDH | Lactate dehydrogenase |
| mAb | Monoclonal antibody |
| MAP | Mean Arterial Pressure |
| MCP-1α | Monocyte Chemoattractant Protein-1 Alpha |
| MCV | Mean cell volume |
| MCH | Mean corpuscular hemoglobin |
| MedDRA | Medical dictionary for regulatory activities |
| μg | Microgram |
| $m^2$ | Square Meters |
| mg | Milligram |
| Min | Minute |
| MIP-1 | Macrophage Inflammatory Protein-1 |
| mL | Milliliter |
| $mm^3$ | Cubic Millimeters |

| | |
|---|---|
| mmHg | Millimeters of Mercury |
| MS | Multiple sclerosis |
| MSDS | Material safety data sheet |
| NCI-CTCAE | National Cancer Institute Common Terminology Criteria for Adverse Events |
| NOAEL | No observed adverse effect level |
| NYHA | New York Heart Association |
| OR | Odds Ratio |
| PAP | Pulmonary alveolar proteinosis |
| PD | Pharmacodynamic |
| PFS | Prefilled syringe |
| PI | Principal Investigator |
| PT | Prothrombin Time |
| PK | Pharmacokinetics |
| RA | Rheumatoid arthritis |
| RBC | Red Blood Cell |
| RRT | Renal replacement therapy |
| RT-PCR | Reverse Transcriptase Polymerase Chain Reaction |
| SADE | Serious Adverse Device Effect |
| SARS-CoV-2 | Severe Acute Respiratory Syndrome–Coronavirus-2 |
| SC | Subcutaneous |
| SAE(s) | Serious adverse event(s) |
| SoA | Schedule of Activities |
| SOFA | Sequential Organ Failure Assessment |
| $SpO_2$ | Blood Oxygen Saturation |
| SRM | Study Reference Manual |
| SRT | Safety Review Team |
| SSD | Safety Syringe Device |
| TB | Tuberculosis |
| TNFα | Tumor necrosis factor alpha |
| ULN | Upper limit of normal |
| USADE | Unanticipated Serious Adverse Device Effect |
| VAP | Ventilator-associated Pneumonia |
| WBC | White Blood Cell |
| WHO | World Health Organization |
| WOCBP | Woman of Childbearing Potential |
| WONCBP | Woman of Non-Childbearing Potential |

#### Trademark Information

| Trademarks of the GlaxoSmithKline group of companies |
|---|
| None |

| Trademarks not owned by the GlaxoSmithKline group of companies |
|---|
| MedDRA |

#### 9.6. Appendix 6: Protocol Amendment History

The Protocol Amendment Summary of Changes Table for the current amendment is located directly before the Table of Contents (TOC).

**Amendment 1: 18-MAY-2020**

This amendment is considered to be substantial based on the criteria set forth in Article 10(a) of Directive 2001/20/EC of the European Parliament and the Council of the European Union.

**Overall Rationale for the Amendment:** modifications to the protocol in response to regulatory feedback, and clarifications based on investigator feedback. The key change is revision of the primary endpoint (where definition of "free of respiratory failure" is participants in categories 1-4 on GSK Ordinal Scale). Other changes are (1) to revise recording of blood pressure and pulse rate; (2) to add details of randomization caps; (3) to simplify/clarify some Inclusion and Exclusion Criteria; (4) to clarify medications permitted during the study.

| Section # and Name | Description of Change | Brief Rationale |
|---|---|---|
| **Section 1.1 and Section 3 Objectives and Endpoints** | Revised "independent of supplementary oxygen" to "free of respiratory failure" for Primary Endpoint; added or revised Secondary Endpoints (added Time to all-cause mortality, Time to recovery from respiratory failure and Time to last dependence on supplementary oxygen); added or revised some Exploratory Endpoints (added Change in COVID-19 signs and symptoms).<br><br>Removed details of organ support in the Ordinal Scale. | Regulatory feedback, new FDA guidance for the development of COVID-19 treatments, and clarification.<br><br>Clarification based on regulatory and investigator feedback and to ensure alignment with Exclusion Criteria 3 and description of categories in the Ordinal Scale. |
| **Section 1.3 Schedule of Activities** | Revised recording of blood pressure and pulse rate to daily up to Day 28 or discharge. | Regulatory request. |
| **Section 4.1.2. Main Cohort (after 20 Participants)** | Acknowledgment that randomization caps will be used. | Text added in line with the IDMC Charter for transparency purposes. |
| **Section 5.1. Inclusion Criteria** | Inclusion 2: Removal of SpO2 ≤93% on room air. | #2 Feedback from investigators based on clinical practice to identify participants with oxygenation impairment. |

| | | |
|---|---|---|
| | Inclusion 4: Clarification of sexual abstinence. | #4 Regulatory feedback. |
| **Section 5.2. Exclusion Criteria** | Exclusion 3: Added use of high-dose or multiple vasopressors as an exclusion. | Clarification of Exclusion Criteria related to organ support. |
| **Section 6.6.2. Medication/Treatment Permitted During the Study** | Added statement that all treatments for participants in the ICU are permitted, and clarification of additional treatments of COVID-19 related disease. | Clarification based on regulatory and investigator feedback and to ensure alignment with Exclusion Criteria 3 and description of categories on the ordinal scale. |
| **Section 7.2.6, Ordinal Scale** | Removed details of organ support in the Ordinal Scale. | Clarification based on regulatory and investigator feedback and to ensure alignment with Exclusion Criteria 3 and description of categories in the Ordinal Scale. |
| **Section 8.1. Statistical Hypotheses** | Revised null hypothesis for consistency with changes to the Primary Endpoint and added alternative to the null hypothesis. | Regulatory feedback for Primary Endpoint and clarification for alternative to null hypothesis. |
| **Section 8.4.2. Primary Endpoint(s)** | Revised “independent of supplementary oxygen” to “free of respiratory failure” for Primary Endpoint. | Regulatory feedback. |
| **8.5. Interim Analysis** | Safety stopping rules have been put in place for the IDMC to stop the study if there is an excess risk of mortality. | Regulatory request. |
| **Section 9.4.2. Contraception Guidance:** | Clarification of situations when sexual abstinence is acceptable. | Regulatory request. |
| **All sections** | Other minor, grammatical and typographical corrections to improve readability. | |

### 10. REFERENCES

Alessandris S, Ferguson JG, Dodd AJ *et al.* Neutrophil GM-CSF receptor dynamics in acute lung injury. J Leukoc Biol 2019;105:1183-94.

Arentz M, Yim E, Klaff L, *et al*. Characteristics and outcomes of 21 critically ill patients with COVID-19 in Washington State. JAMA 2020; doi: 10.1001/jama.2020.4326.

Bhatraju PK, Ghassemieh BJ, Nichols M, *et al*. Covid-19 in critically ill patients in the Seattle region - case series. N Engl J Med 2020; doi: 10.1056/NEJMoa2004500.

Bozinovski S, Jones J, Beavitt SJ *et al.* Innate immune responses to LPS in mouse lung are suppressed and reversed by neutralization of GM-CSF via repression of TLR-4. Am J Physiol Lung Cell Mol Physiol 2004;286:L877-85.

Bozinovski S, Jones JE, Vlahos R, *et al.* Granulocyte/macrophage-colony-stimulating factor (GM-CSF) regulates lung innate immunity to lipopolysaccharide through Akt/Erk activation of NFkappa B and AP-1 *in vivo*. J Biol Chem 2002;277:42808-14.

Burmester GR, McInnes IB, Kremer JM, *et al*. Mavrilimumab, a fully human granulocyte-macrophage colony-stimulating factor receptor α monoclonal antibody: long-term safety and efficacy in patients with rheumatoid arthritis. Arthritis Rheumatol 2018;70:679-689.

Cao B, Wang Y, Wen D, *et al*. A Trial of lopinavir-ritonavir in adults hospitalized with severe Covid-19. N Engl J Med 2020; doi: 10.1056/NEJMoa2001282.

Carsana L, Sonzogni A, Nasr A, *et al.* Pulmonary post-mortem findings in a large series of COVID-19 cases from Northern Italy 2020 medRxiv preprint doi: https://doi.org/10.1101/2020.04.19.20054262.

Coomes EA, Haghbayan H. Interleukin-6 in COVID-19: A Systematic Review and Meta-Analysis MedRix Preprint 2020 doi: https://doi.org/10.1101/2020.03.30.20048058.

DK, Betts AM. Antibody biodistribution coefficients. mAbs 2013;5:297-305.

EMA *Guidance on the Management of Clinical Trials during the COVID-19 (Coronavirus) pandemic* (Version 2, 27-Mar-20).

FDA Guidance on Conduct of Clinical Trials of Medical Products during the COVID-19 Pandemic (updated 16 April 2020). https://www.fda.gov/media/136238/download.

Grasselli G, Zangrillo A, Zanella A, *et al*. Baseline characteristics and outcomes of 1591 patients infected with SARS-CoV-2 admitted to ICUs of the Lombardy region, Italy. JAMA 2020; doi: 10.1001/jama.2020.5394.

Guo X, Higgs BW, Bay-Jensen AC, *et al*. Blockade of GM-CSF pathway induced sustained suppression of myeloid and T cell activities in rheumatoid arthritis. Rheumatology (Oxford) 2018;57:175-84.

Gupta A, Zecchin C, Fisheleva E, *et al.* Exposure-efficacy analysis in DMARD inadequate response rheumatoid arthritis patients treated with GSK3196165 along with methotrexate. Arthritis Rheumatol 2018;70(Supplement 10, abstract 2519).

Gupta A, Zecchin C, Watts S, *et al.* Model informed selection of Phase 3 dosing regimens not fully tested in Phase 2: anti-GM-CSF mAb, GSK3196165 in moderate to severe rheumatoid arthritis. ACoP10, Orlando FL, USA. Abstract M-024. ISSN:2688-3953, 2019.

Halstead ES, Chroneos ZC. Lethal influenza infection: Is a macrophage to blame? Expert Rev Anti-Infective Ther 2015;13:1425-8.

Halstead ES, Umstead TM, Davies ML, *et al.* GM-CSF overexpression after influenza a virus infection prevents mortality and moderates M1-like airway monocyte/macrophage polarization. Respir Res 2018;19:3.

Hamilton JA. GM-CSF-dependent inflammatory pathways. Front Immunol 2019;10:2055.

Hermine O, Mariette X, Tharaux PL, *et al*. 2020. Press release. https://www.aphp.fr/contenu/tocilizumab-improves-significantly-clinical-outcomes-patients-moderate-or-severe-covid-19.

Huang C, Wang Y, Li X, Ren L, *et al*. Clinical features of patients infected with 2019 novel coronavirus in Wuhan, China. Lancet 2020;395(10223):497-506.

Huang FF, Barnes PF, Feng Y, *et al.* GM-CSF in the lung protects against lethal influenza infection. Am J Respir Crit Care Med 2011;184:259-68.

ICNARC report on COVID-19 in critical care, 2020. https://www.icnarc.org/DataServices/Attachments/Download/c31dd38d-d77b-ea11-9124-00505601089b.

Kiniksa, 2020. Press release. https://investors.kiniksa.com/news-releases/news-release-details/kiniksa-announces-early-evidence-treatment-response-mavrilimumab.

Lan KKG, DeMets DL. Discrete sequential boundaries for clinical trials. Biometrika 1983;70:659-63.

Lee DW, Gardner R, Porter DL, *et al*. Current concepts in the diagnosis and management of cytokine release syndrome. Blood 2014;124:188-95.

LeVine AM, Reed JA, Kurak KE, *et al*. GM-CSF-deficient mice are susceptible to pulmonary group B streptococcal infection. J Clin Invest 1999;103:563-9.

Li H, Liu L, Zhang D, *et al.* SARS-CoV-2 and viral sepsis: observations and hypotheses. Lancet. 2020 Apr 17. pii: S0140-6736(20)30920-X. doi: 10.1016/S0140-6736(20)30920-X.

Liao M, Liu Y, Yuan J, *et al.* The landscape of lung bronchoalveolar immune cells in COVID-19 revealed by single-cell RNA sequencing. medRxiv preprint doi: https://doi.org/10.1101/2020.02.23.20026690, accessed March 2020.

Matute-Bello G, Liles WC, Radella F 2nd, *et al*. Modulation of neutrophil apoptosis by granulocyte colony-stimulating factor and granulocyte/macrophage colony-stimulating factor during the course of acute respiratory distress syndrome. Crit Care Med 2000;28:1-7.

McGonagle D, Sharifa K, O'Regand A, Bridgewood. The role of cytokines including interleukin-6 in COVID-19 induced pneumonia and macrophage activation syndrome-like disease. Autoimmunity Rev 2020:102537. doi: 10.1016/j.autrev.2020.102537

Mehta P, McAuley DF, Brown M, *et al*. COVID-19: consider cytokine storm syndromes and immunosuppression. Lancet 2020;395(10229):1033-4.

Molfino NA, Kuna P, Leff JA, *et al*. Phase 2, randomised placebo-controlled trial to evaluate the efficacy and safety of an anti-GM-CSF antibody (KB003) in patients with inadequately controlled asthma. BMJ Open 2016;6:e007709. doi:10.1136/bmjopen-2015-007709.

Overgaard CE, Schlingmann B, Dorsainvil White S, *et al*. The relative balance of GM-CSF and TGF-β1 regulates lung epithelial barrier function. Am J Physiol Lung Cell Mol Physiol 2015;308:L1212-23.

Paine R 3rd, Standiford TJ, Dechert RE, *et al*. A randomized trial of recombinant human granulocyte-macrophage colony stimulating factor for patients with acute lung injury. Crit Care Med 2012;40:90-7.

Paine R 3rd, Standiford TJ, Dechert RE, *et al*. A randomized trial of recombinant human granulocyte-macrophage colony stimulating factor for patients with acute lung injury. Crit Care Med 2012;40:90-7.

Paine R, Morris SB, Jin H, *et al*. Impaired functional activity of alveolar macrophages from GM-CSF-deficient mice. Am J Physiol Lung Cell Mol Physiol 2001*;*281: L1210–8.

Patnaik MM, Sallman DA, Mangaonkar A, *et al*. Phase 1 study of lenzilumab, a recombinant anti-human GM-CSF antibody, for chronic myelomonocytic leukemia (CMML). Blood 2020 Apr 15. pii: blood.2019004352. doi: 10.1182/blood.2019004352.

Roche. Press release. https://www.roche.com/media/releases/med-cor-2020-03-19.htm.

Sachdeva M, Duchateau P, Depil S, *et al.* Granulocyte-macrophage colony-stimulating factor inactivation in CAR T-cells prevents monocyte-dependent release of key cytokine release syndrome mediators J Biol Chem. 2019;294(14):5430-37.

Sakagami T, Beck D, Uchida K, *et al*. Patient-derived granulocyte/macrophage colony-stimulating factor autoantibodies reproduce pulmonary alveolar proteinosis in nonhuman primates. Am J Respir Crit Care Med 2010;182:49-61.

Sanofi. Press release. https://www.sanofi.com/en/media-room/press-releases/2020/2020-04-27-12-58-00.

Shah DK, Betts AM. Antibody biodistribution coefficients. mAbs 2013;5:297-305.

Siddiqi HK, Mehra MR. COVID-19 illness in native and immunosuppressed states: a clinical-therapeutic staging proposal, Journal of Heart and Lung Transplantation 2020; doi: 10.1016/j.healun.2020.03.012.

Stanley E, Lieschke GJ, Grail D, *et al*. Granulocyte/macrophage colony-stimulating factor-deficient mice show no major perturbation of hematopoiesis but develop a characteristic pulmonary pathology. Proc Natl Acad Sci USA 1994;91:5592-6.

Sterner RM, Sakemura R, Cox MJ *et al.* GM-CSF inhibition reduces cytokine release syndrome and neuroinflammation but enhances CAR-T cell function in xenografts. Blood 201914; 20194;133:697–709.

Taylor PC, Saurigny D, Vencovsky J, *et al.* Efficacy and safety of namilumab, a human monoclonal antibody against granulocyte-macrophage colony-stimulating factor (GM-CSF) ligand in patients with rheumatoid arthritis (RA) with either an inadequate response to background methotrexate therapy or an inadequate response or intolerance to an anti-TNF (tumour necrosis factor) biologic therapy: a randomized, controlled trial. Arthritis Res Ther 2019;21:101.

Trapnell BC, Nakata K, Bonella F, *et al*. Pulmonary alveolar proteinosis. Nat Rev Dis Primers 2019;5:16.

Vincent JL Moreno R, Takala J, *et al*. The SOFA (Sepsis-related Organ Failure Assessment) score to describe organ dysfunction /failure. On behalf of the Working Group on Sepsis-Related Problems of the European Society of Intensive Care Medicine. Intensive Care Med 1996;22:707-10.

WHO. https://www.who. int/blueprint/priority-diseases/key-action/COVID-19_Treatment_Trial_Design_Master_Protocol_synopsis_Final_18022020.pdf, 2020

Wicks IP, Roberts AW. Targeting GM-CSF in inflammatory diseases. Nat Rev Rheumatol 2016;12:37-48.

Wu C, Chen X, Cai Y, *et al*. Risk factors associated with acute respiratory distress syndrome and death in patients with coronavirus disease 2019 pneumonia in Wuhan, China. JAMA Intern Med 2020; doi: 10.1001/jamainternmed.2020.0994.

Xu X, Han M, Li T. *et al*. Effective treatment of severe COVID-19 patients with tocilizumab. chinaXiv:202003.00026v1. 2020.

Yang X, Yu Y, Xu J, *et al*. Clinical course and outcomes of critically ill patients with SARS-CoV-2 pneumonia in Wuhan, China: a single-centered, retrospective, observational study. Lancet Respir Med 2020 Feb 24. pii: S2213-2600(20)30079-5. doi: 10.1016/S2213-2600(20)30079-5.

Zhou Y, Fu B, Zheng X, *et al*. Aberrant pathogenic GM-CSF+ T cells and inflammatory CD14+CD16+ monocytes in severe pulmonary syndrome patients of a new coronavirus. 2020; bioRxiv preprint doi: https://doi.org/10.1101/2020.02.12.945576.
